# Phase 1 durability study of a single dose of the live-attenuated cholera vaccine PanChol and the safety and immunogenicity of a second dose

**DOI:** 10.64898/2026.10.13.26365415

**Authors:** Deborah R. Leitner, Stephen R. Walsh, Amy C. Sherman, Alisse Hannaford, Sudhir Doranga, Masataka Suzuki, George I. Balazs, Elliot Hammerness, Kaylee T. Nguyen, Natalie E Izaguirre, Akash R. Sinha, Mariana I. Haugh-Klockner, Alexandra Tong, Xiaofang Li, Damien M. Slater, Nirajan Puri, Jason B. Harris, Steven Piantadosi, Lindsey R. Baden, Matthew K. Waldor

## Abstract

**Background:** Killed whole-cell oral cholera vaccines require multiple doses and have reduced efficacy in young children, limiting their utility. PanChol is a single-dose, oral live-attenuated cholera vaccine engineered from a current seventh pandemic El Tor O1 *Vibrio cholerae* strain. PanChol was shown to be safe and immunogenic in a previous phase 1a trial. In participants from this first-in-human trial, we aimed to assess the durability of the initial PanChol dose and the safety and immunogenicity of a second dose.

**Methods:** This open-label phase 1a booster study was conducted at Brigham and Women’s Hospital (Boston, MA). Eligible participants were healthy adults aged 18–55 years without a history of cholera or gastrointestinal disorders and had received a PanChol dose ranging from 10^5^-10^10^ colony-forming units (CFU) within the previous 322-1111 days. All participants received a booster dose of 2×10^7^ CFU. The primary outcome was safety, including solicited, unsolicited, and serious adverse events following the second dose of PanChol. Secondary endpoints were the magnitude of pre- and post-vaccination serum vibriocidal titers to both Inaba and Ogawa *V. cholerae*, including seroconversion (4-fold rise in titers over baseline), and stool shedding of PanChol organisms using quantitative and qualitative stool cultures. Comparisons of the amount of vaccine shedding in stool following primary and secondary immunization were used to assess the intestinal mucosal immunity elicited by the initial PanChol dose. All outcomes were assessed in all participants. This trial is registered with ClinicalTrials.gov, NCT05657782, and is complete.

**Findings:** Between Aug 25, 2025, and Feb 20, 2026, 20 healthy adults who previously received a single oral dose of PanChol were enrolled to receive a booster dose of 2×10^7^ CFU. Eleven (55%) of 20 participants were male and the median age was 43·6 (IQR 30·2-49·5). Ten (50%) were White and 17 (85%) were not Hispanic or Latino. Nine (45%) of 20 boost recipients reported at least one solicited adverse event; 13 (93%) of 14 events were mild, and all were transient. The most common solicited adverse event was nausea, reported by 4 (20%) participants; all cases were mild. Most unsolicited adverse events were mild; one was grade 3 and was deemed unrelated to vaccination. Two unsolicited adverse events were deemed related to vaccination (mild gassy sensation and heartburn, both of which occurred on day 1 and resolved the same day). In most participants, vibriocidal antibody titers measured on the day of boosting exceeded baseline titers prior to the initial dose (17 (85%) for Inaba and 14 (70%) for Ogawa). Fifteen (75%) of 20 boost recipients seroconverted vibriocidal antibody titers to both serotypes. After the boost, 17 (85%) participants had no or self-limited vaccine shedding, compared with 10% after the first PanChol dose.

**Interpretation:** A second oral dose of PanChol had a favorable safety profile and induced vibriocidal seroconversion to both serotypes in 75% of boost recipients. Intestinal colonization of PanChol following the boost serves as a proxy for measuring mucosal immunity targeting *V. cholerae*. The decrease in in the abundance of PanChol fecal shedding following the second dose, compared with the first, suggests that a single dose of PanChol is sufficient to confer durable protective immunity against a cholera challenge. These observations underscore the need for further clinical development of PanChol.

**Research in context:** *Evidence before this study:* Vaccines are important tools for cholera control but current killed oral cholera vaccines (OCVs) require multiple doses for maximum efficacy and have limited efficacy in young children. In adults, protection mediated by killed OCVs is ∼55% 1 year after vaccination and lower in children. While estimates of the duration of protection afforded by natural infection vary, it is generally thought that immunity following infection is more durable across all age groups. PanChol is a live-attenuated cholera vaccine derived from a contemporary El Tor *V. cholerae* strain expressing both Inaba and Ogawa serotype antigens that was recently found to be safe and immunogenic in a first-in-human trial. We searched PubMed from database inception to September 1, 2026, for studies evaluating the durability of immunity to cholera following vaccination using the terms “duration of immunity to cholera vaccines”, yielding 11 non-review clinical trial articles. The search was limited to the English language. Previous challenge studies have not investigated the durability of protection from intestinal colonization and shedding beyond 6 months post-vaccination.

*Added value of this study:* A second dose of PanChol was administered 322-1111 days after participants had received an initial oral dose. The second dose of the vaccine was safe and well-tolerated, and 15 (75%) of 20 boost recipients seroconverted vibriocidal titers to both Inaba and Ogawa serotypes. Vaccine shedding was monitored to gauge PanChol intestinal replication. In striking contrast to primary vaccination, where 95% of participants shed the vaccine, only 30% of booster recipients shed PanChol and half of these individuals had self-limited shedding.

*Implications of all the available evidence:* A single dose of PanChol elicited mucosal immune responses that prevented *V. cholerae* intestinal colonization for up to 3 years post-vaccination. These observations suggest that a single dose of PanChol could limit *V. cholerae* transmission within communities by blocking *V. cholerae* intestinal colonization. PanChol’s favorable safety profile and stimulation of long-lived mucosal immunity support its continued development as a new single-dose agent for cholera control. Given these positive findings in adults in a non-endemic setting, future studies of the vaccine’s performance in cholera-endemic regions and in a variety of age groups are warranted. Further advancement of PanChol will require the development of a scalable manufacturing process to produce a heat-stable product that can be easily deployed in cholera-endemic areas.

## Introduction

Cholera continues to be a major global health challenge. There are an estimated 1·3-4 million cases and tens of thousands of deaths annually from this acute diarrheal disease ^1^. Cholera disproportionately affects the world’s most impoverished and vulnerable populations, particularly those lacking access to clean water and sanitation. Currently, ∼1·3 billion people are at risk of contracting cholera ^1^. While ensuring universal access to safe water and the provision of improved sanitation systems remain critical goals for eliminating this ancient scourge, cholera vaccines can serve as an effective intervention and are a key tool for cholera prevention and outbreak control ^2^.

Oral whole-cell killed cholera vaccines are currently the only WHO prequalified cholera vaccines authorized for use in endemic areas or during outbreaks. These vaccines are approved in a two-dose regimen and provide an average two-dose efficacy of 55% 12 months after vaccination in adults, which declines to 44% after 4 years. However, in children younger than 5 years old, the pooled two-dose efficacy was 31% with a 36-month weighted mean follow-up period ^3^. In response to the global cholera vaccine shortage, recent trials of a single dose of a killed cholera vaccine demonstrated an average efficacy of 57% at 6 months and 58% at 24 months among individuals aged 5 years and older. In contrast, this regimen provided only 16% protection among children under 5 years at 6 months and 13% after 24 months ^3^. Thus, killed cholera vaccines are less efficacious in children younger than 5 years, the demographic group that bears the highest burden of cholera ^4^.

In contrast to killed vaccines, natural infection with the cholera pathogen, the gram-negative rod *Vibrio cholerae*, generally confers protection against subsequent disease across all age groups ^5,6^. Based on these epidemiologic observations, we developed PanChol, a novel oral live-attenuated cholera vaccine. PanChol was derived from a contemporary El Tor biotype O1 serogroup *V. cholerae* strain that is the cause of the ongoing seventh pandemic of cholera. The strain was engineered to express both the Inaba and Ogawa *V. cholerae* O1 serotype antigens and to over-express the non-toxic B subunit of cholera toxin ^7^. Furthermore, additional genetic modifications were introduced into PanChol to ensure its biosafety for both individuals and communities and to maintain its genetic stability. These alterations include a CRISPR system targeting the gene encoding the enzymatic A-subunit of cholera toxin, making the strain resistant to toxigenic reversion.

We recently assessed the safety, tolerability and immunogenicity of a single oral dose of PanChol in healthy volunteers in Boston, MA, USA ^8^. PanChol doses ranging from 10^5^ to 10^10^ CFU were safe and well-tolerated. One hundred percent of participants who received PanChol at any of these doses seroconverted vibriocidal antibody responses to both the Inaba and Ogawa *V. cholerae* serotypes. PanChol shedding in feces, a marker for vaccine replication in the intestine, was detected in 96% of recipients who received at least 10^5^ CFU of PanChol. Despite its intestinal replication, the vaccine’s *in vivo* genetic stability was confirmed by whole-genome sequencing of vaccine cells obtained from fecal samples. To study the safety and immunogenicity of a second dose of PanChol, we conducted a PanChol boost study. Moreover, since the abundance of PanChol fecal shedding reflects *V. cholerae* (PanChol) intestinal colonization, shedding after the boost indicated the durability of the intestinal mucosal immunity stimulated by the initial PanChol dose. In this trial, individuals who had received an initial dose of PanChol 1-3 years previously were administered a second oral dose of the vaccine. This study design enables direct comparison of PanChol safety, immunogenicity, and shedding following primary and secondary vaccination within the same individuals. The second dose of PanChol proved safe and immunogenic. Notably, compared to primary vaccination, there was a marked reduction in PanChol shedding following the boost, suggesting that a single dose of PanChol stimulates durable intestinal mucosal immunity that impedes *V. cholerae* colonization.

## Methods

### Study design and participants

This open-label, fixed-dose booster study was conducted at the Brigham and Women’s Hospital (Boston, MA, USA) as an addendum to the phase 1a, first-in-human trial. Participants who received a single dose of PanChol (ranging from 10^5^ to 10^10^ CFU) from the previous dose-escalation or the randomized, double-blind, placebo-controlled module (2×10^8^ and 2×10^7^ CFU) were enrolled to assess the safety and immunogenicity of a second dose of 2×10^7^ CFU PanChol. Participants were excluded if they had a fever within the past 7 days, a gastrointestinal illness, or diarrhea. The complete list of inclusion and exclusion criteria is available in the appendix pp 3-4. Female participants were required to use acceptable contraceptive methods as outlined in the inclusion criteria for at least 28 days before vaccination and until three months after receiving the booster dose. All participants provided written informed consent prior to enrollment. Participant demographic characteristics, including age, sex (male or female) assigned at birth, gender (male, female, intersex, transgender, prefer not to disclose or other), race (American Indian or Alaska Native; Asian; Black or African American; Native Hawaiian or Other Pacific Islander; White; or other) and ethnicity (Hispanic or Latino; or not Hispanic or Latino) were self-reported by participants.

The research protocol (appendix 2) was approved by the Mass General Brigham Healthcare Institutional Review Board (2022P000461) and the Institutional Biosafety Committee (2022B000015) and conducted under a US Food and Drug Administration (FDA) Investigational New Drug Application (28217). This study is registered with ClinicalTrials.gov (NCT05657782) and has been completed.

### Procedures

Participants were enrolled and admitted to the Center for Clinical Investigation at Brigham and Women’s Hospital, where they received second dose of PanChol. The vaccine was manufactured and vialed under standards of good manufacturing practice at Walter Reed Army Institute of Research (Silver Spring, MD, USA; lot number 2081) and stored at −80 °C. On the day of the booster vaccination, PanChol was diluted to a concentration of 2×10^7^ CFU in sodium bicarbonate buffer and administered orally. Each dose was independently confirmed through titration at the time of each volunteer’s administration. Throughout the study period, the vaccine’s viability was monitored and remained intact. During the inpatient period, blood and fecal samples were collected, vital signs were monitored daily, and reactogenicity (solicited adverse events) and unsolicited adverse events were recorded. If participants had no detectable PanChol organism in their stool by day 3, they were discharged on the succedding day after administration of their first doxycycline dose (initial dose 200 mg, followed by 100 mg twice a day for 4 days). Participants with detectable vaccine shedding received doxycycline on day 5 and were discharged once a negative stool sample was documented. Participants discharged before day 7 were contacted daily by study staff to assess adverse events until day 7, the initial outpatient visit. Subsequent outpatient visits were scheduled for days 15, 29, 57, 85, and 180. During these visits, additional blood and stool samples were collected, and adverse events were assessed. All unsolicited adverse events were collected up to day 29; medically attended adverse events, new onset chronic medical conditions, and serious adverse events were collected up to day 180. Adverse events were graded in accordance with the FDA Toxicity Grading Scale for Healthy Adult and Adolescent Volunteers Enrolled in Preventive Vaccine Clinical Trials, September 2007. Laboratory procedures are listed in the appendix 1 p 4.

### Outcomes

The primary safety outcome was the incidence of solicited and unsolicited adverse events, including serious adverse events, following a second oral dose of PanChol.

Secondary outcomes were the magnitude of pre- and post-vaccination serum vibriocidal titers to both Inaba and Ogawa *V. cholerae*, the evaluation of seroconversion (4-fold rise in titer over baseline) of vibriocidal titers to both *V. cholerae* serotypes at 7 days post-vaccination (day 7) as well as direct assessment of stool shedding of PanChol organisms using both quantitative and qualitative stool cultures.

Exploratory outcomes were measurements of IgM, IgG, and IgA antibodies in serum and antibody-secreting cell responses (antibodies in lymphocyte supernatant (ALS)) targeting Inaba and Ogawa O-specific polysaccharides (OSP), cholera toxin B-subnit (CT-B) and TcpA, the major subunit of the pathogen’s critical colonization pilus via multiplex bead assay (Luminex).

### Statistical analysis

Safety analyses included all particiapnts who received a booster dose. The incidence of any adverse event, whether solicited, unsolicited, or serious, was determined. The number of participants with an adverse event was calculated, both overall and categorized by grade and relationship with a secondary vaccination. The immunogenicity data is presented as the magnitude of antibody responses in all boost recipients. A comparison of peak vibriocidal titres between primary and secondary vaccination was performed using Wilcoxon test on log-transformed data. Stool shedding and the measurement of IgM, IgG, and IgA antibodies in serum and lymphocyte supernatants were evaluated in all participants. GraphPad Prism (version 11) was used for all statistical analyses. Values for missing safety and immunogenicity data were not imputed and were excluded from the calculation of summary statistics for that datapoint.

### Role of funding source

The funder of the study had no role in study design, data collection, data analysis, data interpretation, or writing of the report.

## Results

Between Aug 25, 2025, and Feb 20, 2026, 20 participants who previously received an initial PanChol dose of 10^5^ CFU (2 at 10^5^, 1 at 10^6^, 6 at 10^7^, 9 at 10^8^ and 2 at 10^10^ CFU) were re-enrolled and admitted (figure 1). All participants received a second dose of 2×10^7^ CFU of PanChol. The time interval between the primary and secondary doses varied, with a median of 502 days (ranging from 322 to 1111 days) (figure 1). Most participants (13 [65%]) received the boost 1-2 years after the initial dose. All 20 (100%) participants completed 6 months of follow-up. 11 (55%) participants were male and 9 (45%) were female, with a median age of the full cohort of 43·6 years (IQR: 30·2-49·5). Most participants were White (10 [50%]) and not Hispanic or Latino (17 [85%]; table 1).

**Figure 1:**
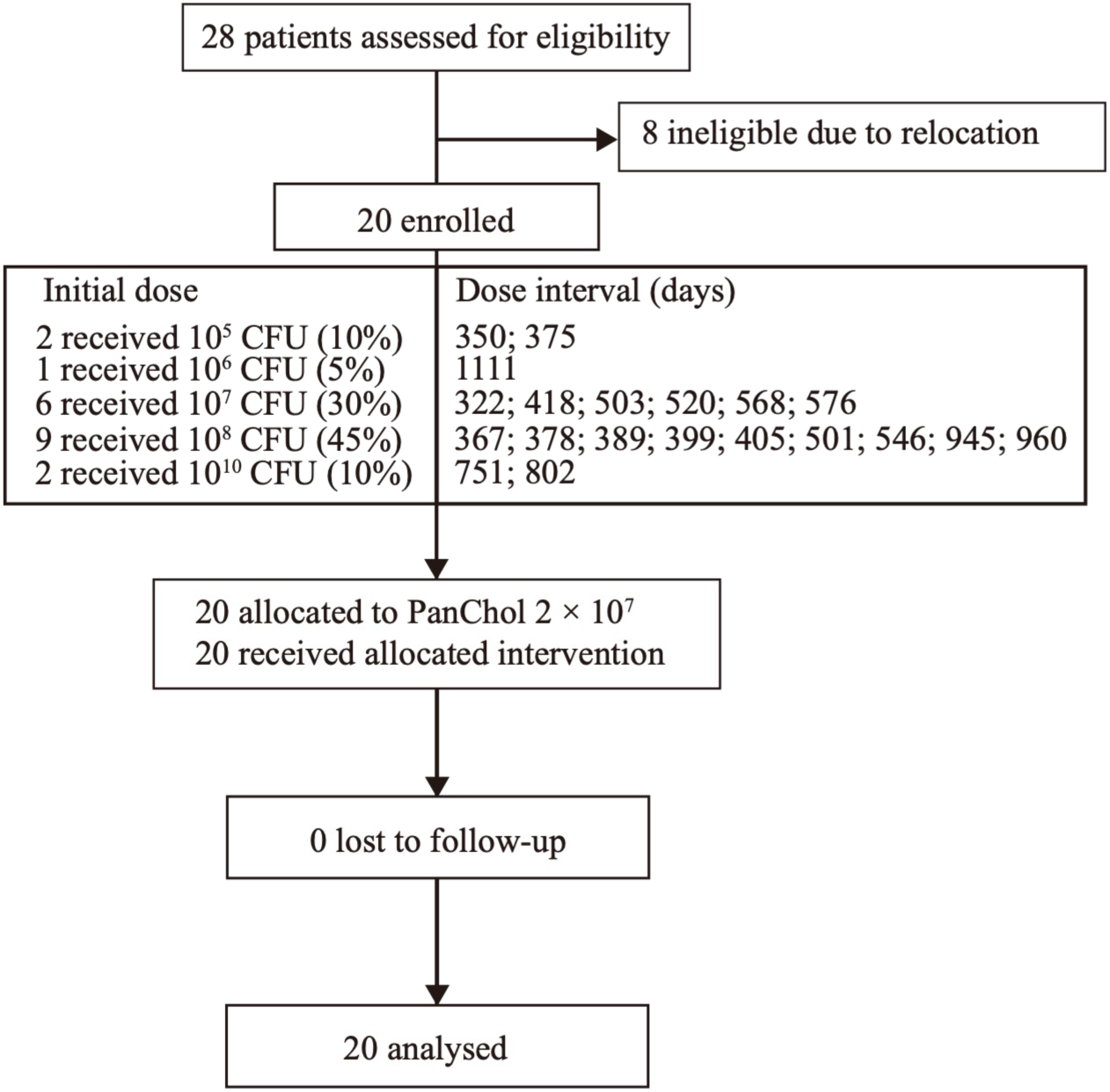
Trial profile. CFU=colony-forming units.

**Table 1:**
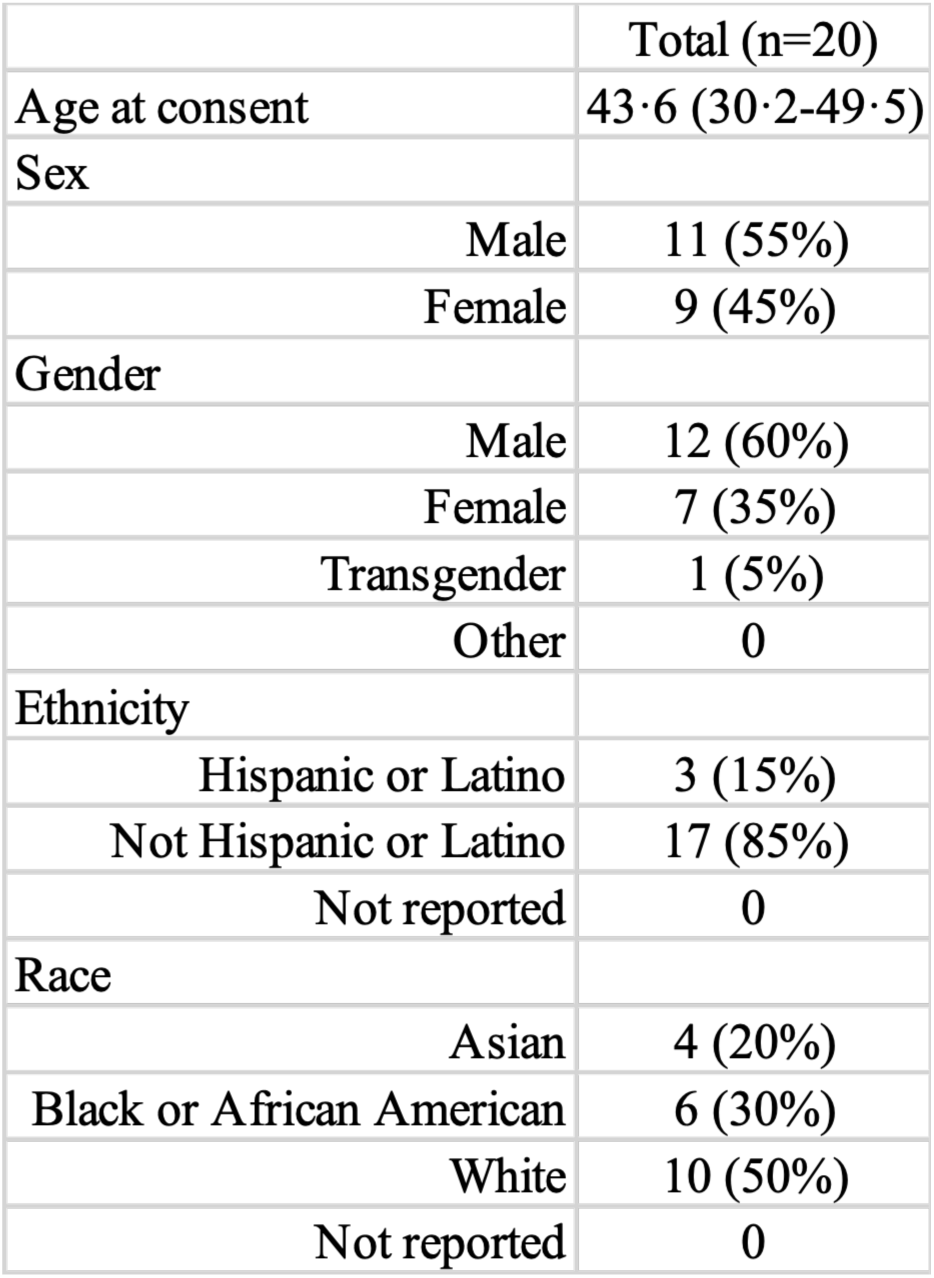
Baseline characteristic. Data are median (IQR) or n (%).

|  |  |
| --- | --- |
|  | Total (n=20) |
| Age at consent | 43·6 (30·2-49·5) |
| Sex |  |
| Male | 11 (55%) |
| Female | 9 (45%) |
| Gender |  |
| Male | 12 (60%) |
| Female | 7 (35%) |
| Transgender | 1 (5%) |
| Other | 0 |
| Ethnicity |  |
| Hispanic or Latino | 3 (15%) |
| Not Hispanic or Latino | 17 (85%) |
| Not reported | 0 |
| Race |  |
| Asian | 4 (20%) |
| Black or African American | 6 (30%) |
| White | 10 (50%) |
| Not reported | 0 |

The second dose of PanChol was safe and well tolerated (figure 2, appendix 1 p 8). Of the 20 participants, 11 (55%) had no solicited adverse events during the first 4 days after the second dose. Among nine vaccine recipients with solicited adverse events, 14 events were reported: 13 (93%) were mild, and one (7%), a headache, was moderate. There were a greater number of solicited adverse events among these 20 participants following the initial PanChol dose than after the booster dose (28, primary vs 14, boost) (figure 2). While abdominal pain was the most common solicited adverse event in primary vaccinees (7 [35%] of 20 primary vaccine recipients, all cases were mild), nausea (4 [20%] of 20 vaccine recipients, all cases were mild) was the most frequently reported solicited adverse event after the booster dose. Five of 20 participants (25%) experienced an adverse reaction after both vaccine doses. All solicited adverse events were transient and resolved within 2 days.

**Figure 2:**
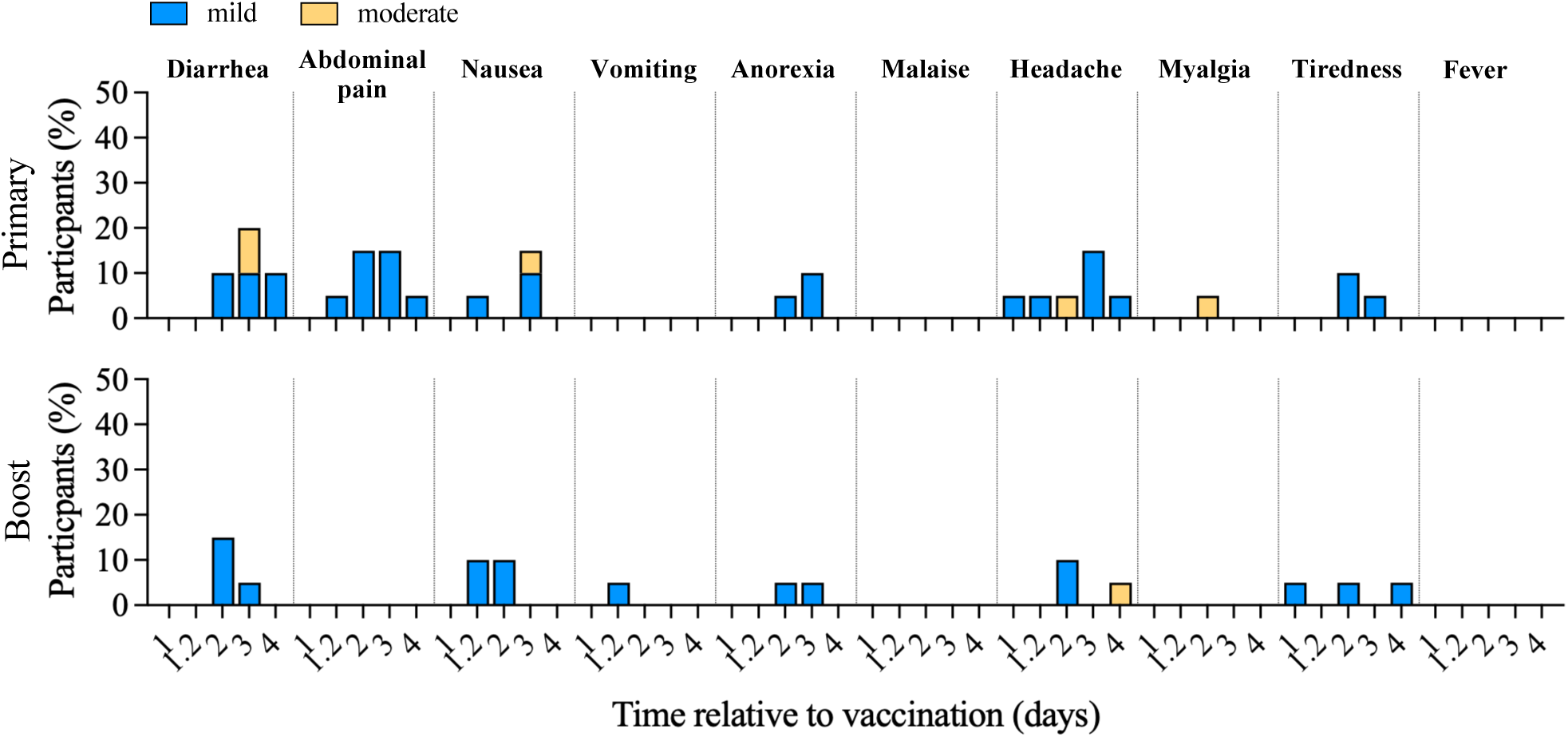
Solicited adverse events among study participants following primary vaccination and a booster dose. Recorded adverse events from hospital admission (day 1) until administration of doxycycline on day 5 (primary vaccination, top) or on day 4 (boost, bottom) are shown. Day 1·2 represents approximately 4 h post vaccination. Six participants received doxycycline on day 5 after the second dose.

Thirteen (65%) of 20 participants reported 31 unsolicited adverse events during the 6-month study period (appendix 1 pp 9-10). Two of these adverse events, mild gassy sensation and heartburn, were deemed related to the vaccine (both occurred on day 1 and resolved on the same day). Of the 29 unsolicited adverse events deemed unrelated to the treatment, 23 (79%) were mild (grade 1), 5 (17%) were moderate (grade 2) and 1 (4%) was severe (grade 3). The only adverse event more severe than grade 2 was an ear infection, which occurred on day 56 after vaccination.

Serum vibriocidal antibody titers against the two *V*. *cholerae* serotypes were measured to assess whether a second dose of PanChol stimulated changes in the abundance of these antibody responses, which can serve as a surrogate marker of protection from cholera ^9^. The same target strains - isogenic Inaba and Ogawa O1 *V. cholerae* strains ^10^ that were used for vibriocidal assays following the first dose of PanChol were used, facilitating direct comparisons of antibody titers and the kinetics of responses after primary and secondary immunizations.

Vibriocidal antibodies induced by primary vaccination and measured on the day of boosting were generally higher than the initial baseline titer prior to the initial PanChol dose, suggesting that a single dose of PanChol can stimulate antibody responses that remain detectable up to three years after primary vaccination (figure 3; appendix 1 pp 5, 11-12). Although pre-boost titers were elevated compared to baseline, 15 participants (75%) seroconverted following the second dose, indicating at least a fourfold increase in vibriocidal antibody titers against both serotypes relative to pre-boost titers (figure 3; appendix 1 pp 5, 11-12). Among the remaining five participants, four participants (80%) had a two-fold increase in their serotype-specific vibriocidal response against both serotypes. The kinetics of the vibriocidal antibody responses differed following primary and secondary immunization with PanChol. Vibriocidal titers peaked 15 days after the initial vaccination and 7 days after the second dose. Despite the slower time to peak titers following the initial dose, peak vibriocidal responses to both serotypes were significantly greater after the first dose of PanChol (day 15 geometric mean titers (GMT) for Inaba primary: 6089 (95% CI 3493-10614) and for Ogawa primary: 2473 (1137-5377)) than after the second dose (day 7 GMT for Inaba boost: 640 (295-1387) and for Ogawa boost: 469 (227-968), appendix 1 p13; p < 0·0001 for peak Inaba vibriocidal primary vs boost responses and p = 0·0024 for peak Ogawa vibriocidal primary vs boost responses; Wilcoxon test on log-transformed data, figure 3; appendix 1 pp 11-12). The lower peak vibriocidal responses after boosting may result from the more limited replication of the vaccine, see below.

**Figure 3:**
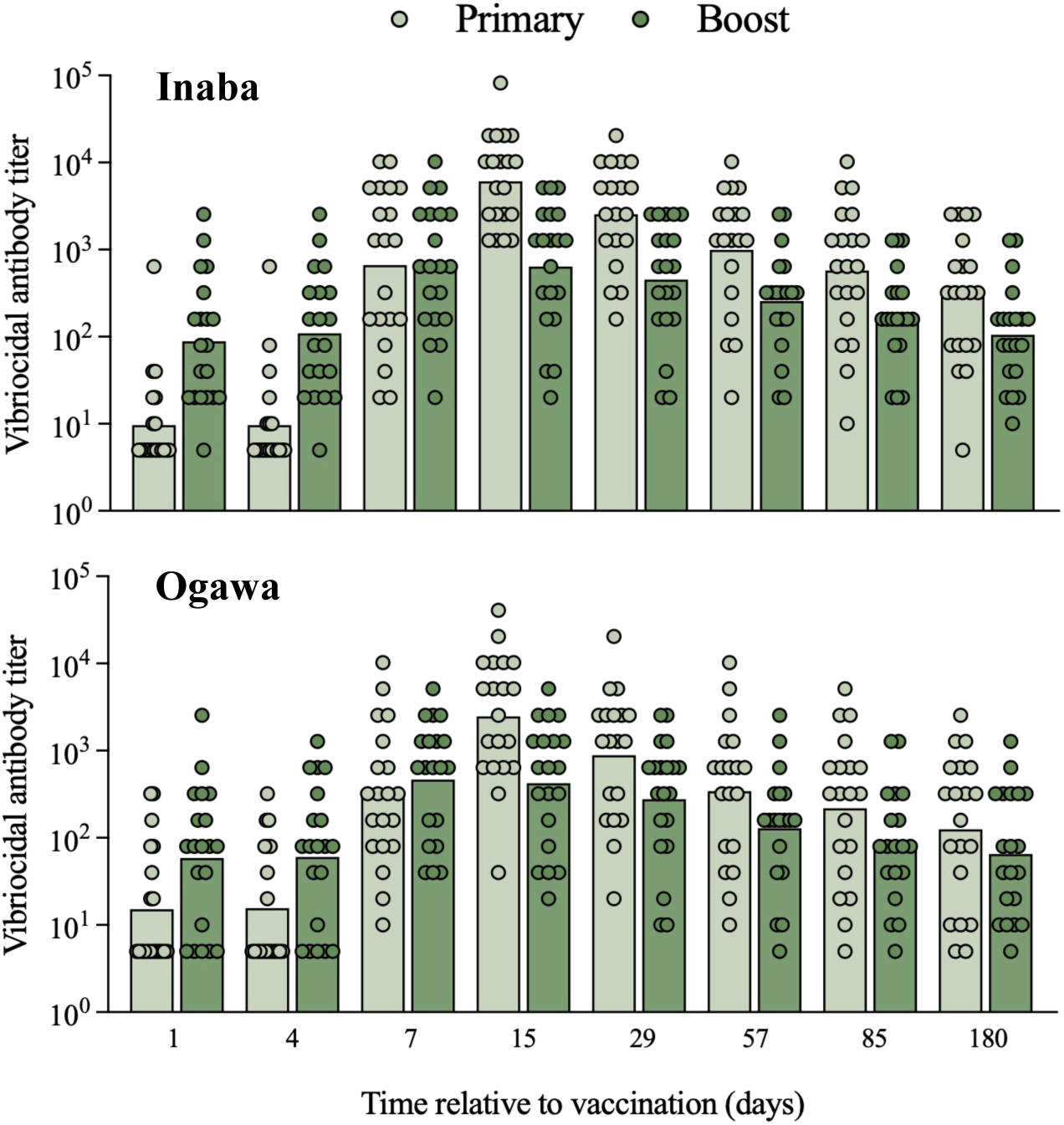
Serum vibriocidal antibody responses to Inaba and Ogawa *V. cholerae*. Temporal course of vibriocidal antibody titers starting at baseline (day 1) until study end (day 180) to Inaba (top) and Ogawa (bottom) *V. cholerae*. Primary responses are presented in light and boost responses in dark green. Bars represent geometric means.

Antigen-specific and isotype-specific serum responses to Inaba and Ogawa OSP, CT-B and TcpA after the second PanChol dose were compared to those after the first dose. While IgM responses to Inaba and Ogawa OSP were higher after the primary vaccination (appendix 1 pp 6, 14, 17), the magnitude of IgG and IgA responses to these antigens was greater following the boost (figure 4A-D, appendix 1 p 15-16, 18-19), reflecting isotype class switching, a characteristic of anamnestic antibody responses. The second PanChol dose also stimulated greater IgG and IgA responses to TcpA, the major *V. cholerae* colonization factor, compared to primary immunization (figure 4E-F, appendix 1 pp 24-25). However, the PanChol boost did not trigger greater elevations in isotype-specific antibody responses against CT-B compared to the first dose (appendix 1 pp 6, 20-22).

**Figure 4:**
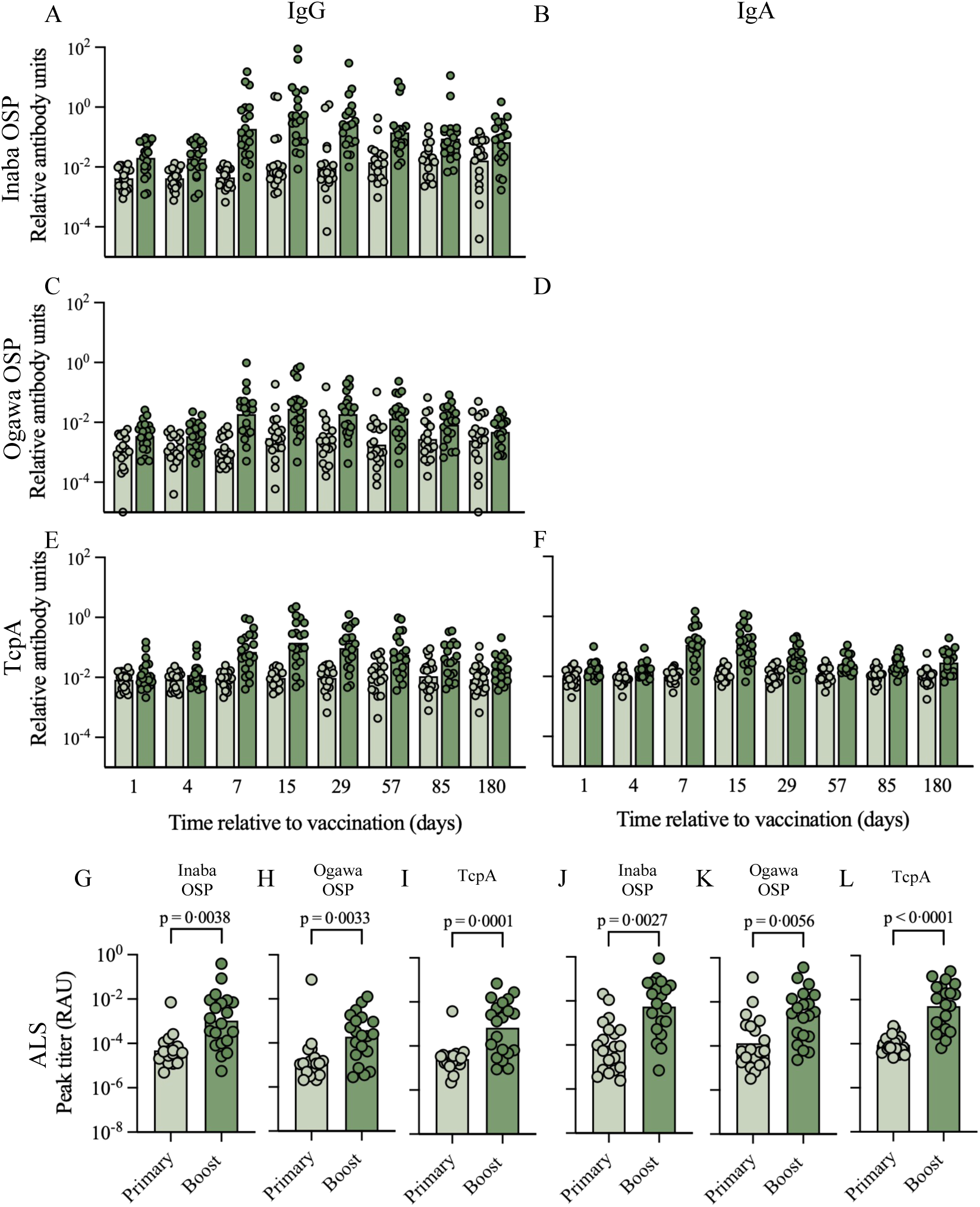
Antigen-specific and isotype-specific immune responses in serum and lymphocyte supernatant following primary and secondary vaccination. A-F) Relative antibody units of IgG (A,C and E) and IgA (B, D and F) immune response to Inaba OSP, Ogawa OSP and TcpA. G-L) Peak IgA (G-I) and peak IgG (J-L) responses in lymphocyte supernatants (ALS) to the indicated antigens. Values were calculated using each individual’s peak response. Data is presented as relative antibody units (RAU). Statistical comparisons of primary vs boost responses were performed using Wilcoxon test on log-transformed data. In all panels, the primary responses are presented in light and the boost responses in dark green. Bars represent geometric means.

The supernatants of antibody-secreting lymphocytes (ALS) were measured to assess antigen-specific and isotype-specific mucosal immune responses. Similar to serum responses, ALS IgG and IgA peak titers to Inaba and Ogawa OSP and to TcpA were greater after the second dose than after the first dose of PanChol (figure 4 G-L, appendix 1 pp 27-28, 30-31, 36-37). The IgM titers to Inaba and Ogawa OSP were not significantly altered after the boost, while the responses to TcpA were (appendix 1 pp 7, 26, 29, 35). Isotype-specific ALS titers to CT-B were similar after both doses (appendix 1 pp 7, 32-34).

Fecal shedding of PanChol was measured to assess the vaccine’s replication in the intestine. Shedding was monitored from 24 hours post-vaccination until doxycycline administration; in the current study, doxycycline was given on day 4 after the booster dose to allow participants to leave the hospital on day 5, whereas after the primary vaccination, doxycycline was administered on day 5, and most participants were discharged on day 7. Overall, PanChol shedding differed markedly in abundance and duration between the initial and second doses of the vaccine (figure 5A-C, appendix 1 p 38). After the initial dose, PanChol bacteria were detected in at least one fecal sample from 19 (95%) of the 20 participants and were detectable 4 days post vaccination in 18 (90%) of the 20 participants. In contrast, after the second dose, 14 (70%) of the 20 participants did not shed the vaccine at any point. Calculation of the total shedding per participant as an area under the curve (AUC) from day 2 to day 4 post-primary and secondary vaccination revealed a significant and marked reduction in fecal PanChol abundance post-booster vaccination (AUC geometric mean primary vaccination: 109238 CFU/g; AUC geometric mean booster vaccination: 1134 CFU/g). Furthermore, in 18 (90%) of the 20 participants, individual AUC values decreased after booster vaccination (figure 5D).

**Figure 5:**
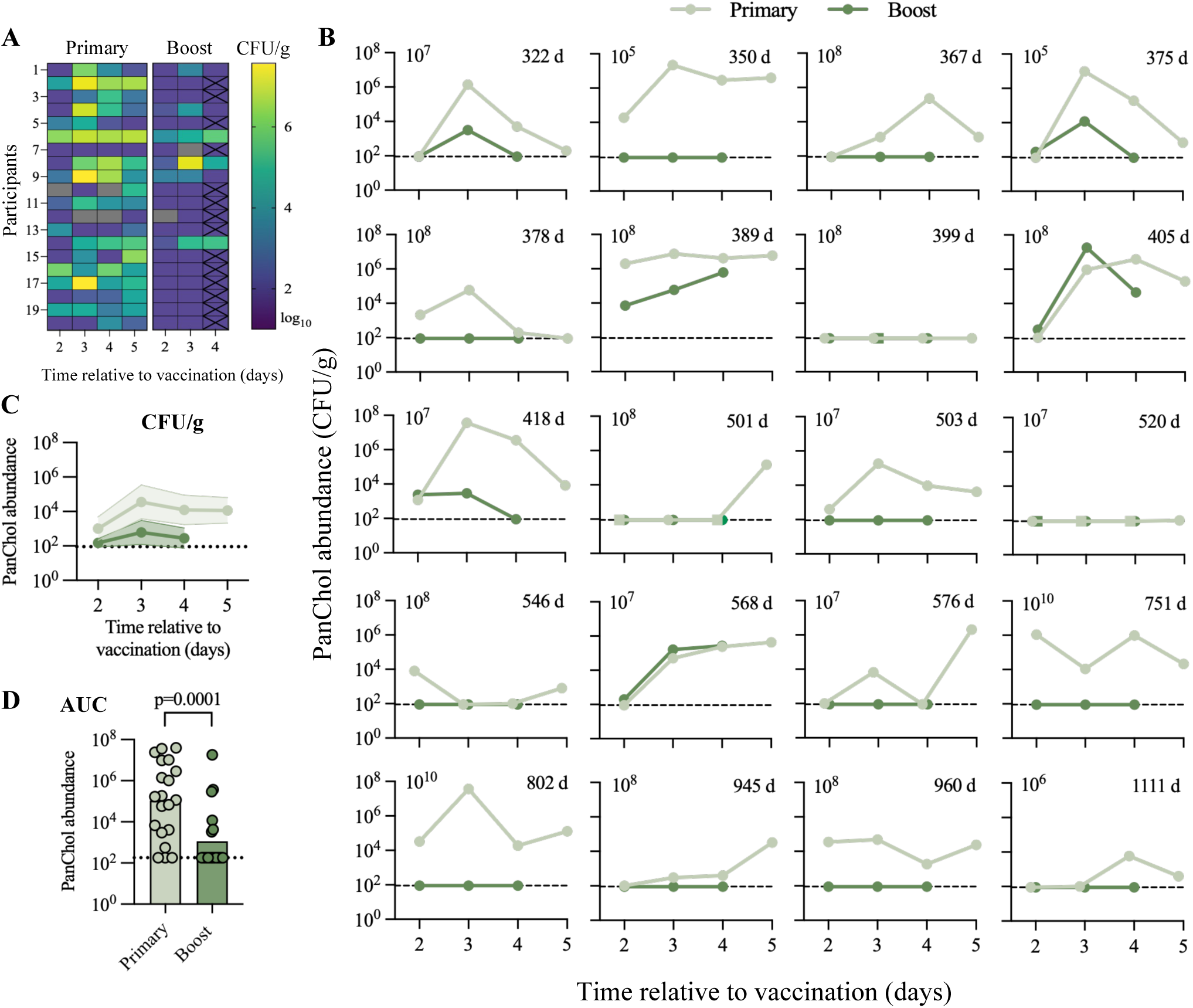
PanChol abundance in stool following primary and boost vaccination. A) Heatmap illustrating the number of PanChol CFU/g in stool samples collected between day 2 and 5 after primary vaccination and day 2 and 4 after the booster dose. Gray cells represent negative rectal swabs, and the black x on day four post-booster dose indicate no sample collection, with the result interpolated from the preceding day. B) Individual PanChol colonization patterns post-vaccination. The upper right corner represents the time interval between first and secondary dose, starting with the shortest interval. The initial dose is indicated in the upper left corner. The limit of detection is 100 CFU/g, and no shedding is represented as 90 CFU/g (dotted line). Circles represent quantitative stool cultures and squares indicate rectal swabs. C) Combined individual colonization patterns post-vaccination. Lines indicate geometric means and the filled area indicates the 95% CI. D) Comparison of the area under curve (AUC) following primary vaccination (day 2-4) and a booster dose (day 2-4). Statistical comparisons of AUC primary versus booster vaccination were performed using Wilcoxon test on log-transformed AUC data. Bars represent geometric means and the dotted line indicates no shedding. For B-D) Light green represents data collected after primary vaccination, and dark green after the second dose. Negative rectal swabs, shown as squares, are replaced with 90 CFU/g.

Among the six (30%) participants who shed the vaccine after the second dose the mean PanChol fecal CFU was ∼1·4-, 45·9- and 115·4-fold higher after the initial vs the second PanChol dose on days 2, 3, and 4 following vaccination respectively (GMT primary day 2: 748 (95% CI 11-49900) vs boost 514 (87-3045), primary day 3: 2372574 (207943-27070390) vs boost 51693 (1650-1619301) and primary day 4: 28990 (445-1889401) vs 4208 (46-381846)). Moreover, in 3/6 individuals, colonization was self-limiting and resolved prior to antibiotic treatment; thus, 17 of the 20 (85%) participants who received the boost had either no detectable shedding or self-limited shedding (figure 5AB, appendix 1 p 38).

Primary immunization with PanChol doses ranging between 10^5^ and 10^10^ CFU were all capable of eliciting responses that impeded PanChol-boost shedding. These observations are consistent with our previous observation that the size of the PanChol dose in the primary immunization does not correlate with the magnitude of immune responses, likely due to vaccine replication in the intestine. Furthermore, there was no apparent correlation between the abundance of vaccine shedding after the second dose and the time interval between primary and secondary immunizations, which ranged from 322 to 1111 days. In fact, none of the 4 participants who received the second PanChol dose at least 2 years after their initial vaccination shed PanChol after the boost, suggesting that the initial dose of PanChol can elicit long-lived mucosal protection against *V. cholerae* colonization.

## Discussion

Vaccines are recognized as key tools for control of both endemic and epidemic cholera. New vaccines, engineered from contemporary El Tor O1 *V. cholerae* strains, such as PanChol, can potentially overcome limitations of current killed oral vaccines by leveraging the enhanced immunogenicity elicited by *V. cholerae* replication in the intestine. Our previous phase 1a study of PanChol showed that a single dose was safe and immunogenic in healthy adults ^8^. Here, we investigated the safety and immunogenicity of a second dose of PanChol in 20 of the participants from the initial trial, representing 41% of the original 49 PanChol participants. All parameters measured initially, including adverse events, immune responses, and vaccine shedding in feces, were again assessed. Thus, this study design enables direct comparison of these outcomes following primary and secondary PanChol vaccination within the same individuals. In the first-in-human study, the safety profile of a single oral dose of PanChol across a wide range of doses (10^5^-10^10^ CFU) was established. The findings here, where all participants received a second dose of 2×10^7^ CFU after the primary vaccination, reinforce PanChol’s favorable safety profile, as fewer adverse events were reported following the boost.

The most salient difference observed among participants between the initial and second doses of PanChol was the marked reduction in vaccine shedding following the boost. After the boost, 85% of participants had no or self-limited vaccine shedding compared with 10% after the first PanChol dose. This striking difference strongly suggests that the initial PanChol dose stimulated a long-lived intestinal mucosal immune response that blocked *V. cholerae* intestinal colonization. Since participants received the boost up to three years after their primary vaccination, these observations suggest that a single dose of PanChol can trigger durable protective immunity against cholera.

Concordant with the idea that PanChol stimulates long-lived immune responses, we observed that vibriocidal titers measured on the day of the PanChol boost, which occurred approximately 1-3 years after primary vaccination, were higher than baseline titers prior to vaccination in most participants. Prior to boost, Inaba and Ogawa titers were elevated in 85% and 70% of participants respectively relative to baseline titers before primary vaccination; Inaba titers were 9·2−fold higher and Ogawa titers were 3·9−fold higher. These observations suggest that PanChol induces a more durable immune response than the live-attenuated Inaba vaccine CVD103-HgR. A study in which Swiss adults received a CVD103-HgR booster dose 15 or 24 months after primary immunization showed that the GMT of vibriocidal antibodies was similar to pre-vaccination titers ^11^. For an oral killed cholera vaccine, there was a 1·7- and 2·8-fold elevation in Inaba and Ogawa vibriocidal titers relative to pre-vaccination titers, one year after the two-dose vaccine regimen in participants in Kolkata ^12^, suggesting that the single dose of PanChol elicits a more durable immune response than the two-dose killed vaccine regimen. In Bangladesh, Harris et al found that vibriocidal titers returned to baseline one year after symptomatic culture-confirmed cholera ^13^. It will be of interest to determine whether PanChol elicits more durable vibriocidal responses in regions where cholera is endemic and populations are repeatedly exposed to the pathogen ^14^.

Despite elevated vibriocidal titers prior to the PanChol boost, the second vaccine dose further increased vibriocidal titers. Although PanChol expresses both Inaba and Ogawa antigens, there was a trend toward numerically higher Inaba responses, which was also observed after primary vaccination. Following the boost, vibriocidal titers peaked earlier than after the primary immunization. Fifteen (75%) of 20 PanChol boost recipients seroconverted their Inaba and Ogawa vibriocidal titers. Similar seroconversion against both serotypes was observed in a study of a Shanchol boost, a killed OCV, in Kolkata ^15^. Peak vibriocidal titers were lower than after the primary PanChol immunization. A relative diminution in peak vibriocidal titers was also observed following a boost with the live-attenuated vaccine CVD-103HgR, where only 29% (Inaba) and 23% (Ogawa) of participants seroconverted ^11^.

In addition to the accelerated peak in vibriocidal responses, additional manifestations of recall responses to the PanChol boost included isotype class-switched antigen-specific antibody responses in serum and in lymphocyte supernatants. As expected for a memory response following a vaccine boost, there were greater IgG and IgA titers to Inaba and Ogawa OSP, the major protective antigens, and TcpA, the major subunit of the pathogen’s critical colonization pilus, following the boost. In contrast, IgM responses to most antigens tested were less than those observed following primary vaccination. In general, recall antibody responses to CT-B were less robust than those detected for the other 3 antigens tested. IgG antibodies targeting CT-B increased in 45% of participants following the boost, whereas 80% of participants had elevated IgG titers to this cholera toxin subunit after primary immunization. Reduced frequency of IgG antitoxin responses was also reported in an experimental re-challenge with wild-type *V. cholerae* ^16^ and with CVD-103HgR ^11^.

PanChol shedding in feces, a marker of *V. cholerae* intestinal replication, was markedly reduced following the booster dose compared with primary vaccination. All boost recipients had either no or reduced vaccine shedding compared to their shedding following the initial vaccine dose. Reduction in PanChol (*V. cholerae*) colonization and fecal shedding following the second vaccine exposure has also been observed in studies of experimental cholera ^16^ and presumably reflects the durability of the intestinal mucosal response elicited by the initial dose of PanChol. There is no known threshold value of vibriocidal titers above which protection is guaranteed ^17^ and in our study, we did not find a clear correlation between serum vibriocidal titers at boost and subsequent shedding. Additional measures of mucosal responses that block *V. cholerae* colonization are required. Our study demonstrates the utility of using PanChol, a well-tolerated contemporary El Tor O1 *V. cholerae* strain, for measuring the effectiveness of mucosal immunity in impeding *V. cholerae* intestinal colonization, as an alternative to virulent human challenge strains. Similar applications of PanChol could be used in future studies to compare the efficacy of current and future cholera vaccines.

Most experimental challenge studies with *V. cholerae* primarily assess diarrhea and associated symptoms and do not provide a quantitative assessment of *V. cholerae* shedding. In a small study, Levine and colleagues found that 2 of 4 individuals rechallenged with El Tor Inaba *V*. *cholerae* ∼1-3 months after initial infection had positive stool cultures ^18^; similarly in a re-challenge study with classical *V. cholerae*, 1 of 4 individuals shed the pathogen after re-challenge ∼3 years after the initial infection ^16^. Three months after vaccination with a single dose of the live-attenuated vaccine CVD103HgR, 61% of people challenged with 10^5^ of a wild type *V. cholerae* strain shed the challenge strain although at a much lower level than controls ^19^. In the current study, even though we challenged participants with a 100-fold larger dose of a live-attenuated vaccine (PanChol) ∼1-3 years after initial exposure, no *V. cholerae* shedding was detected in 70% of participants. These observations suggest that a single-dose of PanChol, by blocking *V. cholerae* intestinal colonization, has the potential to block *V. cholerae* transmission.

The limitations of this study include its small sample size and its single-site, non-endemic setting, which may limit the generalizability of the findings. 20 of the 49 original PanChol recipients agreed to enroll in this trial, creating potential selection bias as well as some heterogeneity in the initial vaccine doses and dose-to-boost interval. Furthermore, the study design does not reflect the real-world vaccine use, as booster recipients received doxycycline on day 4 after vaccination to eliminate PanChol shedding. Reduced shedding after a second dose is an indirect marker of mucosal immunity to PanChol and may not reflect protection against wild-type *V. cholerae* colonization. A phase 1b study (NCT07107516) assessing the safety and immunogenicity of PanChol in Lusaka, Zambia, is ongoing and will further our understanding of PanChol safety and immunogenicity in a cholera-endemic setting and provide additional insight into the kinetics of vaccine shedding, as participants do not receive doxycycline until day 14 post-vaccination.

Given the emerging data demonstrating the safety, immunogenicity and long-term durability and potency of a single dose of PanChol, additional trials of this vaccine, particularly in children, the demographic group that bears the highest burden of cholera are warranted. In addition, the development of a scalable manufacturing process to create a heat-stable form of PanChol is critical to realize its promise to boost global cholera control measures.

## Data Availability

De-identified individual participant data and the data dictionary, or supporting clinical documents, will be made available upon request to the corresponding author after publication for at least five years. Proposals will undergo a scientific review by the PanChol protocol team, require a signed data access agreement and may involve institutional ethics committee review.

## Contributors

MKW, LRB, and SP conceptualized the study. DRL, SRW, ACS, AH, SD, MS, GIB, KTN, DMS, and LRB were involved in the data collection. All authors had access to all data and DRL, SRW, NP, SP, LRB, and MKW verified the data. DRL, NP, and SP did the statistical analysis. DRL, ARS and DMS conducted the laboratory analyses for the study. SRW, AH, ACS, EH, MHK, AT and NEI were involved in the clinical organization of this study. SRW, ACS, AH, and LRB were involved in protocol development and NP was the data manager. JBH, XL, LRB and MKW supervised aspects of this study. All authors contributed to the data interpretation, provided critical review, and approved the original version of the manuscript. All authors had final responsibility for the decision to submit for publication.

## Declaration of interests

MKW is an inventor on a patent related to PanChol. SRW has received institutional grants or contracts from Sanofi Pasteur, Janssen Vaccines/Johnson & Johnson, Moderna Tx, Pfizer, AbbVie, F2G, Hookipa, Vir Biotechnology, and Worcester HIV Vaccine; has participated on data safety monitoring or advisory boards for Janssen Vaccines/Johnson & Johnson, CyanVac, CSL Behring, and BioNTech; and his spouse holds stock/stock options in Regeneron Pharmaceuticals. ACS reports consulting fees from Bain Capital and Pfizer. All other authors declare no competing interests.

## Data sharing

De-identified individual participant data and the data dictionary, or supporting clinical documents, will be made available upon request to the corresponding author after publication for at least five years. Proposals will undergo a scientific review by the PanChol protocol team, require a signed data access agreement and may involve institutional ethics committee review. The protocol for this article is available in appendix 2 pp1-90.

## Acknowledgements

The authors wish to thank the participants for their involvement in the study. The authors wish to thank Jon A. Gothing, Kimberly Gaffney, Erin R. Curry, Virginia Dimalanta, Cynthia Mompremier, Jacquelyn Koser, Rhonda Furlonge, Virginia Cyr, Roselande Muscade, Vivian Tran, Lori Golden, Heather Rios, Caitlin Sugrue, Kevin M. Zinchuk, Stephanie Manners, Charles Kelly III, Emily Koleske, Esther Arbona-Haddad, Jose Humberto Licona, Abboud Kaok, Hannah Drackley and Caleb Lambert. We thank RSG member John J. Mekalanos for suggesting that a boost study could serve as proxy for mucosal immunity. Thank you to the Waldor lab for their feedback, particularly Ian W. Campbell and Franz G. Zingl on the project and comments on the manuscript.

This study is supported by Wellcome Trust (218443/Z/19/Z) to MKW and LRB. MKW is supported by NIH RO1AI042347-30 and the Howard Hughes Medical Institute (HHMI). LRB is supported by NIAID 5-UM1-AI-069412-19 and the Harvard Clinical and Translational Science Center (1UL1TR002541-01). ACS is supported by NIH 1K23AI185318-01A1. JBH is supported by NIAID R01AI179917.

This article is subject to HHMI’s Open Access to Publications policy. HHMI lab heads have previously granted a non-exclusive CC BY 4.0 license to the public and a sublicensable license to HHMI in their research articles. Pursuant to those licenses, the author-accepted manuscript of this article can be made freely available under a CC BY 4.0 license immediately upon publication.

## Funding

Wellcome Trust.

## Notes

### Clinical Trial

NCT05657782

### Author Declarations

The Institutional Review Board of Mass General Brigham gave ethical approval for this work.

