## Appendix 1 for "Phase 1 durability study of a single dose of the live-attenuated cholera vaccine PanChol and the safety and immunogenicity of a second dose"

**Full eligibility criteria**

To be eligible to participate in this study healthy volunteers must meet the following criteria within 56 days of Study Day 1 or at the time point specified in the individual eligibility criterion listed.

Inclusion Criteria

2. Must have given written informed consent (signed and dated) and any other authorizations required by local law and be able to comply with all study requirements.
3. Healthy adults aged from 18 to 55 years old.
4. Considered healthy, as judged by the clinical investigator, according to medical history, physical examination, vital signs, screening laboratories, and medication history.
5. Capable of understanding, consenting, and complying with the entire study protocol including the inpatient period.
6. Female participants must be non-pregnant and non-lactating and either
    a. surgically sterile (history of bilateral ligation, bilateral salpingectomy, bilateral oophorectomy, total hysterectomy) or postmenopausal (defined as as amenorrhea for at least 12 consecutive months before screening without an alternative medical cause)
    b. be of child-bearing potential and practicing an acceptable method of contraception or abstaining from all activities that could result in pregnancy for at least 28 days before vaccination until 3 months after receiving the IP.
    Acceptable methods of contraception include barrier methods (such as condom, diaphragm, or cervical cap used in conjunction with spermicide), intrauterine device, hormonal contraception (that may be taken or administered by oral, intravaginal, transdermal, subdermal or IM route), vasectomized partner (the vasectomized partner should be the sole partner for that participant)

Exclusion Criteria

1. Confirmed or suspected immunosuppressive condition, as a result of a disease (e.g., primary immune deficiency, malignancy, HIV infection) or have taken any systemic immunosuppressive therapy within 6 months of enrollment.
2. Pregnant or lactating women.
3. History of gastrointestinal (GI) disorder, such as previous major GI surgery, malabsorption, or any chronic GI disorders that would interfere, according to the investigator, with the IP.
4. Acute GI or febrile illness within 7 days of enrollment.
5. Have any acute or chronic medical condition that, in the opinion of the investigator, would make vaccination unsafe or interfere with the evaluation of immune response to study vaccination.
6. History of cholera vaccination prior to enrollment.
7. History of cholera infection.
8. Abnormal stool pattern, defined as < 3 or >21 stools per week.
9. Serious allergic reaction to PanChol or placebo component (sodium bicarbonate, lactose, ascorbic acid)
10. Use of any systemic antibiotics within 1 month of PanChol administration.
11. Receipt of a live vaccine in the previous 4 weeks or planned in the 4 weeks following enrollment.
12. Receipt of a killed or subunit (non-live) vaccine in the previous 2 weeks or planned in the 2 weeks following enrollment.
13. Individuals who do not speak English will not be enrolled into this trial. This study involves more than minimal risk and no prospect of direct benefit for participants. Additionally, a subject who did not speak English may not be able to easily communicate safety concerns in a timely fashion to the study investigators
14. Childcare workers with direct contact with children ≤ 2 years of age
15. Individuals whose occupation involves handling of food

16. Healthcare workers who have direct contact with patients who are immunodeficient, HIV-positive, or have an unstable medical condition
17. Use laxatives regularly
18. Have diarrhea within 48 hours before enrollment
19. Have a history of hypersensitivity to any of the tetracyclines
20. Have a history of hypersensitivity to streptomycin or any aminoglycoside due to the known cross-sensitivity of patients to drugs in this class.
21. Individuals who have a household member who are immunodeficient, HIV-positive, or have an unstable medical condition.

**Quantification of PanChol bacteria**

The quantification of PanChol bacteria in the vaccine dose and participants' stool samples was performed as previously described [1]. Every vaccine dose was independently titrated by plating on Luria-Bertani agar containing streptomycin (200 µg/mL) and X-gal (40µg/mL) to determine the number of PanChol CFU per 100 ml. To determine the number of PanChol CFU per gram of stool, one stool sample per day per study participant was collected during the inpatient period and serial dilutions were directly plated without enrichment on thiosulfate-citrate-bile salts-sucrose (TCBS) agar (Sigma) containing streptomycin (200 µg/mL). If no stool specimen was produced, a rectal swab was obtained and streaked on TCBS plates to monitor for the presence of PanChol.

**Vibriocidal antibody assay**

Quantification of serum vibriocidal antibodies using the isogenic Inaba and Ogawa ZChol *V. cholerae* strain*s* as targets assessed in serum collected before vaccination and on day, 7, 15, 29, 57, 85 and 180 days post vaccination was performed as previously reported [1,2]. Inaba and Ogawa vibriocidal responses were reported as the highest dilution of serum resulting in a ≥50% reduction in target optical density compared to control wells without serum. A titer of 5 was assigned when no vibriocidal antibody responses were detected. The mouse monoclonal antibody 432A.1G8.G1.H12 targeting *V. cholerae* O1 OSP was used as a positive control. A ≥4-fold increase in titer relative to baseline was considered seroconversion.

**Multiplex Bead Assay for isotype- and antigen-specific antibody responses**

Multiplex bead analysis was performed as described previously [1]. Briefly, serum was diluted 100-fold (final dilution, 1:1,000) and tested in duplicate on a FlexMap3D instrument (Luminex, Austin, TX). Each plate included a standard curve prepared from a pooled convalescent-phase sera reference. Relative antibody units were calculated from mean NetMFI values using logistic regression in GraphPad Prism, v10.4.0. A detailed version of the protocol can be found at: dx.doi.org/10.17504/protocols.io.3byl4b1x8vo5/v1

**Antibody in lymphocyte supernatant (ALS) assay**

Participants blood samples were collected and processed as previously described [1]. The culture supernatant of ten million cells was collected after 48h of incubation in 5% $CO_2$ incubator at 37 °C, centrifuged and stored at -80 °C until subsequent Luminex analysis.

**Figure S1: Individual vibriocidal responses to Inaba and Ogawa of study participants following primary and booster vaccination from baseline (day 1) until study end (day 180).** The initial dose is displayed in the left upper corner, and the dose interval in the right upper corner. Primary responses in light and boost responses in dark green.

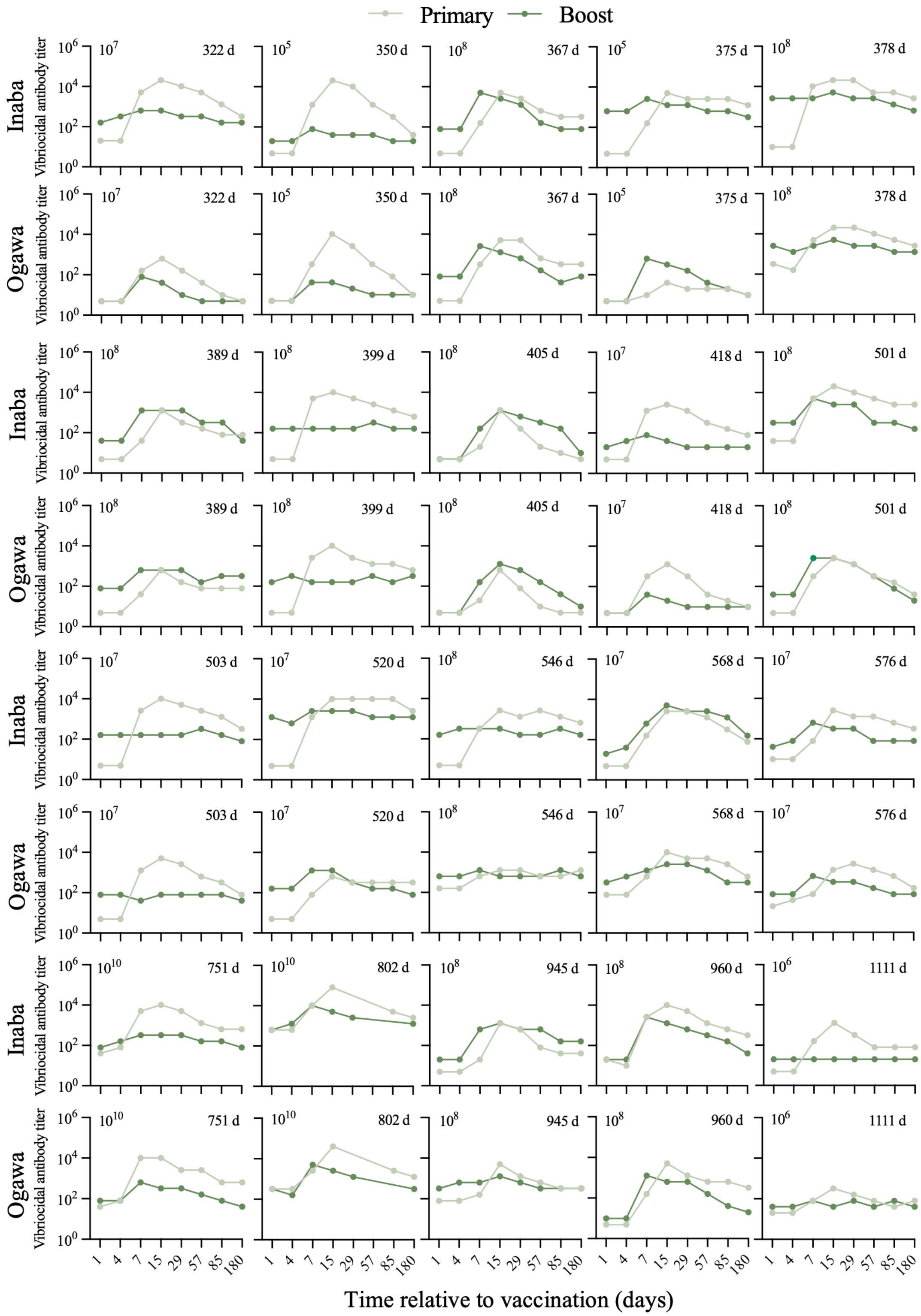

**Figure S2: Antigen-specific and isotype-specific immune responses in serum of study participants following primary and booster vaccination from baseline (day 1) until study end (day 180).** A-C) IgM specific immune responses to Inaba OSP (A), Ogawa OSP (B) and TcpA (C) after primary vaccination and boost. D-F) Isotype-specific responses to CT-B after primary and secondary vaccination. Data are represented as relative antibody units. For all graphs, bars represent geometric means, primary responses are presented in light green and secondary responses are indicated in dark green.

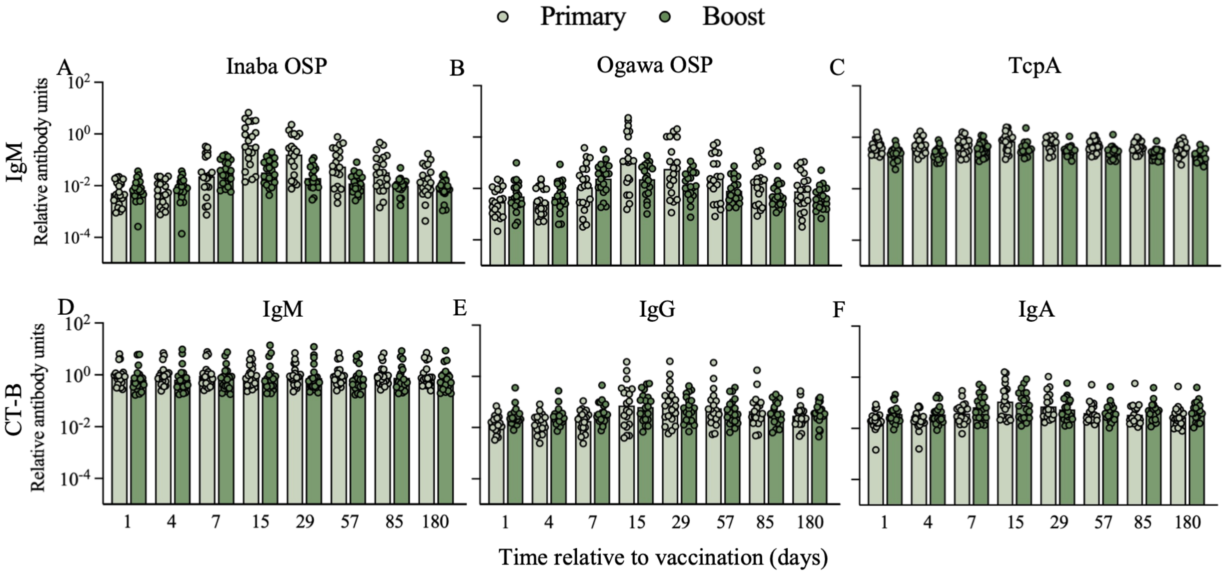

**Figure S3: Antigen-specific and isotype-specific immune responses in lymphocyte supernatants.** A-C) IgM specific peak antigen responses to Inaba OSP (A), Ogawa OSP (B), and TcpA (C) after primary vaccination and boost. D-F) Isotype-specific immune responses to CT-B after primary and secondary vaccination. Values were calculated using an individual's peak response. Data are represented as relative antibody units (RAU). Statistical comparisons of primary vs boost were performed using Wilcoxon test on log-transformed data. For all graphs, bars represent geometric means, primary responses are presented in light green and secondary responses are indicated in dark green.

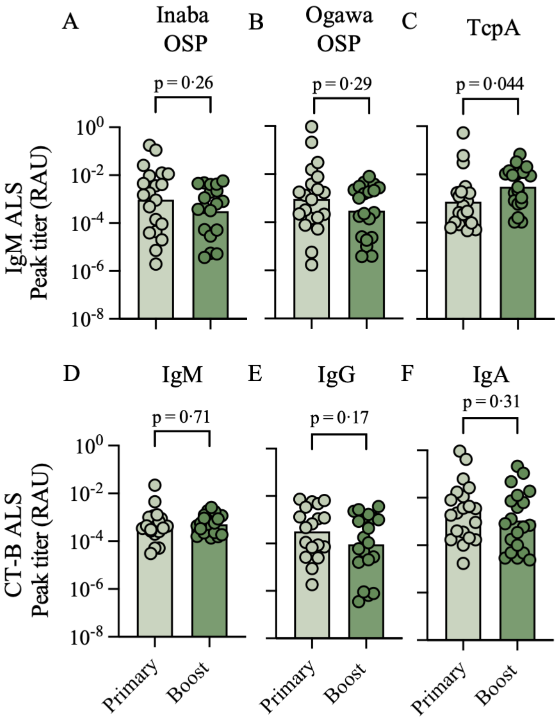

**Table S1: Solicited adverse events reported from participants after primary and booster vaccination during days 1 to 7.**

| | No. of participants with symptom/total no. of participants after primary and booster vaccination during days 1 to 7 | |
|---|---|---|
| **Symptom** | **Primary** | **Boost** |
| Diarrhea | | |
| Mild grade | 4/20 | 3/20 |
| Moderate grade | 2/20 | 0/20 |
| Severe grade | 0/20 | 0/20 |
| Abdominal pain | | |
| Mild grade | 7/20 | 0/20 |
| Moderate grade | 0/20 | 0/20 |
| Nausea | | |
| Mild grade | 5/20 | 6/20 |
| Moderate grade | 1/20 | 0/20 |
| Vomiting | | |
| Mild grade | 0/20 | 1/20 |
| Moderate grade | 0/20 | 0/20 |
| Anorexia | | |
| Mild grade | 3/20 | 2/20 |
| Moderate grade | 0/20 | 0/20 |
| Malaise | | |
| Mild grade | 0/20 | 1/20 |
| Moderate grade | 0/20 | 0/20 |
| Headache | | |
| Mild grade | 5/20 | 4/20 |
| Moderate grade | 2/20 | 1/20 |
| Myalgia | | |
| Mild grade | 0/20 | 0/20 |
| Moderate grade | 1/20 | 0/20 |
| Tiredness | | |
| Mild grade | 2/20 | 2/20 |
| Moderate grade | 0/20 | 0/20 |
| Fever | | |
| Mild grade | 0/20 | 0/20 |
| Moderate grade | 0/20 | 0/20 |

**Table S2A: Unsolicited adverse events greater than grade 2**

| Pub-ID | Dose | Verbatim Term | Days since vaccination | Maximum Severity Grade[1] | Relatedness |
|---|---|---|---|---|---|
| 100-10 | $2 \times 10^7$ | Double ear infection | 56 | 3 | Not related |

[1]Graded according to the FDA Toxicity Grading Scale for Healthy Adult and Adolescent Volunteers Enrolled in Preventive Vaccine Clinical Trials (September 2007)

**Table S2B: Unsolicited adverse events grade 2 and below**

| Pub-ID | Dose | Verbatim Term | Days since vaccination | Maximum Severity Grade[1] | Relatedness |
|---|---|---|---|---|---|
| 100-01 | $2 \times 10^7$ | Grade 1 (Mild) Elevated Bilirubin | 28 | 1 | Not related |
| 100-01 | $2 \times 10^7$ | Mild Tachypnea | 1 | 1 | Not related |
| 100-01 | $2 \times 10^7$ | Mild Elevated Bilirubin | 3 | 1 | Not related |
| 100-02 | $2 \times 10^7$ | Elevated ALT | 3 | 1 | Not related |
| 100-03 | $2 \times 10^7$ | Tachypnea | 1 | 1 | Not related |
| 100-03 | $2 \times 10^7$ | Bradycardia | 0 | 1 | Not related |
| 100-04 | $2 \times 10^7$ | Right cheek facial cyst | 18 | 2 | Not related |
| 100-04 | $2 \times 10^7$ | Right forearm tick bite | 11 | 1 | Not related |
| 100-05 | $2 \times 10^7$ | BD Increased creatinine | 14 | 1 | Not related |
| 100-05 | $2 \times 10^7$ | N/A Forehead Laceration | 54 | 2 | Not related |
| 100-06 | $2 \times 10^7$ | Sinus Infection | 33 | 2 | Not related |
| 100-06 | $2 \times 10^7$ | Reversible Coronary Ischemia | 80 | 1 | Not related |
| 100-06 | $2 \times 10^7$ | Non-radiating chest pain | 0 | 1 | Not related |
| 100-07 | $2 \times 10^7$ | Tachypnea | 1 | 1 | Not related |
| 100-08 | $2 \times 10^7$ | (N/A) Tachypnea (BD) | 1 | 1 | Not related |
| 100-08 | $2 \times 10^7$ | Increased AST (BD) | 3 | 1 | Not related |
| 100-08 | $2 \times 10^7$ | (N/A) Increased ALT (BD) | 3 | 1 | Not related |
| 100-09 | $2 \times 10^7$ | Hypertension (systolic) | 0 | 1 | Not related |
| 100-09 | $2 \times 10^7$ | Tachypnea | 0 | 1 | Not related |
| 100-10 | $2 \times 10^7$ | Hypertension (diastolic) | 14 | 1 | Not related |
| 100-11 | $2 \times 10^7$ | Upper respiratory infection | 50 | 1 | Not related |
| 100-11 | $2 \times 10^7$ | Upper respiratory infection | 28 | 1 | Not related |
| 100-11 | $2 \times 10^7$ | Viral Upper Respiratory Tract Syndrome | 5 | 1 | Not related |
| 100-11 | $2 \times 10^7$ | Heartburn | 3 | 1 | Not related |
| 100-11 | $2 \times 10^7$ | Dizziness | 1 | 1 | Not related |
| 100-11 | $2 \times 10^7$ | Backache | 12 | 2 | Not related |
| 100-11 | $2 \times 10^7$ | Gassy abdominal feeling | 1 | 1 | Related |
| 100-11 | $2 \times 10^7$ | Broken left big toe | 83 | 2 | Not related |
| 100-12 | $2 \times 10^7$ | Elevated AST | 1 | 1 | Not related |
| 100-13 | $2 \times 10^7$ | Heartburn | 1 | 2 | Related |

[1]Graded according to the FDA Toxicity Grading Scale for Healthy Adult and Adolescent Volunteers Enrolled in Preventive Vaccine Clinical Trials (September 2007)

**Table S3: Serum vibriocidal antibody responses (VAT) to Inaba and Ogawa *V. cholerae* O1 after primary and booster vaccination.**

Blue: no serum sample.

| Boost Inaba (VAT) | | | | | | | | | | |
|---|---|---|---|---|---|---|---|---|---|---|
| Dose interval (days) | Dose (CFU) | D1 | D4 | D7 | D15 | D29 | D57 | D85 | D180 | max |
| 322 | $2\times10^7$ | 160 | 320 | 640 | 640 | 320 | 320 | 160 | 160 | 640 |
| 350 | $2\times10^7$ | 20 | 20 | 80 | 40 | 40 | 40 | 20 | 20 | 80 |
| 367 | $2\times10^7$ | 80 | 80 | 5120 | 2560 | 1280 | 160 | 80 | 80 | 5120 |
| 375 | $2\times10^7$ | 640 | 640 | 2560 | 1280 | 1280 | 640 | 640 | 320 | 2560 |
| 378 | $2\times10^7$ | 2560 | 2560 | 2560 | 5120 | 2560 | 2560 | 1280 | 640 | 5120 |
| 389 | $2\times10^7$ | 40 | 40 | 1280 | 1280 | 1280 | 320 | 320 | 40 | 1280 |
| 399 | $2\times10^7$ | 160 | 160 | 160 | 160 | 160 | 320 | 160 | 160 | 320 |
| 405 | $2\times10^7$ | 5 | 5 | 160 | 1280 | 640 | 320 | 160 | 10 | 1280 |
| 418 | $2\times10^7$ | 20 | 40 | 80 | 40 | 20 | 20 | 20 | 20 | 80 |
| 501 | $2\times10^7$ | 320 | 320 | 5120 | 2560 | 2560 | 320 | 320 | 160 | 5120 |
| 503 | $2\times10^7$ | 160 | 160 | 160 | 160 | 160 | 320 | 160 | 80 | 320 |
| 520 | $2\times10^7$ | 1280 | 640 | 2560 | 2560 | 2560 | 1280 | 1280 | 1280 | 2560 |
| 546 | $2\times10^7$ | 160 | 320 | 320 | 320 | 160 | 160 | 320 | 160 | 320 |
| 568 | $2\times10^7$ | 20 | 40 | 640 | 5120 | 2560 | 2560 | 1280 | 160 | 5120 |
| 576 | $2\times10^7$ | 40 | 80 | 640 | 320 | 320 | 80 | 80 | 80 | 640 |
| 751 | $2\times10^7$ | 80 | 160 | 320 | 320 | 320 | 160 | 160 | 80 | 320 |
| 802 | $2\times10^7$ | 640 | 1280 | 10240 | 5120 | 2560 | | | 1280 | 10240 |
| 945 | $2\times10^7$ | 20 | 20 | 640 | 1280 | 640 | 640 | 160 | 160 | 1280 |
| 960 | $2\times10^7$ | 20 | 20 | 2560 | 1280 | 640 | 320 | 160 | 40 | 2560 |
| 1111 | $2\times10^7$ | 20 | 20 | 20 | 20 | 20 | 20 | 20 | 20 | 20 |

| Boost fold change to baseline Inaba | | | | | | | | | |
|---|---|---|---|---|---|---|---|---|---|
| Dose (CFU) | D1 | D4 | D7 | D15 | D29 | D57 | D85 | D180 | max FC |
| $2\times10^7$ | 1 | 2 | 4 | 4 | 2 | 2 | 1 | 1 | 4 |
| $2\times10^7$ | 1 | 1 | 4 | 2 | 2 | 2 | 1 | 1 | 4 |
| $2\times10^7$ | 1 | 1 | 64 | 32 | 16 | 2 | 1 | 1 | 64 |
| $2\times10^7$ | 1 | 1 | 4 | 2 | 2 | 1 | 1 | 0.5 | 4 |
| $2\times10^7$ | 1 | 1 | 1 | 2 | 1 | 1 | 0.5 | 0.25 | 2 |
| $2\times10^7$ | 1 | 1 | 32 | 32 | 32 | 8 | 8 | 1 | 32 |
| $2\times10^7$ | 1 | 1 | 1 | 1 | 1 | 2 | 1 | 1 | 2 |
| $2\times10^7$ | 1 | 1 | 32 | 256 | 128 | 64 | 32 | 2 | 256 |
| $2\times10^7$ | 1 | 2 | 4 | 2 | 1 | 1 | 1 | 1 | 4 |
| $2\times10^7$ | 1 | 1 | 16 | 8 | 8 | 1 | 1 | 0.5 | 16 |
| $2\times10^7$ | 1 | 1 | 1 | 1 | 1 | 2 | 1 | 0.5 | 2 |
| $2\times10^7$ | 1 | 0.5 | 2 | 2 | 2 | 1 | 1 | 1 | 2 |
| $2\times10^7$ | 1 | 2 | 2 | 2 | 1 | 1 | 2 | 1 | 2 |
| $2\times10^7$ | 1 | 2 | 32 | 256 | 128 | 128 | 64 | 8 | 256 |
| $2\times10^7$ | 1 | 2 | 16 | 8 | 8 | 2 | 2 | 2 | 16 |
| $2\times10^7$ | 1 | 2 | 4 | 4 | 4 | 2 | 2 | 1 | 4 |
| $2\times10^7$ | 1 | 2 | 16 | 8 | 4 | | | 2 | 16 |
| $2\times10^7$ | 1 | 1 | 32 | 64 | 32 | 32 | 8 | 8 | 64 |
| $2\times10^7$ | 1 | 1 | 128 | 64 | 32 | 16 | 8 | 2 | 128 |
| $2\times10^7$ | 1 | 1 | 1 | 1 | 1 | 1 | 1 | 1 | 1 |

| Primary immunization Inaba (VAT) | | | | | | | | | | |
|---|---|---|---|---|---|---|---|---|---|---|
| Dose interval (days) | Dose (CFU) | D1 | D4 | D7 | D15 | D29 | D57 | D85 | D180 | max |
| 322 | $10^7$ | 20 | 20 | 5120 | 20480 | 10240 | 5120 | 1280 | 320 | 20480 |
| 350 | $10^5$ | 5 | 5 | 1280 | 20480 | 10240 | 1280 | 320 | 40 | 20480 |
| 367 | $10^8$ | 5 | 5 | 160 | 5120 | 2560 | 640 | 320 | 320 | 5120 |
| 375 | $10^5$ | 5 | 5 | 160 | 5120 | 2560 | 2560 | 2560 | 1280 | 5120 |
| 378 | $10^8$ | 10 | 10 | 10240 | 20480 | 20480 | 5120 | 5120 | 2560 | 20480 |
| 389 | $10^8$ | 5 | 5 | 40 | 1280 | 320 | 160 | 80 | 80 | 1280 |
| 399 | $10^8$ | 5 | 5 | 5120 | 10240 | 5120 | 2560 | 1280 | 640 | 10240 |
| 405 | $10^8$ | 5 | 5 | 20 | 1280 | 160 | 20 | 10 | 5 | 1280 |
| 418 | $10^7$ | 5 | 5 | 1280 | 2560 | 1280 | 320 | 160 | 80 | 2560 |
| 501 | $10^8$ | 40 | 40 | 5120 | 20480 | 10240 | 5120 | 2560 | 2560 | 20480 |
| 503 | $10^7$ | 5 | 5 | 2560 | 10240 | 5120 | 2560 | 1280 | 320 | 10240 |
| 520 | $10^7$ | 5 | 5 | 1280 | 10240 | 10240 | 10240 | 10240 | 2560 | 10240 |
| 546 | $10^8$ | 5 | 5 | 320 | 2560 | 1280 | 2560 | 1280 | 640 | 2560 |
| 568 | $10^7$ | 5 | 5 | 160 | 2560 | 2560 | 1280 | 320 | 80 | 2560 |
| 576 | $10^7$ | 10 | 10 | 80 | 2560 | 1280 | 1280 | 640 | 320 | 2560 |
| 751 | $10^{10}$ | 40 | 80 | 5120 | 10240 | 5120 | 1280 | 640 | 640 | 10240 |
| 802 | $10^{10}$ | 640 | 640 | 10240 | 81920 | | | 5120 | 2560 | 81920 |
| 945 | $10^8$ | 5 | 5 | 20 | 1280 | 640 | 80 | 40 | 40 | 1280 |
| 960 | $10^8$ | 20 | 10 | 2560 | 10240 | 5120 | 1280 | 640 | 320 | 10240 |
| 1111 | $10^6$ | 5 | 5 | 160 | 1280 | 320 | 80 | 80 | 80 | 1280 |

| Primary immunization fold change to baseline Inaba | | | | | | | | | |
|---|---|---|---|---|---|---|---|---|---|
| Dose (CFU) | D1 | D4 | D7 | D15 | D29 | D57 | D85 | D180 | max FC |
| $10^7$ | 1 | 1 | 256 | 1024 | 512 | 256 | 64 | 16 | 1024 |
| $10^5$ | 1 | 1 | 256 | 4096 | 2048 | 256 | 64 | 8 | 4096 |
| $10^8$ | 1 | 1 | 32 | 1024 | 512 | 128 | 64 | 64 | 1024 |
| $10^5$ | 1 | 1 | 32 | 1024 | 512 | 512 | 512 | 256 | 1024 |
| $10^8$ | 1 | 1 | 1024 | 2048 | 2048 | 512 | 512 | 256 | 2048 |
| $10^8$ | 1 | 1 | 8 | 256 | 64 | 32 | 16 | 16 | 256 |
| $10^8$ | 1 | 1 | 1024 | 2048 | 1024 | 512 | 256 | 128 | 2048 |
| $10^8$ | 1 | 1 | 4 | 256 | 32 | 4 | 2 | 1 | 256 |
| $10^7$ | 1 | 1 | 256 | 512 | 256 | 64 | 32 | 16 | 512 |
| $10^8$ | 1 | 1 | 128 | 512 | 256 | 128 | 64 | 64 | 512 |
| $10^7$ | 1 | 1 | 512 | 2048 | 1024 | 512 | 256 | 64 | 2048 |
| $10^7$ | 1 | 1 | 256 | 2048 | 2048 | 2048 | 2048 | 512 | 2048 |
| $10^8$ | 1 | 1 | 64 | 512 | 256 | 512 | 256 | 128 | 512 |
| $10^7$ | 1 | 1 | 32 | 512 | 512 | 256 | 64 | 16 | 512 |
| $10^7$ | 1 | 1 | 8 | 256 | 128 | 128 | 64 | 32 | 256 |
| $10^{10}$ | 1 | 2 | 128 | 256 | 128 | 32 | 16 | 16 | 256 |
| $10^{10}$ | 1 | 1 | 16 | 128 | | | 8 | 4 | 128 |
| $10^8$ | 1 | 1 | 4 | 256 | 128 | 16 | 8 | 8 | 256 |
| $10^8$ | 1 | 0.5 | 128 | 512 | 256 | 64 | 32 | 16 | 512 |
| $10^6$ | 1 | 1 | 32 | 256 | 64 | 16 | 16 | 16 | 256 |

(blue) no serum sample

| Boost Ogawa (VAT) | | | | | | | | | | |
|---|---|---|---|---|---|---|---|---|---|---|
| Dose interval (days) | Dose (CFU) | D1 | D4 | D7 | D15 | D29 | D57 | D85 | D180 | max |
| 322 | $2\times10^7$ | 5 | 5 | 80 | 40 | 10 | 5 | 5 | 5 | 80 |
| 350 | $2\times10^7$ | 5 | 5 | 40 | 40 | 20 | 10 | 10 | 10 | 40 |
| 367 | $2\times10^7$ | 80 | 80 | 2560 | 1280 | 640 | 160 | 40 | 80 | 2560 |
| 375 | $2\times10^7$ | 5 | 5 | 640 | 320 | 160 | 40 | 20 | 10 | 640 |
| 378 | $2\times10^7$ | 2560 | 1280 | 2560 | 5120 | 2560 | 2560 | 1280 | 1280 | 5120 |
| 389 | $2\times10^7$ | 80 | 80 | 640 | 640 | 640 | 160 | 320 | 320 | 640 |
| 399 | $2\times10^7$ | 160 | 320 | 160 | 160 | 160 | 320 | 160 | 320 | 320 |
| 405 | $2\times10^7$ | 5 | 5 | 160 | 1280 | 640 | 160 | 40 | 10 | 1280 |
| 418 | $2\times10^7$ | 5 | 5 | 40 | 20 | 10 | 10 | 10 | 10 | 40 |
| 501 | $2\times10^7$ | 40 | 40 | 2560 | 2560 | 1280 | 320 | 80 | 20 | 2560 |
| 503 | $2\times10^7$ | 80 | 80 | 40 | 80 | 80 | 80 | 80 | 40 | 80 |
| 520 | $2\times10^7$ | 160 | 160 | 1280 | 1280 | 320 | 160 | 160 | 80 | 1280 |
| 546 | $2\times10^7$ | 640 | 640 | 1280 | 640 | 640 | 640 | 1280 | 640 | 1280 |
| 568 | $2\times10^7$ | 320 | 640 | 1280 | 2560 | 2560 | 1280 | 320 | 320 | 2560 |
| 576 | $2\times10^7$ | 80 | 80 | 640 | 320 | 320 | 160 | 80 | 80 | 640 |
| 751 | $2\times10^7$ | 80 | 80 | 640 | 320 | 320 | 160 | 80 | 40 | 640 |
| 802 | $2\times10^7$ | 320 | 160 | 5120 | 2560 | 1280 | | | 320 | 5120 |
| 945 | $2\times10^7$ | 320 | 640 | 640 | 1280 | 640 | 320 | 320 | 320 | 1280 |
| 960 | $2\times10^7$ | 10 | 10 | 1280 | 640 | 640 | 160 | 40 | 20 | 1280 |
| 1111 | $2\times10^7$ | 40 | 40 | 80 | 40 | 80 | 40 | 80 | 40 | 80 |

| Boost fold change to baseline Ogawa | | | | | | | | | |
|---|---|---|---|---|---|---|---|---|---|
| Dose (CFU) | D1 | D4 | D7 | D15 | D29 | D57 | D85 | D180 | max FC |
| $2\times10^7$ | 1 | 1 | 16 | 8 | 2 | 1 | 1 | 1 | 16 |
| $2\times10^7$ | 1 | 1 | 8 | 8 | 4 | 2 | 2 | 2 | 8 |
| $2\times10^7$ | 1 | 1 | 32 | 16 | 8 | 2 | 0.5 | 1 | 32 |
| $2\times10^7$ | 1 | 1 | 128 | 64 | 32 | 8 | 4 | 2 | 128 |
| $2\times10^7$ | 1 | 0.5 | 1 | 2 | 1 | 1 | 0.5 | 0.5 | 2 |
| $2\times10^7$ | 1 | 1 | 8 | 8 | 8 | 2 | 4 | 4 | 8 |
| $2\times10^7$ | 1 | 2 | 1 | 1 | 1 | 2 | 1 | 2 | 2 |
| $2\times10^7$ | 1 | 1 | 32 | 256 | 128 | 32 | 8 | 2 | 256 |
| $2\times10^7$ | 1 | 1 | 8 | 4 | 2 | 2 | 2 | 2 | 8 |
| $2\times10^7$ | 1 | 1 | 64 | 64 | 32 | 8 | 2 | 0.5 | 64 |
| $2\times10^7$ | 1 | 1 | 0.5 | 1 | 1 | 1 | 1 | 0.5 | 1 |
| $2\times10^7$ | 1 | 1 | 8 | 8 | 2 | 1 | 1 | 0.5 | 8 |
| $2\times10^7$ | 1 | 1 | 2 | 1 | 1 | 1 | 2 | 1 | 2 |
| $2\times10^7$ | 1 | 2 | 4 | 8 | 8 | 4 | 1 | 1 | 8 |
| $2\times10^7$ | 1 | 1 | 8 | 4 | 4 | 2 | 1 | 1 | 8 |
| $2\times10^7$ | 1 | 1 | 8 | 4 | 4 | 2 | 1 | 0.5 | 8 |
| $2\times10^7$ | 1 | 0.5 | 16 | 8 | 4 | | | 1 | 16 |
| $2\times10^7$ | 1 | 2 | 2 | 4 | 2 | 1 | 1 | 1 | 4 |
| $2\times10^7$ | 1 | 1 | 128 | 64 | 64 | 16 | 4 | 2 | 128 |
| $2\times10^7$ | 1 | 1 | 2 | 1 | 2 | 1 | 2 | 1 | 2 |

| Primary immunization Ogawa (VAT) | | | | | | | | | | |
|---|---|---|---|---|---|---|---|---|---|---|
| Dose interval (days) | Dose (CFU) | D1 | D4 | D7 | D15 | D29 | D57 | D85 | D180 | max |
| 322 | $10^7$ | 5 | 5 | 160 | 640 | 160 | 40 | 10 | 5 | 640 |
| 350 | $10^5$ | 5 | 5 | 320 | 10240 | 2560 | 320 | 80 | 10 | 10240 |
| 367 | $10^8$ | 5 | 5 | 320 | 5120 | 5120 | 640 | 320 | 320 | 5120 |
| 375 | $10^5$ | 5 | 5 | 10 | 40 | 20 | 20 | 20 | 10 | 40 |
| 378 | $10^8$ | 320 | 160 | 5120 | 20480 | 20480 | 10240 | 5120 | 2560 | 20480 |
| 389 | $10^8$ | 5 | 5 | 40 | 640 | 160 | 80 | 80 | 80 | 640 |
| 399 | $10^8$ | 5 | 5 | 2560 | 10240 | 2560 | 1280 | 1280 | 640 | 10240 |
| 405 | $10^8$ | 5 | 5 | 20 | 640 | 80 | 10 | 5 | 5 | 640 |
| 418 | $10^7$ | 5 | 5 | 320 | 1280 | 320 | 40 | 20 | 10 | 1280 |
| 501 | $10^8$ | 5 | 5 | 320 | 2560 | 1280 | 320 | 160 | 40 | 2560 |
| 503 | $10^7$ | 5 | 5 | 1280 | 5120 | 2560 | 640 | 320 | 80 | 5120 |
| 520 | $10^7$ | 5 | 5 | 80 | 640 | 320 | 320 | 320 | 320 | 640 |
| 546 | $10^8$ | 160 | 160 | 640 | 1280 | 1280 | 640 | 640 | 1280 | 1280 |
| 568 | $10^7$ | 80 | 80 | 640 | 10240 | 5120 | 5120 | 2560 | 640 | 10240 |
| 576 | $10^7$ | 20 | 40 | 80 | 1280 | 2560 | 1280 | 640 | 160 | 2560 |
| 751 | $10^{10}$ | 40 | 80 | 10240 | 10240 | 2560 | 2560 | 640 | 640 | 10240 |
| 802 | $10^{10}$ | 320 | 320 | 2560 | 40960 | | | 2560 | 1280 | 40960 |
| 945 | $10^8$ | 80 | 80 | 160 | 5120 | 1280 | 640 | 320 | 320 | 5120 |
| 960 | $10^8$ | 5 | 5 | 160 | 5120 | 1280 | 640 | 640 | 320 | 5120 |
| 1111 | $10^6$ | 20 | 20 | 80 | 320 | 160 | 80 | 40 | 80 | 320 |

| Primary immunization fold change to baseline Ogawa | | | | | | | | | |
|---|---|---|---|---|---|---|---|---|---|
| Dose (CFU) | D1 | D4 | D7 | D15 | D29 | D57 | D85 | D180 | max FC |
| $10^7$ | 1 | 1 | 32 | 128 | 32 | 8 | 2 | 1 | 128 |
| $10^5$ | 1 | 1 | 64 | 2048 | 512 | 64 | 16 | 2 | 2048 |
| $10^8$ | 1 | 1 | 64 | 1024 | 1024 | 128 | 64 | 64 | 1024 |
| $10^5$ | 1 | 1 | 2 | 8 | 4 | 4 | 4 | 2 | 8 |
| $10^8$ | 1 | 0.5 | 16 | 64 | 64 | 32 | 16 | 8 | 64 |
| $10^8$ | 1 | 1 | 8 | 128 | 32 | 16 | 16 | 16 | 128 |
| $10^8$ | 1 | 1 | 512 | 2048 | 512 | 256 | 256 | 128 | 2048 |
| $10^8$ | 1 | 1 | 4 | 128 | 16 | 2 | 1 | 1 | 128 |
| $10^7$ | 1 | 1 | 64 | 256 | 64 | 8 | 4 | 2 | 256 |
| $10^8$ | 1 | 1 | 64 | 512 | 256 | 64 | 32 | 8 | 512 |
| $10^7$ | 1 | 1 | 256 | 1024 | 512 | 128 | 64 | 16 | 1024 |
| $10^7$ | 1 | 1 | 16 | 128 | 64 | 64 | 64 | 64 | 128 |
| $10^8$ | 1 | 1 | 4 | 8 | 8 | 4 | 4 | 8 | 8 |
| $10^7$ | 1 | 1 | 8 | 128 | 64 | 64 | 32 | 8 | 128 |
| $10^7$ | 1 | 2 | 4 | 64 | 128 | 64 | 32 | 8 | 128 |
| $10^{10}$ | 1 | 2 | 256 | 256 | 64 | 64 | 16 | 16 | 256 |
| $10^{10}$ | 1 | 1 | 8 | 128 | | | 8 | 4 | 128 |
| $10^8$ | 1 | 1 | 2 | 64 | 16 | 8 | 4 | 4 | 64 |
| $10^8$ | 1 | 1 | 32 | 1024 | 256 | 128 | 128 | 64 | 1024 |
| $10^6$ | 1 | 1 | 4 | 16 | 8 | 4 | 2 | 4 | 16 |

**Table S4: Kinetics of mean vibriocidal titers to both serotypes in participants following primary and booster vaccination.** Geometric mean titers (GMT) with 95% confidence intervals (CI) are presented at baseline and day 7, 15, 29 and 180 post-vaccination.

| Serotype | Dose interval | n | GMT (95% CI) on day of vaccination | | | | |
|---|---|---|---|---|---|---|---|
| | | | Baseline | 7 | 15 | 29 | 180 |
| Inaba | Primary | 20 | 9·7 (5·5-17) | 663 (250-1753) | 6089 (3493-10614) | 2560 (1325-4947) | 288 (131-635) |
| | Boost | 20 | 89 (41-193) | 640 (295-1387) | 640 (291-1407) | 453 (216-949) | 106 (56-197) |
| Ogawa | Primary | 20 | 15 (7·3-31) | 299 (128-697) | 2473 (1137-5377) | 889 (388-2035) | 126 (50-315) |
| | Boost | 20 | 59 (25-139) | 469 (227-968) | 422 (196-911) | 279 (129-602) | 65 (30-140) |

**Table S5: Isotype- and antigen-specific immune responses in sera after primary and booster vaccination.**
Blue: no serum sample; light grey: below limit of detection.

| Boost Inaba OSP IgM (RAU) | | | | | | | | | | |
|---|---|---|---|---|---|---|---|---|---|---|
| Dose interval (days) | Dose (CFU) | D1 | D4 | D7 | D15 | D29 | D57 | D85 | D180 | max |
| 389 | $2\times10^7$ | 0·00389 | 0·00422 | 0·03847 | 0·02040 | 0·00333 | 0·00621 | 0·00710 | 0·00709 | 0·03847 |
| 405 | $2\times10^7$ | 0·00026 | 0·00014 | 0·00650 | 0·03889 | 0·01440 | 0·00365 | 0·00174 | 0·00119 | 0·03889 |
| 322 | $2\times10^7$ | 0·00583 | 0·00599 | 0·01618 | 0·01345 | 0·01000 | 0·00579 | 0·01356 | 0·01418 | 0·01618 |
| 418 | $2\times10^7$ | 0·00243 | 0·00229 | 0·00584 | 0·00434 | 0·00285 | 0·00270 | 0·00326 | 0·00179 | 0·00584 |
| 378 | $2\times10^7$ | 0·03662 | 0·03696 | 0·07753 | 0·19548 | 0·11976 | 0·08124 | 0·04969 | 0·02445 | 0·19548 |
| 399 | $2\times10^7$ | 0·01426 | 0·01398 | 0·01272 | 0·01298 | 0·01374 | 0·02216 | 0·01588 | 0·01721 | 0·02216 |
| 568 | $2\times10^7$ | 0·00412 | 0·00363 | 0·01893 | 0·11267 | 0·05925 | 0·03773 | 0·01464 | 0·00506 | 0·11267 |
| 367 | $2\times10^7$ | 0·00466 | 0·00758 | 0·10136 | 0·06557 | 0·02798 | 0·01534 | 0·00849 | 0·00776 | 0·10136 |
| 503 | $2\times10^7$ | 0·00810 | 0·00808 | 0·00726 | 0·00772 | 0·00868 | 0·01179 | 0·01122 | 0·00815 | 0·01179 |
| 501 | $2\times10^7$ | 0·00604 | 0·00585 | 0·10726 | 0·08039 | 0·04560 | 0·02026 | 0·01125 | 0·00720 | 0·10726 |
| 751 | $2\times10^7$ | 0·00667 | 0·00879 | 0·02517 | 0·02642 | 0·01277 | 0·00816 | 0·00916 | 0·00771 | 0·02642 |
| 546 | $2\times10^7$ | 0·01034 | 0·01026 | 0·01252 | 0·01084 | 0·00876 | 0·00935 | 0·00880 | 0·00781 | 0·01252 |
| 945 | $2\times10^7$ | 0·00254 | 0·00222 | 0·04493 | 0·04629 | 0·01576 | 0·00462 | 0·00327 | 0·00110 | 0·04629 |
| 802 | $2\times10^7$ | 0·02588 | 0·02764 | 0·15590 | 0·13644 | 0·09735 | | | 0·01592 | 0·15590 |
| 960 | $2\times10^7$ | 0·00647 | 0·00644 | 0·14477 | 0·11953 | 0·06015 | 0·01432 | 0·01048 | 0·01348 | 0·14477 |
| 375 | $2\times10^7$ | 0·00714 | 0·00669 | 0·03819 | 0·02341 | 0·01405 | 0·00976 | 0·00775 | 0·00546 | 0·03819 |
| 350 | $2\times10^7$ | 0·02166 | 0·02095 | 0·02529 | 0·01982 | 0·02018 | 0·02112 | 0·01952 | 0·02704 | 0·02704 |
| 1111 | $2\times10^7$ | 0·00873 | 0·00937 | 0·00901 | 0·00847 | 0·00839 | 0·00776 | 0·00989 | 0·00588 | 0·00989 |
| 576 | $2\times10^7$ | 0·01447 | 0·01452 | 0·07295 | 0·04878 | 0·03853 | 0·02489 | 0·01518 | 0·00969 | 0·07295 |
| 520 | $2\times10^7$ | 0·01327 | 0·01619 | 0·13504 | 0·09325 | 0·05363 | 0·03185 | 0·01750 | 0·01057 | 0·13504 |

| Boost fold change Inaba OSP IgM | | | | | | | | | |
|---|---|---|---|---|---|---|---|---|---|
| Dose (CFU) | D1 | D4 | D7 | D15 | D29 | D57 | D85 | D180 | max FC |
| $2\times10^7$ | 1·00 | 1·08 | 9·90 | 5·25 | 0·86 | 1·60 | 1·83 | 1·83 | 9·90 |
| $2\times10^7$ | 1·00 | 0·55 | 25·34 | 151·56 | 56·12 | 14·23 | 6·77 | 4·64 | 151·56 |
| $2\times10^7$ | 1·00 | 1·03 | 2·78 | 2·31 | 1·71 | 0·99 | 2·33 | 2·43 | 2·78 |
| $2\times10^7$ | 1·00 | 0·94 | 2·40 | 1·79 | 1·17 | 1·11 | 1·34 | 0·74 | 2·40 |
| $2\times10^7$ | 1·00 | 1·01 | 2·12 | 5·34 | 3·27 | 2·22 | 1·36 | 0·67 | 5·34 |
| $2\times10^7$ | 1·00 | 0·98 | 0·89 | 0·91 | 0·96 | 1·55 | 1·11 | 1·21 | 1·55 |
| $2\times10^7$ | 1·00 | 0·88 | 4·60 | 27·36 | 14·39 | 9·16 | 3·56 | 1·23 | 27·36 |
| $2\times10^7$ | 1·00 | 1·63 | 21·77 | 14·08 | 6·01 | 3·29 | 1·82 | 1·67 | 21·77 |
| $2\times10^7$ | 1·00 | 1·00 | 0·90 | 0·95 | 1·07 | 1·46 | 1·39 | 1·01 | 1·46 |
| $2\times10^7$ | 1·00 | 0·97 | 17·77 | 13·32 | 7·55 | 3·36 | 1·86 | 1·19 | 17·77 |
| $2\times10^7$ | 1·00 | 1·32 | 3·78 | 3·96 | 1·92 | 1·22 | 1·37 | 1·16 | 3·96 |
| $2\times10^7$ | 1·00 | 0·99 | 1·21 | 1·05 | 0·85 | 0·90 | 0·85 | 0·75 | 1·21 |
| $2\times10^7$ | 1·00 | 0·87 | 17·68 | 18·22 | 6·20 | 1·82 | 1·29 | 0·43 | 18·22 |
| $2\times10^7$ | 1·00 | 1·07 | 6·02 | 5·27 | 3·76 | | | 0·62 | 6·02 |
| $2\times10^7$ | 1·00 | 0·99 | 22·36 | 18·46 | 9·29 | 2·21 | 1·62 | 2·08 | 22·36 |
| $2\times10^7$ | 1·00 | 0·94 | 5·35 | 3·28 | 1·97 | 1·37 | 1·09 | 0·77 | 5·35 |
| $2\times10^7$ | 1·00 | 0·97 | 1·17 | 0·92 | 0·93 | 0·97 | 0·90 | 1·25 | 1·25 |
| $2\times10^7$ | 1·00 | 1·07 | 1·03 | 0·97 | 0·96 | 0·89 | 1·13 | 0·67 | 1·13 |
| $2\times10^7$ | 1·00 | 1·00 | 5·04 | 3·37 | 2·66 | 1·72 | 1·05 | 0·67 | 5·04 |
| $2\times10^7$ | 1·00 | 1·22 | 10·18 | 7·03 | 4·04 | 2·40 | 1·32 | 0·80 | 10·18 |

| Primary Inaba OSP IgM (RAU) | | | | | | | | | | |
|---|---|---|---|---|---|---|---|---|---|---|
| Dose interval (days) | Dose (CFU) | D1 | D4 | D7 | D15 | D29 | D57 | D85 | D180 | max |
| 389 | $10^8$ | 0·02210 | 0·02278 | 0·02289 | 0·11330 | 0·02619 | 0·00940 | 0·00640 | 0·00484 | 0·11330 |
| 405 | $10^8$ | 0·00196 | 0·00151 | 0·00151 | 0·03434 | 0·01100 | 0·00211 | 0·00145 | 0·00043 | 0·03434 |
| 322 | $10^7$ | 0·00332 | 0·00415 | 0·18596 | 3·16258 | 1·40009 | 0·28981 | 0·06714 | 0·00838 | 3·16258 |
| 418 | $10^7$ | 0·00456 | 0·00541 | 0·03353 | 0·54943 | 0·28543 | 0·02710 | 0·01302 | 0·00696 | 0·54943 |
| 378 | $10^8$ | 0·00143 | 0·00166 | 0·20431 | 3·97839 | 2·31500 | 0·77571 | 0·37617 | 0·17124 | 3·97839 |
| 399 | $10^8$ | 0·00449 | 0·00388 | 0·13228 | 3·00340 | 1·02997 | 0·21627 | 0·14575 | 0·07240 | 3·00340 |
| 568 | $10^7$ | 0·00400 | 0·00392 | 0·00891 | 1·10863 | 0·82969 | 0·28677 | 0·12419 | 0·02575 | 1·10863 |
| 367 | $10^8$ | 0·00100 | 0·00112 | 0·00353 | 0·30476 | 0·18180 | 0·05164 | 0·02710 | 0·01474 | 0·30476 |
| 503 | $10^7$ | 0·01156 | 0·00962 | 0·02580 | 0·78914 | 0·33725 | 0·08086 | 0·05265 | 0·01212 | 0·78914 |
| 501 | $10^8$ | 0·01293 | 0·01374 | 0·14948 | 3·30181 | 0·96768 | 0·39240 | 0·10271 | 0·03311 | 3·30181 |
| 751 | $10^{10}$ | 0·00426 | 0·00453 | 0·33315 | 0·94811 | 0·26635 | 0·11948 | 0·03265 | 0·01855 | 0·94811 |
| 546 | $10^8$ | 0·02113 | 0·02248 | 0·02040 | 0·06589 | 0·08768 | 0·05750 | 0·03167 | 0·01348 | 0·08768 |
| 945 | $10^8$ | 0·00089 | 0·00076 | 0·00075 | 0·02221 | 0·01372 | 0·00391 | 0·00228 | 0·00177 | 0·02221 |
| 802 | $10^{10}$ | 0·01880 | 0·02335 | 0·30780 | 6·79807 | | | 0·46855 | 0·10156 | 6·79807 |
| 960 | $10^8$ | 0·01379 | 0·01208 | 0·01937 | 0·18950 | 0·05626 | 0·03126 | 0·00977 | 0·00854 | 0·18950 |
| 375 | $10^5$ | 0·00126 | 0·00095 | 0·00162 | 0·01764 | 0·01516 | 0·00761 | 0·01024 | 0·00612 | 0·01764 |
| 350 | $10^5$ | 0·01793 | 0·01833 | 0·03245 | 0·31615 | 0·14471 | 0·03311 | 0·01909 | 0·01317 | 0·31615 |
| 1111 | $10^6$ | 0·00561 | 0·00682 | 0·00965 | 0·01408 | 0·00768 | 0·00737 | 0·00640 | 0·00437 | 0·01408 |
| 576 | $10^7$ | 0·00975 | 0·01157 | 0·02102 | 1·71489 | 1·01604 | 0·45261 | 0·24544 | 0·05908 | 1·71489 |
| 520 | $10^7$ | 0·00272 | 0·00239 | 0·00892 | 0·20537 | 0·21348 | 0·15186 | 0·11463 | 0·06743 | 0·21348 |

| Primary fold change Inaba OSP IgM | | | | | | | | | |
|---|---|---|---|---|---|---|---|---|---|
| Dose (CFU) | D1 | D4 | D7 | D15 | D29 | D57 | D85 | D180 | max FC |
| $10^8$ | 1·00 | 1·03 | 1·04 | 5·13 | 1·19 | 0·43 | 0·29 | 0·22 | 5·13 |
| $10^8$ | 1·00 | 0·77 | 0·77 | 17·52 | 5·61 | 1·08 | 0·74 | 0·22 | 17·52 |
| $10^7$ | 1·00 | 1·25 | 56·01 | 952·58 | 421·71 | 87·29 | 20·22 | 2·52 | 952·58 |
| $10^7$ | 1·00 | 1·19 | 7·35 | 120·49 | 62·59 | 5·94 | 2·86 | 1·53 | 120·49 |
| $10^8$ | 1·00 | 1·16 | 142·87 | 2782·09 | 1618·88 | 542·45 | 263·06 | 119·75 | 2782·09 |
| $10^8$ | 1·00 | 0·86 | 29·46 | 668·91 | 229·39 | 48·17 | 32·46 | 16·12 | 668·91 |
| $10^7$ | 1·00 | 0·98 | 2·23 | 277·16 | 207·42 | 71·69 | 31·05 | 6·44 | 277·16 |
| $10^8$ | 1·00 | 1·12 | 3·53 | 304·76 | 181·80 | 51·64 | 27·10 | 14·74 | 304·76 |
| $10^7$ | 1·00 | 0·83 | 2·23 | 68·26 | 29·17 | 6·99 | 4·55 | 1·05 | 68·26 |
| $10^8$ | 1·00 | 1·06 | 11·56 | 255·36 | 74·84 | 30·35 | 7·94 | 2·56 | 255·36 |
| $10^{10}$ | 1·00 | 1·06 | 78·20 | 222·56 | 62·52 | 28·05 | 7·66 | 4·35 | 222·56 |
| $10^8$ | 1·00 | 1·06 | 0·97 | 3·12 | 4·15 | 2·72 | 1·50 | 0·64 | 4·15 |
| $10^8$ | 1·00 | 0·85 | 0·84 | 24·96 | 15·42 | 4·39 | 2·56 | 1·99 | 24·96 |
| $10^{10}$ | 1·00 | 1·24 | 16·37 | 361·60 | | | 24·92 | 5·40 | 361·60 |
| $10^8$ | 1·00 | 0·88 | 1·40 | 13·74 | 4·08 | 2·27 | 0·71 | 0·62 | 13·74 |
| $10^5$ | 1·00 | 0·75 | 1·29 | 14·00 | 12·03 | 6·04 | 8·13 | 4·86 | 14·00 |
| $10^5$ | 1·00 | 1·02 | 1·81 | 17·63 | 8·07 | 1·85 | 1·06 | 0·73 | 17·63 |
| $10^6$ | 1·00 | 1·22 | 1·72 | 2·51 | 1·37 | 1·31 | 1·14 | 0·78 | 2·51 |
| $10^7$ | 1·00 | 1·19 | 2·16 | 175·89 | 104·21 | 46·42 | 25·17 | 6·06 | 175·89 |
| $10^7$ | 1·00 | 0·88 | 3·28 | 75·50 | 78·49 | 55·83 | 42·14 | 24·79 | 78·49 |

no serum sample

| Boost Inaba OSP IgG (RAU) | | | | | | | | | | |
|---|---|---|---|---|---|---|---|---|---|---|
| Dose interval (days) | Dose (CFU) | D1 | D4 | D7 | D15 | D29 | D57 | D85 | D180 | max |
| 389 | $2\times10^{7}$ | 0·09484 | 0·07074 | 5·46950 | 39·37130 | 1·11013 | 6·98312 | 0·14295 | 0·05241 | 39·37130 |
| 405 | $2\times10^{7}$ | 0·00133 | 0·00094 | 0·01223 | 0·99583 | 0·26452 | 0·07732 | 0·01854 | 0·00169 | 0·99583 |
| 322 | $2\times10^{7}$ | 0·00510 | 0·00453 | 0·01496 | 0·03048 | 0·02538 | 0·01411 | 0·00769 | 0·00420 | 0·03048 |
| 418 | $2\times10^{7}$ | 0·06830 | 0·05944 | 0·08725 | 0·28744 | 0·24195 | 0·23428 | 0·19256 | 0·09101 | 0·28744 |
| 378 | $2\times10^{7}$ | 0·00989 | 0·01032 | 0·05017 | 0·29322 | 0·13823 | 0·06612 | 0·03237 | 0·01979 | 0·29322 |
| 399 | $2\times10^{7}$ | 0·03290 | 0·02760 | 0·02677 | 0·02806 | 0·02612 | 0·03321 | 0·02940 | 0·02811 | 0·03321 |
| 568 | $2\times10^{7}$ | 0·02110 | 0·01919 | 0·98121 | 88·43447 | 4·04981 | 3·36243 | 11·36431 | 0·11010 | 88·43447 |
| 367 | $2\times10^{7}$ | 0·00805 | 0·00993 | 7·05891 | 3·20888 | 0·61704 | 0·18287 | 0·05414 | 0·01460 | 7·05891 |
| 503 | $2\times10^{7}$ | 0·07187 | 0·07932 | 0·05463 | 0·07202 | 0·06458 | 0·06550 | 0·06932 | 0·19260 | 0·19260 |
| 501 | $2\times10^{7}$ | 0·05652 | 0·06048 | 0·13183 | 0·52084 | 0·54387 | 0·18645 | 0·15037 | 0·32238 | 0·54387 |
| 751 | $2\times10^{7}$ | 0·08828 | 0·09706 | 0·19705 | 0·53778 | 0·33842 | 0·17892 | 0·20319 | 0·21489 | 0·53778 |
| 546 | $2\times10^{7}$ | 0·03583 | 0·02742 | 0·06945 | 0·07310 | 0·07215 | 0·05146 | 0·05251 | 0·09847 | 0·09847 |
| 945 | $2\times10^{7}$ | 0·08298 | 0·07606 | 0·98195 | 3·73892 | 29·90304 | 4·67535 | 2·34500 | 1·49878 | 29·90304 |
| 802 | $2\times10^{7}$ | 0·02711 | 0·02732 | 15·38269 | 4·52850 | 2·67965 | | | 0·47751 | 15·38269 |
| 960 | $2\times10^{7}$ | 0·01580 | 0·01411 | 0·78760 | 1·76406 | 0·71601 | 0·24334 | 0·13769 | 0·32973 | 1·76406 |
| 375 | $2\times10^{7}$ | 0·01066 | 0·01022 | 0·14070 | 0·11466 | 0·08387 | 0·05952 | 0·03950 | 0·08171 | 0·14070 |
| 350 | $2\times10^{7}$ | 0·07080 | 0·05541 | 0·41937 | 0·51234 | 0·27070 | 0·13919 | 0·09696 | 0·47739 | 0·51234 |
| 1111 | $2\times10^{7}$ | 0·00402 | 0·00512 | 0·00457 | 0·00852 | 0·01006 | 0·01129 | 0·02251 | 0·02174 | 0·02251 |
| 576 | $2\times10^{7}$ | 0·00120 | 0·00126 | 0·02613 | 0·06616 | 0·05669 | 0·02882 | 0·00684 | 0·00373 | 0·06616 |
| 520 | $2\times10^{7}$ | 0·03161 | 0·03748 | 0·50867 | 0·35426 | 0·22370 | 0·12055 | 0·18825 | 0·11858 | 0·50867 |

| Boost fold change Inaba OSP IgG | | | | | | | | | |
|---|---|---|---|---|---|---|---|---|---|
| Dose (CFU) | D1 | D4 | D7 | D15 | D29 | D57 | D85 | D180 | max FC |
| $2\times10^{7}$ | 1·00 | 0·75 | 57·67 | 415·11 | 11·70 | 73·63 | 1·51 | 0·55 | 415·11 |
| $2\times10^{7}$ | 1·00 | 0·71 | 9·18 | 747·35 | 198·52 | 58·03 | 13·91 | 1·27 | 747·35 |
| $2\times10^{7}$ | 1·00 | 0·89 | 2·93 | 5·97 | 4·97 | 2·76 | 1·51 | 0·82 | 5·97 |
| $2\times10^{7}$ | 1·00 | 0·87 | 1·28 | 4·21 | 3·54 | 3·43 | 2·82 | 1·33 | 4·21 |
| $2\times10^{7}$ | 1·00 | 1·04 | 5·08 | 29·66 | 13·98 | 6·69 | 3·27 | 2·00 | 29·66 |
| $2\times10^{7}$ | 1·00 | 0·84 | 0·81 | 0·85 | 0·79 | 1·01 | 0·89 | 0·85 | 1·01 |
| $2\times10^{7}$ | 1·00 | 0·91 | 46·50 | 4190·66 | 191·91 | 159·34 | 538·52 | 5·22 | 4190·66 |
| $2\times10^{7}$ | 1·00 | 1·23 | 877·21 | 398·77 | 76·68 | 22·73 | 6·73 | 1·81 | 877·21 |
| $2\times10^{7}$ | 1·00 | 1·10 | 0·76 | 1·00 | 0·90 | 0·91 | 0·96 | 2·68 | 2·68 |
| $2\times10^{7}$ | 1·00 | 1·07 | 2·33 | 9·21 | 9·62 | 3·30 | 2·66 | 5·70 | 9·62 |
| $2\times10^{7}$ | 1·00 | 1·10 | 2·23 | 6·09 | 3·83 | 2·03 | 2·30 | 2·43 | 6·09 |
| $2\times10^{7}$ | 1·00 | 0·77 | 1·94 | 2·04 | 2·01 | 1·44 | 1·47 | 2·75 | 2·75 |
| $2\times10^{7}$ | 1·00 | 0·92 | 11·83 | 45·06 | 360·38 | 56·35 | 28·26 | 18·06 | 360·38 |
| $2\times10^{7}$ | 1·00 | 1·01 | 567·38 | 167·03 | 98·84 | | | 17·61 | 567·38 |
| $2\times10^{7}$ | 1·00 | 0·89 | 49·85 | 111·65 | 45·32 | 15·40 | 8·71 | 20·87 | 111·65 |
| $2\times10^{7}$ | 1·00 | 0·96 | 13·20 | 10·75 | 7·87 | 5·58 | 3·71 | 7·66 | 13·20 |
| $2\times10^{7}$ | 1·00 | 0·78 | 5·92 | 7·24 | 3·82 | 1·97 | 1·37 | 6·74 | 7·24 |
| $2\times10^{7}$ | 1·00 | 1·27 | 1·14 | 2·12 | 2·50 | 2·81 | 5·60 | 5·41 | 5·60 |
| $2\times10^{7}$ | 1·00 | 1·06 | 21·85 | 55·31 | 47·40 | 24·10 | 5·72 | 3·12 | 55·31 |
| $2\times10^{7}$ | 1·00 | 1·19 | 16·09 | 11·21 | 7·08 | 3·81 | 5·96 | 3·75 | 16·09 |

| Primary Inaba OSP IgG (RAU) | | | | | | | | | | |
|---|---|---|---|---|---|---|---|---|---|---|
| Dose interval (days) | Dose (CFU) | D1 | D4 | D7 | D15 | D29 | D57 | D85 | D180 | max |
| 389 | $10^{8}$ | 0·00772 | 0·00852 | 0·00840 | 0·00931 | 0·00201 | 0·01163 | 0·02465 | 0·03705 | 0·03705 |
| 405 | $10^{8}$ | 0·00151 | 0·00247 | 0·00254 | 0·00125 | 0·00007 | | | 0·00004 | 0·00254 |
| 322 | $10^{7}$ | 0·00192 | 0·00231 | 0·00167 | 0·00257 | 0·00299 | 0·00279 | 0·00230 | 0·00249 | 0·00299 |
| 418 | $10^{7}$ | 0·01160 | 0·00981 | 0·00961 | 0·01058 | 0·00981 | 0·02887 | 0·03418 | 0·08353 | 0·08353 |
| 378 | $10^{8}$ | 0·00382 | 0·00457 | 0·00319 | 0·00405 | 0·00518 | 0·00441 | 0·00380 | 0·00448 | 0·00518 |
| 399 | $10^{8}$ | 0·00675 | 0·00481 | 0·00747 | 0·00678 | 0·00381 | 0·00319 | 0·00474 | 0·02701 | 0·02701 |
| 568 | $10^{7}$ | 0·00764 | 0·00837 | 0·00866 | 2·31134 | 1·23467 | 0·43538 | 0·22251 | 0·07230 | 2·31134 |
| 367 | $10^{8}$ | 0·00087 | 0·00076 | 0·00066 | 0·00142 | 0·00083 | 0·02790 | 0·06279 | 0·11848 | 0·11848 |
| 503 | $10^{7}$ | 0·00787 | 0·00764 | 0·00832 | 0·09016 | 0·04984 | 0·03909 | 0·04295 | 0·08024 | 0·09016 |
| 501 | $10^{8}$ | 0·00404 | 0·00400 | 0·00490 | 0·00629 | 0·00616 | 0·01773 | 0·06905 | 0·15288 | 0·15288 |
| 751 | $10^{10}$ | 0·00815 | 0·00801 | 0·01056 | 0·01120 | 0·01153 | 0·01305 | 0·01632 | 0·02353 | 0·02353 |
| 546 | $10^{8}$ | 0·01246 | 0·01342 | 0·01285 | 0·01217 | 0·01325 | 0·02392 | 0·02033 | 0·06327 | 0·06327 |
| 945 | $10^{8}$ | 0·01150 | 0·01121 | 0·01150 | 0·01074 | 0·01214 | 0·01020 | 0·01503 | 0·07338 | 0·07338 |
| 802 | $10^{10}$ | 0·00168 | 0·00157 | 0·00208 | 0·00580 | | | 0·00265 | 0·00168 | 0·00580 |
| 960 | $10^{8}$ | 0·00977 | 0·00909 | 0·00793 | 0·08510 | 0·04097 | 0·03496 | 0·01479 | 0·01644 | 0·08510 |
| 375 | $10^{5}$ | 0·00140 | 0·00130 | 0·00205 | 0·00660 | 0·00423 | 0·00433 | 0·00456 | 0·00753 | 0·00753 |
| 350 | $10^{5}$ | 0·00250 | 0·00239 | 0·00268 | 2·22367 | 0·94033 | 0·17544 | 0·13084 | 0·07709 | 2·22367 |
| 1111 | $10^{6}$ | 0·00241 | 0·00286 | 0·00338 | 0·00497 | 0·00590 | 0·01282 | 0·01726 | 0·02262 | 0·02262 |
| 576 | $10^{7}$ | 0·00243 | 0·00199 | 0·00250 | 0·00364 | 0·00374 | 0·00095 | | 0·00055 | 0·00374 |
| 520 | $10^{7}$ | 0·00525 | 0·00504 | 0·00557 | 0·00580 | 0·00670 | 0·00694 | 0·00906 | 0·03351 | 0·03351 |

| Primary fold change Inaba OSP IgG | | | | | | | | | |
|---|---|---|---|---|---|---|---|---|---|
| Dose (CFU) | D1 | D4 | D7 | D15 | D29 | D57 | D85 | D180 | max FC |
| $10^{8}$ | 1·00 | 1·10 | 1·09 | 1·21 | 0·26 | 1·51 | 3·19 | 4·80 | 4·80 |
| $10^{8}$ | 1·00 | 1·64 | 1·68 | 0·83 | 0·05 | | | 0·03 | 1·68 |
| $10^{7}$ | 1·00 | 1·20 | 0·87 | 1·34 | 1·56 | 1·45 | 1·20 | 1·30 | 1·56 |
| $10^{7}$ | 1·00 | 0·85 | 0·83 | 0·91 | 0·85 | 2·49 | 2·95 | 7·20 | 7·20 |
| $10^{8}$ | 1·00 | 1·20 | 0·84 | 1·06 | 1·36 | 1·15 | 0·99 | 1·17 | 1·36 |
| $10^{8}$ | 1·00 | 0·71 | 1·11 | 1·00 | 0·56 | 0·47 | 0·70 | 4·00 | 4·00 |
| $10^{7}$ | 1·00 | 1·10 | 1·13 | 302·53 | 161·61 | 56·99 | 29·12 | 9·46 | 302·53 |
| $10^{8}$ | 1·00 | 0·87 | 0·76 | 1·63 | 0·95 | 32·07 | 72·17 | 136·18 | 136·18 |
| $10^{7}$ | 1·00 | 0·97 | 1·06 | 11·46 | 6·33 | 4·97 | 5·46 | 10·20 | 11·46 |
| $10^{8}$ | 1·00 | 0·99 | 1·21 | 1·56 | 1·52 | 4·39 | 17·09 | 37·84 | 37·84 |
| $10^{10}$ | 1·00 | 0·98 | 1·30 | 1·37 | 1·41 | 1·60 | 2·00 | 2·89 | 2·89 |
| $10^{8}$ | 1·00 | 1·08 | 1·03 | 0·98 | 1·06 | 1·92 | 1·63 | 5·08 | 5·08 |
| $10^{8}$ | 1·00 | 0·97 | 1·00 | 0·93 | 1·06 | 0·89 | 1·31 | 6·38 | 6·38 |
| $10^{10}$ | 1·00 | 0·93 | 1·24 | 3·45 | | | 1·58 | 1·00 | 3·45 |
| $10^{8}$ | 1·00 | 0·93 | 0·81 | 8·71 | 4·19 | 3·58 | 1·51 | 1·68 | 8·71 |
| $10^{5}$ | 1·00 | 0·93 | 1·46 | 4·71 | 3·02 | 3·09 | 3·26 | 5·38 | 5·38 |
| $10^{5}$ | 1·00 | 0·96 | 1·07 | 889·47 | 376·13 | 70·18 | 52·34 | 30·84 | 889·47 |
| $10^{6}$ | 1·00 | 1·19 | 1·40 | 2·06 | 2·45 | 5·32 | 7·16 | 9·39 | 9·39 |
| $10^{7}$ | 1·00 | 0·82 | 1·03 | 1·50 | 1·54 | 0·39 | | 0·23 | 1·54 |
| $10^{7}$ | 1·00 | 0·96 | 1·06 | 1·10 | 1·28 | 1·32 | 1·73 | 6·38 | 6·38 |

(grey box) below limit of detection

| Boost Inaba OSP IgA (RAU) | | | | | | | | | | |
|---|---|---|---|---|---|---|---|---|---|---|
| **Dose interval (days)** | **Dose (CFU)** | **D1** | **D4** | **D7** | **D15** | **D29** | **D57** | **D85** | **D180** | **max** |
| 389 | $2\times10^{7}$ | 0·00388 | 0·00290 | 0·60801 | 0·31811 | 0·00650 | 0·02911 | 0·00320 | 0·00422 | 0·60801 |
| 405 | $2\times10^{7}$ | 0·00113 | 0·00128 | 0·02326 | 0·35503 | 0·03543 | 0·00452 | 0·00258 | 0·00223 | 0·35503 |
| 322 | $2\times10^{7}$ | 0·00145 | 0·00115 | 0·04920 | 0·06769 | 0·02923 | 0·00473 | 0·00503 | 0·00412 | 0·06769 |
| 418 | $2\times10^{7}$ | 0·00494 | 0·00470 | 0·02130 | 0·02766 | 0·01178 | 0·00877 | 0·01149 | 0·00709 | 0·02766 |
| 378 | $2\times10^{7}$ | 0·00537 | 0·00547 | 0·39488 | 1·02596 | 0·16110 | 0·03189 | 0·01215 | 0·01046 | 1·02596 |
| 399 | $2\times10^{7}$ | 0·00512 | 0·00501 | 0·00479 | 0·00430 | 0·00570 | 0·00685 | 0·00653 | 0·00698 | 0·00698 |
| 568 | $2\times10^{7}$ | 0·00737 | 0·00534 | 0·07858 | 0·42300 | 0·08046 | 0·02305 | 0·01335 | 0·00766 | 0·42300 |
| 367 | $2\times10^{7}$ | 0·00205 | 0·00205 | 0·37231 | 0·13499 | 0·01444 | 0·00490 | 0·00357 | 0·00331 | 0·37231 |
| 503 | $2\times10^{7}$ | 0·02697 | 0·03213 | 0·02712 | 0·03329 | 0·02476 | 0·02854 | 0·02986 | 0·02432 | 0·03329 |
| 501 | $2\times10^{7}$ | 0·00765 | 0·00744 | 0·11609 | 0·08415 | 0·03212 | 0·01340 | 0·01224 | 0·01387 | 0·11609 |
| 751 | $2\times10^{7}$ | 0·00626 | 0·00626 | 0·07881 | 0·09562 | 0·02195 | 0·00930 | 0·00958 | 0·00735 | 0·09562 |
| 546 | $2\times10^{7}$ | 0·02273 | 0·01950 | 0·06584 | 0·04793 | 0·03111 | 0·02161 | 0·01917 | 0·01939 | 0·06584 |
| 945 | $2\times10^{7}$ | 0·00460 | 0·00415 | 0·20128 | 0·26212 | 0·06984 | 0·02053 | 0·01500 | 0·01351 | 0·26212 |
| 802 | $2\times10^{7}$ | 0·00714 | 0·00770 | 0·71243 | 0·20215 | 0·08845 | | | 0·02209 | 0·71243 |
| 960 | $2\times10^{7}$ | 0·00861 | 0·00881 | 0·28033 | 0·24732 | 0·08121 | 0·02502 | 0·01983 | 0·03022 | 0·28033 |
| 375 | $2\times10^{7}$ | 0·00495 | 0·00433 | 0·22648 | 0·12016 | 0·03958 | 0·01331 | 0·00898 | 0·01559 | 0·22648 |
| 350 | $2\times10^{7}$ | 0·01379 | 0·01321 | 0·12664 | 0·08940 | 0·03482 | 0·01939 | 0·01719 | 0·02919 | 0·12664 |
| 1111 | $2\times10^{7}$ | 0·01145 | 0·01365 | 0·01334 | 0·01152 | 0·01178 | 0·01083 | 0·01334 | 0·01020 | 0·01365 |
| 576 | $2\times10^{7}$ | 0·00333 | 0·00393 | 0·09097 | 0·05733 | 0·01354 | 0·00405 | 0·00339 | 0·00293 | 0·09097 |
| 520 | $2\times10^{7}$ | 0·00602 | 0·00715 | 0·30720 | 0·16067 | 0·03635 | 0·01290 | 0·01583 | 0·00992 | 0·30720 |

| Boost fold change Inaba OSP IgA | | | | | | | | | |
|---|---|---|---|---|---|---|---|---|---|
| **Dose (CFU)** | **D1** | **D4** | **D7** | **D15** | **D29** | **D57** | **D85** | **D180** | **max FC** |
| $2\times10^{7}$ | 1·00 | 0·75 | 156·62 | 81·95 | 1·67 | 7·50 | 0·82 | 1·09 | 156·62 |
| $2\times10^{7}$ | 1·00 | 1·13 | 20·63 | 314·88 | 31·42 | 4·01 | 2·29 | 1·97 | 314·88 |
| $2\times10^{7}$ | 1·00 | 0·79 | 33·92 | 46·67 | 20·16 | 3·26 | 3·47 | 2·84 | 46·67 |
| $2\times10^{7}$ | 1·00 | 0·95 | 4·32 | 5·60 | 2·39 | 1·78 | 2·33 | 1·44 | 5·60 |
| $2\times10^{7}$ | 1·00 | 1·02 | 73·53 | 191·05 | 30·00 | 5·94 | 2·26 | 1·95 | 191·05 |
| $2\times10^{7}$ | 1·00 | 0·98 | 0·94 | 0·84 | 1·11 | 1·34 | 1·28 | 1·36 | 1·36 |
| $2\times10^{7}$ | 1·00 | 0·72 | 10·66 | 57·37 | 10·91 | 3·13 | 1·81 | 1·04 | 57·37 |
| $2\times10^{7}$ | 1·00 | 1·00 | 181·21 | 65·70 | 7·03 | 2·38 | 1·74 | 1·61 | 181·21 |
| $2\times10^{7}$ | 1·00 | 1·19 | 1·01 | 1·23 | 0·92 | 1·06 | 1·11 | 0·90 | 1·23 |
| $2\times10^{7}$ | 1·00 | 0·97 | 15·18 | 11·00 | 4·20 | 1·75 | 1·60 | 1·81 | 15·18 |
| $2\times10^{7}$ | 1·00 | 1·00 | 12·59 | 15·27 | 3·51 | 1·48 | 1·53 | 1·17 | 15·27 |
| $2\times10^{7}$ | 1·00 | 0·86 | 2·90 | 2·11 | 1·37 | 0·95 | 0·84 | 0·85 | 2·90 |
| $2\times10^{7}$ | 1·00 | 0·90 | 43·76 | 56·99 | 15·18 | 4·46 | 3·26 | 2·94 | 56·99 |
| $2\times10^{7}$ | 1·00 | 1·08 | 99·81 | 28·32 | 12·39 | | | 3·09 | 99·81 |
| $2\times10^{7}$ | 1·00 | 1·02 | 32·55 | 28·72 | 9·43 | 2·91 | 2·30 | 3·51 | 32·55 |
| $2\times10^{7}$ | 1·00 | 0·87 | 45·72 | 24·25 | 7·99 | 2·69 | 1·81 | 3·15 | 45·72 |
| $2\times10^{7}$ | 1·00 | 0·96 | 9·18 | 6·48 | 2·52 | 1·41 | 1·25 | 2·12 | 9·18 |
| $2\times10^{7}$ | 1·00 | 1·19 | 1·17 | 1·01 | 1·03 | 0·95 | 1·17 | 0·89 | 1·19 |
| $2\times10^{7}$ | 1·00 | 1·18 | 27·31 | 17·21 | 4·06 | 1·22 | 1·02 | 0·88 | 27·31 |
| $2\times10^{7}$ | 1·00 | 1·19 | 51·04 | 26·69 | 6·04 | 2·14 | 2·63 | 1·65 | 51·04 |

| Primary Inaba OSP IgA (RAU) | | | | | | | | | | |
|---|---|---|---|---|---|---|---|---|---|---|
| **Dose interval (days)** | **Dose (CFU)** | **D1** | **D4** | **D7** | **D15** | **D29** | **D57** | **D85** | **D180** | **max** |
| 389 | $10^{8}$ | 0·00067 | 0·00017 | 0·00034 | 0·00178 | 0·00110 | 0·00119 | 0·00141 | 0·00106 | 0·00178 |
| 405 | $10^{8}$ | 0·00112 | 0·00047 | 0·00100 | 0·00097 | 0·00044 | 0·00059 | 0·00070 | 0·00073 | 0·00112 |
| 322 | $10^{7}$ | 0·00028 | 0·00015 | 0·00236 | 0·02150 | 0·00751 | 0·00211 | 0·00125 | 0·00050 | 0·02150 |
| 418 | $10^{7}$ | 0·00195 | 0·00237 | 0·00392 | 0·01994 | 0·01552 | 0·00686 | 0·00632 | 0·00543 | 0·01994 |
| 378 | $10^{8}$ | 0·00071 | 0·00113 | 0·00158 | 0·01213 | 0·00951 | 0·00520 | 0·00477 | 0·00315 | 0·01213 |
| 399 | $10^{8}$ | 0·00743 | 0·00677 | 0·01973 | 0·06637 | 0·02397 | 0·00980 | 0·00731 | 0·00505 | 0·06637 |
| 568 | $10^{7}$ | 0·00104 | 0·00136 | 0·00384 | 4·00331 | 0·80326 | 0·07279 | 0·04657 | 0·02318 | 4·00331 |
| 367 | $10^{8}$ | 0·00033 | 0·00041 | 0·00047 | 0·00287 | 0·00165 | 0·00219 | 0·00172 | 0·00195 | 0·00287 |
| 503 | $10^{7}$ | 0·00189 | 0·00186 | 0·00289 | 0·32011 | 0·03853 | 0·02678 | 0·03879 | 0·03264 | 0·32011 |
| 501 | $10^{8}$ | 0·00079 | 0·00045 | 0·00138 | 0·09273 | 0·00891 | 0·00669 | 0·00599 | 0·00542 | 0·09273 |
| 751 | $10^{10}$ | 0·00008 | 0·00005 | 0·00403 | 0·11487 | 0·01491 | 0·00439 | 0·00373 | 0·00443 | 0·11487 |
| 546 | $10^{8}$ | 0·00300 | 0·00358 | 0·00464 | 0·00716 | 0·00928 | 0·01226 | 0·00552 | 0·00590 | 0·01226 |
| 945 | $10^{8}$ | 0·00032 | 0·00032 | 0·00036 | 0·00111 | 0·00047 | 0·00077 | 0·00097 | 0·00206 | 0·00206 |
| 802 | $10^{10}$ | 0·00075 | 0·00083 | 0·00319 | 0·02563 | | | 0·01147 | 0·00367 | 0·02563 |
| 960 | $10^{8}$ | 0·00090 | 0·00093 | 0·00058 | 0·00321 | 0·00128 | 0·00110 | 0·00352 | 0·00462 | 0·00462 |
| 375 | $10^{5}$ | 0·00175 | 0·00139 | 0·00190 | 0·00196 | 0·00191 | 0·00186 | 0·00193 | 0·00464 | 0·00464 |
| 350 | $10^{5}$ | 0·00140 | 0·00167 | 0·00194 | 0·04121 | 0·01029 | 0·00513 | 0·00761 | 0·01339 | 0·04121 |
| 1111 | $10^{6}$ | 0·00191 | 0·00208 | 0·01421 | 0·11625 | 0·02677 | 0·01284 | 0·01007 | 0·00819 | 0·11625 |
| 576 | $10^{7}$ | 0·00121 | 0·00214 | 0·00243 | 0·00884 | 0·00831 | 0·00326 | 0·00224 | 0·00097 | 0·00884 |
| 520 | $10^{7}$ | 0·00092 | 0·00106 | 0·00156 | 0·00133 | 0·00197 | 0·00291 | 0·00382 | 0·00624 | 0·00624 |

| Primary fold change Inaba OSP IgA | | | | | | | | | |
|---|---|---|---|---|---|---|---|---|---|
| **Dose (CFU)** | **D1** | **D4** | **D7** | **D15** | **D29** | **D57** | **D85** | **D180** | **max FC** |
| $10^{8}$ | 1·00 | 0·25 | 0·51 | 2·66 | 1·64 | 1·78 | 2·10 | 1·58 | 2·66 |
| $10^{8}$ | 1·00 | 0·42 | 0·89 | 0·87 | 0·39 | 0·53 | 0·62 | 0·65 | 0·89 |
| $10^{7}$ | 1·00 | 0·54 | 8·43 | 76·79 | 26·82 | 7·54 | 4·46 | 1·79 | 76·79 |
| $10^{7}$ | 1·00 | 1·22 | 2·01 | 10·23 | 7·96 | 3·52 | 3·24 | 2·78 | 10·23 |
| $10^{8}$ | 1·00 | 1·59 | 2·23 | 17·08 | 13·39 | 7·32 | 6·72 | 4·44 | 17·08 |
| $10^{8}$ | 1·00 | 0·91 | 2·66 | 8·93 | 3·23 | 1·32 | 0·98 | 0·68 | 8·93 |
| $10^{7}$ | 1·00 | 1·31 | 3·69 | 3849·34 | 772·37 | 69·99 | 44·78 | 22·29 | 3849·34 |
| $10^{8}$ | 1·00 | 1·24 | 1·42 | 8·70 | 5·00 | 6·64 | 5·21 | 5·91 | 8·70 |
| $10^{7}$ | 1·00 | 0·98 | 1·53 | 169·37 | 20·39 | 14·17 | 20·52 | 17·27 | 169·37 |
| $10^{8}$ | 1·00 | 0·57 | 1·75 | 117·38 | 11·28 | 8·47 | 7·58 | 6·86 | 117·38 |
| $10^{10}$ | 1·00 | 0·62 | 50·37 | 1435·87 | 186·37 | 54·87 | 46·62 | 55·37 | 1435·87 |
| $10^{8}$ | 1·00 | 1·19 | 1·55 | 2·39 | 3·09 | 4·09 | 1·84 | 1·97 | 4·09 |
| $10^{8}$ | 1·00 | 1·00 | 1·12 | 3·47 | 1·47 | 2·41 | 3·03 | 6·44 | 6·44 |
| $10^{10}$ | 1·00 | 1·11 | 4·25 | 34·17 | | | 15·29 | 4·89 | 34·17 |
| $10^{8}$ | 1·00 | 1·03 | 0·64 | 3·57 | 1·42 | 1·22 | 3·91 | 5·13 | 5·13 |
| $10^{5}$ | 1·00 | 0·79 | 1·09 | 1·12 | 1·09 | 1·06 | 1·10 | 2·65 | 2·65 |
| $10^{5}$ | 1·00 | 1·19 | 1·39 | 29·44 | 7·35 | 3·66 | 5·44 | 9·57 | 29·44 |
| $10^{6}$ | 1·00 | 1·09 | 7·44 | 60·86 | 14·02 | 6·72 | 5·27 | 4·29 | 60·86 |
| $10^{7}$ | 1·00 | 1·77 | 2·01 | 7·31 | 6·87 | 2·69 | 1·85 | 0·80 | 7·31 |
| $10^{7}$ | 1·00 | 1·15 | 1·70 | 1·45 | 2·14 | 3·16 | 4·15 | 6·78 | 6·78 |

| Boost Ogawa OSP IgM (RAU) | | | | | | | | | | |
|---|---|---|---|---|---|---|---|---|---|---|
| Dose interval (days) | Dose (CFU) | D1 | D4 | D7 | D15 | D29 | D57 | D85 | D180 | max |
| 389 | $2\times10^7$ | 0·02105 | 0·02133 | 0·14128 | 0·11310 | 0·01973 | 0·03771 | 0·01281 | 0·01575 | 0·14128 |
| 405 | $2\times10^7$ | 0·00049 | 0·00041 | 0·01091 | 0·03138 | 0·01092 | 0·00262 | 0·00150 | 0·00128 | 0·03138 |
| 322 | $2\times10^7$ | 0·00423 | 0·00369 | 0·00705 | 0·00590 | 0·00528 | 0·00422 | 0·00240 | 0·00232 | 0·00705 |
| 418 | $2\times10^7$ | 0·00037 | 0·00040 | 0·00199 | 0·00103 | 0·00076 | 0·00183 | 0·00114 | 0·00068 | 0·00199 |
| 378 | $2\times10^7$ | 0·01817 | 0·01946 | 0·03308 | 0·07060 | 0·04124 | 0·02961 | 0·01573 | 0·01036 | 0·07060 |
| 399 | $2\times10^7$ | 0·01276 | 0·01197 | 0·01111 | 0·01156 | 0·01244 | 0·01425 | 0·01033 | 0·00988 | 0·01425 |
| 568 | $2\times10^7$ | 0·02233 | 0·01862 | 0·03745 | 0·09731 | 0·05390 | 0·03826 | 0·02031 | 0·01342 | 0·09731 |
| 367 | $2\times10^7$ | 0·00241 | 0·00254 | 0·03682 | 0·02518 | 0·01040 | 0·00475 | 0·00271 | 0·00253 | 0·03682 |
| 503 | $2\times10^7$ | 0·00290 | 0·00306 | 0·00265 | 0·00308 | 0·00364 | 0·00264 | 0·00229 | 0·00157 | 0·00364 |
| 501 | $2\times10^7$ | 0·00291 | 0·00213 | 0·08226 | 0·07675 | 0·04236 | 0·01303 | 0·00471 | 0·00267 | 0·08226 |
| 751 | $2\times10^7$ | 0·00277 | 0·00331 | 0·01506 | 0·01533 | 0·00673 | 0·00363 | 0·00418 | 0·00403 | 0·01533 |
| 546 | $2\times10^7$ | 0·09937 | 0·08794 | 0·07550 | 0·08102 | 0·07789 | 0·07397 | 0·07447 | 0·05102 | 0·09937 |
| 945 | $2\times10^7$ | 0·00736 | 0·00673 | 0·06722 | 0·06800 | 0·02523 | 0·01014 | 0·00773 | 0·00219 | 0·06800 |
| 802 | $2\times10^7$ | 0·01073 | 0·01166 | 0·32285 | 0·18883 | 0·11562 | | | 0·00723 | 0·32285 |
| 960 | $2\times10^7$ | 0·00212 | 0·00200 | 0·05600 | 0·04813 | 0·02489 | 0·00683 | 0·00373 | 0·00424 | 0·05600 |
| 375 | $2\times10^7$ | 0·00118 | 0·00104 | 0·03749 | 0·02075 | 0·01027 | 0·00414 | 0·00252 | 0·00157 | 0·03749 |
| 350 | $2\times10^7$ | 0·00550 | 0·00562 | 0·00897 | 0·00662 | 0·00600 | 0·00537 | 0·00509 | 0·00513 | 0·00897 |
| 1111 | $2\times10^7$ | 0·00255 | 0·00292 | 0·00187 | 0·00222 | 0·00193 | 0·00182 | 0·00247 | 0·00186 | 0·00292 |
| 576 | $2\times10^7$ | 0·00505 | 0·00503 | 0·02391 | 0·01387 | 0·01168 | 0·00711 | 0·00477 | 0·00379 | 0·02391 |
| 520 | $2\times10^7$ | 0·01103 | 0·01544 | 0·08777 | 0·05921 | 0·03682 | 0·02372 | 0·01473 | 0·00885 | 0·08777 |

| Boost fold change Ogawa OSP IgM | | | | | | | | | |
|---|---|---|---|---|---|---|---|---|---|
| Dose (CFU) | D1 | D4 | D7 | D15 | D29 | D57 | D85 | D180 | max FC |
| $2\times10^7$ | 1·00 | 1·01 | 6·71 | 5·37 | 0·94 | 1·79 | 0·61 | 0·75 | 6·71 |
| $2\times10^7$ | 1·00 | 0·84 | 22·27 | 64·09 | 22·31 | 5·35 | 3·06 | 2·62 | 64·09 |
| $2\times10^7$ | 1·00 | 0·87 | 1·67 | 1·40 | 1·25 | 1·00 | 0·57 | 0·55 | 1·67 |
| $2\times10^7$ | 1·00 | 1·09 | 5·39 | 2·80 | 2·05 | 4·97 | 3·10 | 1·85 | 5·39 |
| $2\times10^7$ | 1·00 | 1·07 | 1·82 | 3·89 | 2·27 | 1·63 | 0·87 | 0·57 | 3·89 |
| $2\times10^7$ | 1·00 | 0·94 | 0·87 | 0·91 | 0·98 | 1·12 | 0·81 | 0·77 | 1·12 |
| $2\times10^7$ | 1·00 | 0·83 | 1·68 | 4·36 | 2·41 | 1·71 | 0·91 | 0·60 | 4·36 |
| $2\times10^7$ | 1·00 | 1·05 | 15·29 | 10·46 | 4·32 | 1·97 | 1·12 | 1·05 | 15·29 |
| $2\times10^7$ | 1·00 | 1·05 | 0·91 | 1·06 | 1·25 | 0·91 | 0·79 | 0·54 | 1·25 |
| $2\times10^7$ | 1·00 | 0·73 | 28·23 | 26·33 | 14·53 | 4·47 | 1·62 | 0·92 | 28·23 |
| $2\times10^7$ | 1·00 | 1·19 | 5·43 | 5·53 | 2·42 | 1·31 | 1·51 | 1·45 | 5·53 |
| $2\times10^7$ | 1·00 | 0·89 | 0·76 | 0·82 | 0·78 | 0·74 | 0·75 | 0·51 | 0·89 |
| $2\times10^7$ | 1·00 | 0·91 | 9·13 | 9·24 | 3·43 | 1·38 | 1·05 | 0·30 | 9·24 |
| $2\times10^7$ | 1·00 | 1·09 | 30·09 | 17·60 | 10·78 | | | 0·67 | 30·09 |
| $2\times10^7$ | 1·00 | 0·95 | 26·45 | 22·73 | 11·76 | 3·22 | 1·76 | 2·00 | 26·45 |
| $2\times10^7$ | 1·00 | 0·88 | 31·69 | 17·54 | 8·68 | 3·50 | 2·13 | 1·33 | 31·69 |
| $2\times10^7$ | 1·00 | 1·02 | 1·63 | 1·20 | 1·09 | 0·98 | 0·93 | 0·93 | 1·63 |
| $2\times10^7$ | 1·00 | 1·15 | 0·73 | 0·87 | 0·76 | 0·71 | 0·97 | 0·73 | 1·15 |
| $2\times10^7$ | 1·00 | 1·00 | 4·74 | 2·75 | 2·31 | 1·41 | 0·94 | 0·75 | 4·74 |
| $2\times10^7$ | 1·00 | 1·40 | 7·96 | 5·37 | 3·34 | 2·15 | 1·34 | 0·80 | 7·96 |

| Primary Ogawa OSP IgM (RAU) | | | | | | | | | | |
|---|---|---|---|---|---|---|---|---|---|---|
| Dose interval (days) | Dose (CFU) | D1 | D4 | D7 | D15 | D29 | D57 | D85 | D180 | max |
| 389 | $10^8$ | 0·00769 | 0·00931 | 0·01095 | 0·10439 | 0·06109 | 0·02945 | 0·02604 | 0·02080 | 0·10439 |
| 405 | $10^8$ | 0·00069 | 0·00077 | 0·00063 | 0·02231 | 0·00679 | 0·00151 | 0·00140 | 0·00057 | 0·02231 |
| 322 | $10^7$ | 0·00189 | 0·00166 | 0·00284 | 0·00553 | 0·00299 | 0·00231 | 0·00218 | 0·00261 | 0·00553 |
| 418 | $10^7$ | 0·00022 | 0·00051 | 0·00032 | 0·00154 | 0·00114 | 0·00082 | 0·00085 | 0·00032 | 0·00154 |
| 378 | $10^8$ | 0·01143 | 0·01262 | 0·17239 | 1·68965 | 1·02668 | 0·24002 | 0·09532 | 0·03970 | 1·68965 |
| 399 | $10^8$ | 0·00331 | 0·00269 | 0·15740 | 4·28939 | 1·05323 | 0·23211 | 0·15355 | 0·07635 | 4·28939 |
| 568 | $10^7$ | 0·01969 | 0·02122 | 0·09912 | 2·77280 | 1·66929 | 0·60782 | 0·29698 | 0·10815 | 2·77280 |
| 367 | $10^8$ | 0·00095 | 0·00134 | 0·05602 | 5·67692 | 2·05979 | 0·26296 | 0·07343 | 0·00936 | 5·67692 |
| 503 | $10^7$ | 0·00403 | 0·00353 | 0·02376 | 0·27542 | 0·10344 | 0·02921 | 0·01780 | 0·00335 | 0·27542 |
| 501 | $10^8$ | 0·00376 | 0·00315 | 0·01289 | 0·11659 | 0·02460 | 0·01327 | 0·00897 | 0·00267 | 0·11659 |
| 751 | $10^{10}$ | 0·00277 | 0·00272 | 0·37863 | 1·04964 | 0·26434 | 0·09112 | 0·02412 | 0·01330 | 1·04964 |
| 546 | $10^8$ | 0·02280 | 0·02315 | 0·05721 | 0·11660 | 0·11261 | 0·09070 | 0·11108 | 0·10286 | 0·11660 |
| 945 | $10^8$ | 0·00108 | 0·00106 | 0·00247 | 0·11431 | 0·06306 | 0·02353 | 0·01882 | 0·00874 | 0·11431 |
| 802 | $10^{10}$ | 0·00903 | 0·00963 | 0·03222 | 0·14568 | | | 0·01764 | 0·00956 | 0·14568 |
| 960 | $10^8$ | 0·00210 | 0·00167 | 0·00389 | 0·11895 | 0·03692 | 0·01398 | 0·00621 | 0·00468 | 0·11895 |
| 375 | $10^5$ | 0·00079 | 0·00053 | 0·00039 | 0·00253 | 0·00345 | 0·00236 | 0·00244 | 0·00102 | 0·00345 |
| 350 | $10^5$ | 0·01259 | 0·01316 | 0·01319 | 0·01754 | 0·01359 | 0·00987 | 0·01069 | 0·00340 | 0·01754 |
| 1111 | $10^6$ | 0·00076 | 0·00101 | 0·00146 | 0·00554 | 0·00366 | 0·00262 | 0·00176 | 0·00118 | 0·00554 |
| 576 | $10^7$ | 0·00959 | 0·01022 | 0·02270 | 1·67601 | 1·06496 | 0·49654 | 0·26575 | 0·05372 | 1·67601 |
| 520 | $10^7$ | 0·00156 | 0·00164 | 0·00286 | 0·02562 | 0·02084 | 0·02080 | 0·02200 | 0·01866 | 0·02562 |

| Primary fold change Ogawa OSP IgM | | | | | | | | | |
|---|---|---|---|---|---|---|---|---|---|
| Dose (CFU) | D1 | D4 | D7 | D15 | D29 | D57 | D85 | D180 | max FC |
| $10^8$ | 1·00 | 1·21 | 1·42 | 13·57 | 7·94 | 3·83 | 3·39 | 2·70 | 13·57 |
| $10^8$ | 1·00 | 1·12 | 0·91 | 32·33 | 9·84 | 2·19 | 2·03 | 0·83 | 32·33 |
| $10^7$ | 1·00 | 0·88 | 1·50 | 2·93 | 1·58 | 1·22 | 1·15 | 1·38 | 2·93 |
| $10^7$ | 1·00 | 2·32 | 1·45 | 7·00 | 5·18 | 3·73 | 3·86 | 1·45 | 7·00 |
| $10^8$ | 1·00 | 1·10 | 15·08 | 147·83 | 89·82 | 21·00 | 8·34 | 3·47 | 147·83 |
| $10^8$ | 1·00 | 0·81 | 47·55 | 1295·89 | 318·20 | 70·12 | 46·39 | 23·07 | 1295·89 |
| $10^7$ | 1·00 | 1·08 | 5·03 | 140·82 | 84·78 | 30·87 | 15·08 | 5·49 | 140·82 |
| $10^8$ | 1·00 | 1·41 | 58·97 | 5975·71 | 2168·20 | 276·80 | 77·29 | 9·85 | 5975·71 |
| $10^7$ | 1·00 | 0·88 | 5·90 | 68·34 | 25·67 | 7·25 | 4·42 | 0·83 | 68·34 |
| $10^8$ | 1·00 | 0·84 | 3·43 | 31·01 | 6·54 | 3·53 | 2·39 | 0·71 | 31·01 |
| $10^{10}$ | 1·00 | 0·98 | 136·69 | 378·93 | 95·43 | 32·90 | 8·71 | 4·80 | 378·93 |
| $10^8$ | 1·00 | 1·02 | 2·51 | 5·11 | 4·94 | 3·98 | 4·87 | 4·51 | 5·11 |
| $10^8$ | 1·00 | 0·98 | 2·29 | 105·84 | 58·39 | 21·79 | 17·43 | 8·09 | 105·84 |
| $10^{10}$ | 1·00 | 1·07 | 3·57 | 16·13 | | | 1·95 | 1·06 | 16·13 |
| $10^8$ | 1·00 | 0·80 | 1·85 | 56·64 | 17·58 | 6·66 | 2·96 | 2·23 | 56·64 |
| $10^5$ | 1·00 | 0·67 | 0·49 | 3·20 | 4·37 | 2·99 | 3·09 | 1·29 | 4·37 |
| $10^5$ | 1·00 | 1·05 | 1·05 | 1·39 | 1·08 | 0·78 | 0·85 | 0·27 | 1·39 |
| $10^6$ | 1·00 | 1·33 | 1·92 | 7·29 | 4·82 | 3·45 | 2·32 | 1·55 | 7·29 |
| $10^7$ | 1·00 | 1·07 | 2·37 | 174·77 | 111·05 | 51·78 | 27·71 | 5·60 | 174·77 |
| $10^7$ | 1·00 | 1·05 | 1·83 | 16·42 | 13·36 | 13·33 | 14·10 | 11·96 | 16·42 |

| Boost Ogawa OSP IgG (RAU) | | | | | | | | | | |
|---|---|---|---|---|---|---|---|---|---|---|
| Dose interval (days) | Dose (CFU) | D1 | D4 | D7 | D15 | D29 | D57 | D85 | D180 | max |
| 389 | $2\times10^{7}$ | 0·01362 | 0·01050 | 0·21199 | 0·44297 | 0·11349 | 0·23682 | 0·02766 | 0·00846 | 0·44297 |
| 405 | $2\times10^{7}$ | 0·00061 | 0·00081 | 0·00436 | 0·01268 | 0·00460 | 0·00153 | 0·00066 | 0·00077 | 0·01268 |
| 322 | $2\times10^{7}$ | 0·00199 | 0·00145 | 0·00274 | 0·00335 | 0·00342 | 0·00307 | 0·00100 | 0·00084 | 0·00342 |
| 418 | $2\times10^{7}$ | 0·00597 | 0·00557 | 0·00692 | 0·01025 | 0·00889 | 0·03382 | 0·00695 | 0·00356 | 0·03382 |
| 378 | $2\times10^{7}$ | 0·00297 | 0·00281 | 0·00477 | 0·01310 | 0·00822 | 0·00586 | 0·00271 | 0·00167 | 0·01310 |
| 399 | $2\times10^{7}$ | 0·02613 | 0·02277 | 0·02165 | 0·02387 | 0·02338 | 0·02253 | 0·02006 | 0·01912 | 0·02613 |
| 568 | $2\times10^{7}$ | 0·00197 | 0·00163 | 0·03050 | 0·32337 | 0·16274 | 0·09843 | 0·04768 | 0·00409 | 0·32337 |
| 367 | $2\times10^{7}$ | 0·00775 | 0·00951 | 0·98285 | 0·62582 | 0·27634 | 0·09192 | 0·03201 | 0·00837 | 0·98285 |
| 503 | $2\times10^{7}$ | 0·00552 | 0·00622 | 0·00534 | 0·00598 | 0·00468 | 0·00433 | 0·00431 | 0·00487 | 0·00622 |
| 501 | $2\times10^{7}$ | 0·00810 | 0·00886 | 0·02711 | 0·05885 | 0·05908 | 0·02271 | 0·01745 | 0·01404 | 0·05908 |
| 751 | $2\times10^{7}$ | 0·01720 | 0·01764 | 0·03266 | 0·05517 | 0·04360 | 0·02926 | 0·03452 | 0·01070 | 0·05517 |
| 546 | $2\times10^{7}$ | 0·00519 | 0·00341 | 0·00960 | 0·00602 | 0·00699 | 0·00456 | 0·00503 | 0·00394 | 0·00960 |
| 945 | $2\times10^{7}$ | 0·00495 | 0·00502 | 0·11458 | 0·72713 | 0·20079 | 0·10831 | 0·08340 | 0·00909 | 0·72713 |
| 802 | $2\times10^{7}$ | 0·00122 | 0·00143 | 0·04226 | 0·02884 | 0·02236 | | | 0·00293 | 0·04226 |
| 960 | $2\times10^{7}$ | 0·00131 | 0·00103 | 0·03019 | 0·04762 | 0·03446 | 0·01632 | 0·00971 | 0·00897 | 0·04762 |
| 375 | $2\times10^{7}$ | 0·00315 | 0·00334 | 0·05115 | 0·04351 | 0·03255 | 0·02485 | 0·01604 | 0·01355 | 0·05115 |
| 350 | $2\times10^{7}$ | 0·00857 | 0·00746 | 0·05112 | 0·05336 | 0·03149 | 0·01717 | 0·01157 | 0·02526 | 0·05336 |
| 1111 | $2\times10^{7}$ | 0·00052 | 0·00082 | 0·00051 | 0·00047 | 0·00042 | 0·00042 | 0·00151 | 0·00143 | 0·00151 |
| 576 | $2\times10^{7}$ | 0·00051 | 0·00043 | 0·00144 | 0·00228 | 0·00199 | 0·00114 | 0·00102 | 0·00077 | 0·00228 |
| 520 | $2\times10^{7}$ | 0·00350 | 0·00417 | 0·04391 | 0·03345 | 0·02296 | 0·01341 | 0·01113 | 0·00937 | 0·04391 |

| Boost fold change Ogawa OSP IgG | | | | | | | | | |
|---|---|---|---|---|---|---|---|---|---|
| Dose (CFU) | D1 | D4 | D7 | D15 | D29 | D57 | D85 | D180 | max FC |
| $2\times10^{7}$ | 1·00 | 0·77 | 15·57 | 32·53 | 8·33 | 17·39 | 2·03 | 0·62 | 32·53 |
| $2\times10^{7}$ | 1·00 | 1·34 | 7·20 | 20·96 | 7·61 | 2·53 | 1·10 | 1·27 | 20·96 |
| $2\times10^{7}$ | 1·00 | 0·73 | 1·38 | 1·69 | 1·72 | 1·54 | 0·50 | 0·42 | 1·72 |
| $2\times10^{7}$ | 1·00 | 0·93 | 1·16 | 1·72 | 1·49 | 5·66 | 1·16 | 0·60 | 5·66 |
| $2\times10^{7}$ | 1·00 | 0·95 | 1·61 | 4·41 | 2·77 | 1·97 | 0·91 | 0·56 | 4·41 |
| $2\times10^{7}$ | 1·00 | 0·87 | 0·83 | 0·91 | 0·89 | 0·86 | 0·77 | 0·73 | 0·91 |
| $2\times10^{7}$ | 1·00 | 0·83 | 15·47 | 164·01 | 82·54 | 49·92 | 24·18 | 2·07 | 164·01 |
| $2\times10^{7}$ | 1·00 | 1·23 | 126·78 | 80·73 | 35·65 | 11·86 | 4·13 | 1·08 | 126·78 |
| $2\times10^{7}$ | 1·00 | 1·13 | 0·97 | 1·08 | 0·85 | 0·78 | 0·78 | 0·88 | 1·13 |
| $2\times10^{7}$ | 1·00 | 1·09 | 3·35 | 7·27 | 7·30 | 2·80 | 2·16 | 1·73 | 7·30 |
| $2\times10^{7}$ | 1·00 | 1·03 | 1·90 | 3·21 | 2·53 | 1·70 | 2·01 | 0·62 | 3·21 |
| $2\times10^{7}$ | 1·00 | 0·66 | 1·85 | 1·16 | 1·35 | 0·88 | 0·97 | 0·76 | 1·85 |
| $2\times10^{7}$ | 1·00 | 1·02 | 23·17 | 147·03 | 40·60 | 21·90 | 16·86 | 1·84 | 147·03 |
| $2\times10^{7}$ | 1·00 | 1·18 | 34·72 | 23·69 | 18·37 | | | 2·40 | 34·72 |
| $2\times10^{7}$ | 1·00 | 0·79 | 22·96 | 36·22 | 26·21 | 12·41 | 7·38 | 6·82 | 36·22 |
| $2\times10^{7}$ | 1·00 | 1·06 | 16·22 | 13·80 | 10·32 | 7·88 | 5·09 | 4·30 | 16·22 |
| $2\times10^{7}$ | 1·00 | 0·87 | 5·96 | 6·22 | 3·67 | 2·00 | 1·35 | 2·95 | 6·22 |
| $2\times10^{7}$ | 1·00 | 1·57 | 0·98 | 0·90 | 0·81 | 0·81 | 2·90 | 2·76 | 2·90 |
| $2\times10^{7}$ | 1·00 | 0·83 | 2·80 | 4·45 | 3·88 | 2·23 | 1·99 | 1·50 | 4·45 |
| $2\times10^{7}$ | 1·00 | 1·19 | 12·55 | 9·56 | 6·56 | 3·83 | 3·18 | 2·68 | 12·55 |

| Primary Ogawa OSP IgG (RAU) | | | | | | | | | | |
|---|---|---|---|---|---|---|---|---|---|---|
| Dose interval (days) | Dose (CFU) | D1 | D4 | D7 | D15 | D29 | D57 | D85 | D180 | max |
| 389 | $10^{8}$ | 0·00408 | 0·00468 | 0·00452 | 0·01236 | 0·00327 | 0·00625 | 0·00894 | 0·00484 | 0·01236 |
| 405 | $10^{8}$ | 0·00127 | 0·00069 | 0·00062 | 0·00211 | 0·00029 | 0·00045 | | 0·00008 | 0·00211 |
| 322 | $10^{7}$ | | 0·00004 | 0·00049 | 0·00056 | 0·00016 | 0·00015 | 0·00016 | 0·00016 | 0·00056 |
| 418 | $10^{7}$ | 0·00590 | 0·00600 | 0·00571 | 0·00542 | 0·00527 | 0·00101 | 0·00132 | 0·00322 | 0·00600 |
| 378 | $10^{8}$ | 0·00085 | 0·00108 | 0·00097 | 0·01293 | 0·01229 | 0·00586 | 0·00311 | 0·00184 | 0·01293 |
| 399 | $10^{8}$ | 0·00293 | 0·00244 | 0·00283 | 0·00301 | 0·00151 | 0·00097 | 0·00433 | 0·02118 | 0·02118 |
| 568 | $10^{7}$ | 0·00403 | 0·00324 | 0·00726 | 0·18673 | 0·15346 | 0·10516 | 0·06851 | 0·01925 | 0·18673 |
| 367 | $10^{8}$ | 0·00020 | | 0·00028 | 0·00276 | 0·00193 | 0·00709 | 0·02478 | 0·05072 | 0·05072 |
| 503 | $10^{7}$ | 0·00423 | 0·00439 | 0·00355 | 0·03180 | 0·01977 | 0·01358 | 0·01093 | 0·00389 | 0·03180 |
| 501 | $10^{8}$ | 0·00355 | 0·00318 | 0·00326 | 0·00321 | 0·00345 | 0·00701 | 0·02209 | 0·02314 | 0·02314 |
| 751 | $10^{10}$ | 0·00134 | 0·00134 | 0·00084 | 0·00140 | 0·00129 | 0·00192 | 0·00182 | 0·00258 | 0·00258 |
| 546 | $10^{8}$ | 0·00452 | 0·00397 | 0·00403 | 0·00700 | 0·00710 | 0·00954 | 0·00721 | 0·01552 | 0·01552 |
| 945 | $10^{8}$ | 0·00131 | 0·00119 | 0·00105 | 0·00144 | 0·00138 | 0·00119 | 0·00251 | 0·00616 | 0·00616 |
| 802 | $10^{10}$ | 0·00025 | 0·00067 | 0·00084 | 0·00140 | | | 0·00049 | 0·00055 | 0·00140 |
| 960 | $10^{8}$ | 0·00078 | 0·00092 | 0·00105 | 0·00560 | 0·00239 | 0·00228 | 0·00156 | 0·00187 | 0·00560 |
| 375 | $10^{5}$ | 0·00001 | | 0·00035 | 0·00030 | 0·00042 | 0·00030 | 0·00057 | 0·00264 | 0·00264 |
| 350 | $10^{5}$ | 0·00032 | 0·00049 | 0·00054 | 0·00470 | 0·00259 | 0·00064 | 0·00133 | 0·00502 | 0·00502 |
| 1111 | $10^{6}$ | 0·00105 | 0·00071 | 0·00028 | 0·00119 | 0·00035 | 0·00085 | 0·00057 | 0·00092 | 0·00119 |
| 576 | $10^{7}$ | 0·00170 | 0·00154 | 0·00144 | 0·00215 | 0·00244 | 0·00008 | | 0·00001 | 0·00244 |
| 520 | $10^{7}$ | 0·00064 | 0·00030 | 0·00036 | 0·00006 | 0·00089 | 0·00077 | 0·00104 | 0·00434 | 0·00434 |

| Primary fold change Ogawa OSP IgG | | | | | | | | | |
|---|---|---|---|---|---|---|---|---|---|
| Dose (CFU) | D1 | D4 | D7 | D15 | D29 | D57 | D85 | D180 | max FC |
| $10^{8}$ | 1·00 | 1·15 | 1·11 | 3·03 | 0·80 | 1·53 | 2·19 | 1·19 | 3·03 |
| $10^{8}$ | 1·00 | 0·54 | 0·49 | 1·66 | 0·23 | 0·35 | | 0·06 | 1·66 |
| $10^{7}$ | | 1·00 | 12·25 | 14·00 | 4·00 | 3·75 | 4·00 | 4·00 | 14·00 |
| $10^{7}$ | 1·00 | 1·02 | 0·97 | 0·92 | 0·89 | 0·17 | 0·22 | 0·55 | 1·02 |
| $10^{8}$ | 1·00 | 1·27 | 1·14 | 15·21 | 14·46 | 6·89 | 3·66 | 2·16 | 15·21 |
| $10^{8}$ | 1·00 | 0·83 | 0·97 | 1·03 | 0·52 | 0·33 | 1·48 | 7·23 | 7·23 |
| $10^{7}$ | 1·00 | 0·80 | 1·80 | 46·33 | 38·08 | 26·09 | 17·00 | 4·78 | 46·33 |
| $10^{8}$ | 1·00 | | 1·40 | 13·80 | 9·65 | 35·45 | 123·90 | 253·60 | 253·60 |
| $10^{7}$ | 1·00 | 1·04 | 0·84 | 7·52 | 4·67 | 3·21 | 2·58 | 0·92 | 7·52 |
| $10^{8}$ | 1·00 | 0·90 | 0·92 | 0·90 | 0·97 | 1·97 | 6·22 | 6·52 | 6·52 |
| $10^{10}$ | 1·00 | 1·00 | 0·63 | 1·04 | 0·96 | 1·43 | 1·36 | 1·93 | 1·93 |
| $10^{8}$ | 1·00 | 0·88 | 0·89 | 1·55 | 1·57 | 2·11 | 1·60 | 3·43 | 3·43 |
| $10^{8}$ | 1·00 | 0·91 | 0·80 | 1·10 | 1·05 | 0·91 | 1·92 | 4·70 | 4·70 |
| $10^{10}$ | 1·00 | 2·68 | 3·36 | 5·60 | | | 1·96 | 2·20 | 5·60 |
| $10^{8}$ | 1·00 | 1·18 | 1·35 | 7·18 | 3·06 | 2·92 | 2·00 | 2·40 | 7·18 |
| $10^{5}$ | 1·00 | | 35·00 | 30·00 | 42·00 | 30·00 | 57·00 | 264·03 | 264·03 |
| $10^{5}$ | 1·00 | 1·53 | 1·69 | 14·69 | 8·09 | 2·00 | 4·16 | 15·69 | 15·69 |
| $10^{6}$ | 1·00 | 0·68 | 0·27 | 1·13 | 0·33 | 0·81 | 0·54 | 0·88 | 1·13 |
| $10^{7}$ | 1·00 | 0·91 | 0·85 | 1·26 | 1·44 | 0·05 | | 0·01 | 1·44 |
| $10^{7}$ | 1·00 | 0·47 | 0·56 | 0·09 | 1·39 | 1·20 | 1·62 | 6·78 | 6·78 |

| Boost Ogawa OSP IgA (RAU) | | | | | | | | | | |
|---|---|---|---|---|---|---|---|---|---|---|
| Dose interval (days) | Dose (CFU) | D1 | D4 | D7 | D15 | D29 | D57 | D85 | D180 | max |
| 389 | $2\times10^7$ | 0·00715 | 0·00564 | 0·41080 | 0·18038 | 0·00682 | 0·01701 | 0·00684 | 0·00979 | 0·41080 |
| 405 | $2\times10^7$ | 0·00302 | 0·00297 | 0·01447 | 0·09454 | 0·01255 | 0·00398 | 0·00457 | 0·00473 | 0·09454 |
| 322 | $2\times10^7$ | 0·00249 | 0·00213 | 0·01108 | 0·01283 | 0·00730 | 0·00272 | 0·00377 | 0·00327 | 0·01283 |
| 418 | $2\times10^7$ | 0·00395 | 0·00376 | 0·01166 | 0·01311 | 0·00648 | 0·00513 | 0·00691 | 0·00552 | 0·01311 |
| 378 | $2\times10^7$ | 0·01699 | 0·01729 | 0·14816 | 0·29565 | 0·06965 | 0·02877 | 0·02406 | 0·02484 | 0·29565 |
| 399 | $2\times10^7$ | 0·00721 | 0·00628 | 0·00663 | 0·00514 | 0·00734 | 0·00991 | 0·00897 | 0·00896 | 0·00991 |
| 568 | $2\times10^7$ | 0·00416 | 0·00329 | 0·02027 | 0·05889 | 0·01391 | 0·00627 | 0·00460 | 0·00390 | 0·05889 |
| 367 | $2\times10^7$ | 0·00332 | 0·00312 | 0·06666 | 0·02841 | 0·00587 | 0·00524 | 0·00405 | 0·00419 | 0·06666 |
| 503 | $2\times10^7$ | 0·00525 | 0·00620 | 0·00515 | 0·00560 | 0·00487 | 0·00719 | 0·00778 | 0·01340 | 0·01340 |
| 501 | $2\times10^7$ | 0·00531 | 0·00445 | 0·06023 | 0·03449 | 0·01379 | 0·00833 | 0·00723 | 0·00956 | 0·06023 |
| 751 | $2\times10^7$ | 0·00756 | 0·00680 | 0·08260 | 0·08853 | 0·01997 | 0·00979 | 0·01026 | 0·00913 | 0·08853 |
| 546 | $2\times10^7$ | 0·01150 | 0·01071 | 0·01736 | 0·01195 | 0·01033 | 0·00859 | 0·00837 | 0·01174 | 0·01736 |
| 945 | $2\times10^7$ | 0·00872 | 0·00849 | 0·14873 | 0·19577 | 0·04227 | 0·01536 | 0·01476 | 0·02118 | 0·19577 |
| 802 | $2\times10^7$ | 0·01013 | 0·01137 | 0·45834 | 0·16368 | 0·06617 | | | 0·02318 | 0·45834 |
| 960 | $2\times10^7$ | 0·00727 | 0·00785 | 0·14860 | 0·11199 | 0·03447 | 0·01189 | 0·00952 | 0·01227 | 0·14860 |
| 375 | $2\times10^7$ | 0·01025 | 0·00860 | 0·19076 | 0·11340 | 0·03967 | 0·01526 | 0·01211 | 0·02749 | 0·19076 |
| 350 | $2\times10^7$ | 0·00888 | 0·00856 | 0·07158 | 0·04522 | 0·01610 | 0·00992 | 0·00897 | 0·01719 | 0·07158 |
| 1111 | $2\times10^7$ | 0·00714 | 0·00806 | 0·00957 | 0·00639 | 0·00634 | 0·00644 | 0·01291 | 0·01337 | 0·01337 |
| 576 | $2\times10^7$ | 0·00362 | 0·00429 | 0·02552 | 0·00997 | 0·00599 | 0·00469 | 0·00452 | 0·00493 | 0·02552 |
| 520 | $2\times10^7$ | 0·00649 | 0·00742 | 0·07543 | 0·03883 | 0·01173 | 0·00834 | 0·01376 | 0·01076 | 0·07543 |

| Boost fold change Ogawa OSP IgA | | | | | | | | | |
|---|---|---|---|---|---|---|---|---|---|
| Dose (CFU) | D1 | D4 | D7 | D15 | D29 | D57 | D85 | D180 | max FC |
| $2\times10^7$ | 1·00 | 0·79 | 57·49 | 25·24 | 0·95 | 2·38 | 0·96 | 1·37 | 57·49 |
| $2\times10^7$ | 1·00 | 0·98 | 4·80 | 31·32 | 4·16 | 1·32 | 1·51 | 1·57 | 31·32 |
| $2\times10^7$ | 1·00 | 0·85 | 4·45 | 5·15 | 2·93 | 1·09 | 1·51 | 1·31 | 5·15 |
| $2\times10^7$ | 1·00 | 0·95 | 2·95 | 3·32 | 1·64 | 1·30 | 1·75 | 1·40 | 3·32 |
| $2\times10^7$ | 1·00 | 1·02 | 8·72 | 17·40 | 4·10 | 1·69 | 1·42 | 1·46 | 17·40 |
| $2\times10^7$ | 1·00 | 0·87 | 0·92 | 0·71 | 1·02 | 1·37 | 1·24 | 1·24 | 1·37 |
| $2\times10^7$ | 1·00 | 0·79 | 4·87 | 14·16 | 3·35 | 1·51 | 1·11 | 0·94 | 14·16 |
| $2\times10^7$ | 1·00 | 0·94 | 20·08 | 8·56 | 1·77 | 1·58 | 1·22 | 1·26 | 20·08 |
| $2\times10^7$ | 1·00 | 1·18 | 0·98 | 1·07 | 0·93 | 1·37 | 1·48 | 2·56 | 2·56 |
| $2\times10^7$ | 1·00 | 0·84 | 11·35 | 6·50 | 2·60 | 1·57 | 1·36 | 1·80 | 11·35 |
| $2\times10^7$ | 1·00 | 0·90 | 10·92 | 11·71 | 2·64 | 1·29 | 1·36 | 1·21 | 11·71 |
| $2\times10^7$ | 1·00 | 0·93 | 1·51 | 1·04 | 0·90 | 0·75 | 0·73 | 1·02 | 1·51 |
| $2\times10^7$ | 1·00 | 0·97 | 17·05 | 22·45 | 4·85 | 1·76 | 1·69 | 2·43 | 22·45 |
| $2\times10^7$ | 1·00 | 1·12 | 45·25 | 16·16 | 6·53 | | | 2·29 | 45·25 |
| $2\times10^7$ | 1·00 | 1·08 | 20·44 | 15·40 | 4·74 | 1·63 | 1·31 | 1·69 | 20·44 |
| $2\times10^7$ | 1·00 | 0·84 | 18·61 | 11·06 | 3·87 | 1·49 | 1·18 | 2·68 | 18·61 |
| $2\times10^7$ | 1·00 | 0·96 | 8·06 | 5·09 | 1·81 | 1·12 | 1·01 | 1·94 | 8·06 |
| $2\times10^7$ | 1·00 | 1·13 | 1·34 | 0·89 | 0·89 | 0·90 | 1·81 | 1·87 | 1·87 |
| $2\times10^7$ | 1·00 | 1·19 | 7·05 | 2·76 | 1·66 | 1·30 | 1·25 | 1·36 | 7·05 |
| $2\times10^7$ | 1·00 | 1·14 | 11·63 | 5·99 | 1·81 | 1·29 | 2·12 | 1·66 | 11·63 |

| Primary Ogawa OSP IgA (RAU) | | | | | | | | | | |
|---|---|---|---|---|---|---|---|---|---|---|
| Dose interval (days) | Dose (CFU) | D1 | D4 | D7 | D15 | D29 | D57 | D85 | D180 | max |
| 389 | $10^8$ | 0·00149 | 0·00135 | 0·00160 | 0·01561 | 0·00239 | 0·00198 | 0·00201 | 0·00155 | 0·01561 |
| 405 | $10^8$ | 0·00170 | 0·00126 | 0·00141 | 0·00272 | 0·00081 | 0·00066 | 0·00103 | 0·00107 | 0·00272 |
| 322 | $10^7$ | 0·00023 | 0·00012 | 0·00051 | 0·00130 | 0·00104 | 0·00052 | 0·00062 | 0·00008 | 0·00130 |
| 418 | $10^7$ | 0·00180 | 0·00201 | 0·00210 | 0·00217 | 0·00233 | 0·00220 | 0·00231 | 0·00255 | 0·00255 |
| 378 | $10^8$ | 0·00212 | 0·00232 | 0·13754 | 2·26371 | 0·75668 | 0·04771 | 0·02122 | 0·01139 | 2·26371 |
| 399 | $10^8$ | 0·00388 | 0·00413 | 0·01759 | 0·07248 | 0·02783 | 0·01112 | 0·00821 | 0·00548 | 0·07248 |
| 568 | $10^7$ | 0·00195 | 0·00251 | 0·00330 | 0·24397 | 0·06934 | 0·02000 | 0·01342 | 0·00554 | 0·24397 |
| 367 | $10^8$ | 0·00086 | 0·00082 | 0·00170 | 0·02312 | 0·01066 | 0·00606 | 0·00284 | 0·00115 | 0·02312 |
| 503 | $10^7$ | 0·00201 | 0·00188 | 0·00695 | 0·16153 | 0·02724 | 0·00582 | 0·00526 | 0·00365 | 0·16153 |
| 501 | $10^8$ | 0·00079 | 0·00118 | 0·00108 | 0·00266 | 0·00192 | 0·00237 | 0·00200 | 0·00179 | 0·00266 |
| 751 | $10^{10}$ | 0·00009 | 0·00016 | 0·00103 | 0·01074 | 0·00256 | 0·00117 | 0·00168 | 0·00307 | 0·01074 |
| 546 | $10^8$ | 0·00309 | 0·00257 | 0·03772 | 0·05365 | 0·02586 | 0·00842 | 0·00426 | 0·00286 | 0·05365 |
| 945 | $10^8$ | 0·00154 | 0·00175 | 0·00144 | 0·00722 | 0·00329 | 0·00252 | 0·00265 | 0·00174 | 0·00722 |
| 802 | $10^{10}$ | 0·00101 | 0·00108 | 0·00126 | 0·00417 | | | 0·00233 | 0·00136 | 0·00417 |
| 960 | $10^8$ | 0·00133 | 0·00118 | 0·00203 | 0·05481 | 0·00898 | 0·00212 | 0·00158 | 0·00133 | 0·05481 |
| 375 | $10^5$ | 0·00260 | 0·00242 | 0·00273 | 0·00231 | 0·00269 | 0·00233 | 0·00260 | 0·01050 | 0·01050 |
| 350 | $10^5$ | 0·00230 | 0·00220 | 0·00230 | 0·00450 | 0·00209 | 0·00195 | 0·00200 | 0·00676 | 0·00676 |
| 1111 | $10^6$ | 0·00478 | 0·00285 | 0·00548 | 0·00754 | 0·00395 | 0·00305 | 0·00278 | 0·00233 | 0·00754 |
| 576 | $10^7$ | 0·00180 | 0·00305 | 0·01172 | 0·03785 | 0·01655 | 0·00450 | 0·00327 | 0·00188 | 0·03785 |
| 520 | $10^7$ | 0·00136 | 0·00152 | 0·00268 | 0·00120 | 0·00196 | 0·00278 | 0·00388 | 0·00360 | 0·00388 |

| Primary fold change Ogawa OSP IgA | | | | | | | | | |
|---|---|---|---|---|---|---|---|---|---|
| Dose (CFU) | D1 | D4 | D7 | D15 | D29 | D57 | D85 | D180 | max FC |
| $10^8$ | 1·00 | 0·91 | 1·07 | 10·48 | 1·60 | 1·33 | 1·35 | 1·04 | 10·48 |
| $10^8$ | 1·00 | 0·74 | 0·83 | 1·60 | 0·48 | 0·39 | 0·61 | 0·63 | 1·60 |
| $10^7$ | 1·00 | 0·52 | 2·22 | 5·65 | 4·52 | 2·26 | 2·70 | 0·35 | 5·65 |
| $10^7$ | 1·00 | 1·12 | 1·17 | 1·21 | 1·29 | 1·22 | 1·28 | 1·42 | 1·42 |
| $10^8$ | 1·00 | 1·09 | 64·88 | 1067·79 | 356·92 | 22·50 | 10·01 | 5·37 | 1067·79 |
| $10^8$ | 1·00 | 1·06 | 4·53 | 18·68 | 7·17 | 2·87 | 2·12 | 1·41 | 18·68 |
| $10^7$ | 1·00 | 1·29 | 1·69 | 125·11 | 35·56 | 10·26 | 6·88 | 2·84 | 125·11 |
| $10^8$ | 1·00 | 0·95 | 1·98 | 26·88 | 12·40 | 7·05 | 3·30 | 1·34 | 26·88 |
| $10^7$ | 1·00 | 0·94 | 3·46 | 80·36 | 13·55 | 2·90 | 2·62 | 1·82 | 80·36 |
| $10^8$ | 1·00 | 1·49 | 1·37 | 3·37 | 2·43 | 3·00 | 2·53 | 2·27 | 3·37 |
| $10^{10}$ | 1·00 | 1·78 | 11·44 | 119·33 | 28·44 | 13·00 | 18·67 | 34·11 | 119·33 |
| $10^8$ | 1·00 | 0·83 | 12·21 | 17·36 | 8·37 | 2·72 | 1·38 | 0·93 | 17·36 |
| $10^8$ | 1·00 | 1·14 | 0·94 | 4·69 | 2·14 | 1·64 | 1·72 | 1·13 | 4·69 |
| $10^{10}$ | 1·00 | 1·07 | 1·25 | 4·13 | | | 2·31 | 1·35 | 4·13 |
| $10^8$ | 1·00 | 0·89 | 1·53 | 41·21 | 6·75 | 1·59 | 1·19 | 1·00 | 41·21 |
| $10^5$ | 1·00 | 0·93 | 1·05 | 0·89 | 1·03 | 0·90 | 1·00 | 4·04 | 4·04 |
| $10^5$ | 1·00 | 0·96 | 1·00 | 1·96 | 0·91 | 0·85 | 0·87 | 2·94 | 2·94 |
| $10^6$ | 1·00 | 0·60 | 1·15 | 1·58 | 0·83 | 0·64 | 0·58 | 0·49 | 1·58 |
| $10^7$ | 1·00 | 1·69 | 6·51 | 21·03 | 9·19 | 2·50 | 1·82 | 1·04 | 21·03 |
| $10^7$ | 1·00 | 1·12 | 1·97 | 0·88 | 1·44 | 2·04 | 2·85 | 2·65 | 2·85 |

| Boost CT-B IgM (RAU) | | | | | | | | | | |
|---|---|---|---|---|---|---|---|---|---|---|
| Dose interval (days) | Dose (CFU) | D1 | D4 | D7 | D15 | D29 | D57 | D85 | D180 | max |
| 389 | $2\times10^7$ | 1·03739 | 1·19064 | 1·43408 | 0·91723 | 0·70165 | 0·69119 | 0·55240 | 0·58591 | 1·43408 |
| 405 | $2\times10^7$ | 0·16936 | 0·17489 | 0·23236 | 1·10771 | 0·36618 | 0·17326 | 0·20675 | 0·19905 | 1·10771 |
| 322 | $2\times10^7$ | 6·02579 | 6·61662 | 5·51308 | 5·21683 | 5·89714 | 6·35438 | 8·52581 | 8·65175 | 8·65175 |
| 418 | $2\times10^7$ | 0·27767 | 0·27845 | 0·28057 | 0·25695 | 0·27309 | 0·30942 | 0·50035 | 0·35443 | 0·50035 |
| 378 | $2\times10^7$ | 0·18636 | 0·20667 | 0·19783 | 0·19104 | 0·20456 | 0·19261 | 0·23945 | 0·21868 | 0·23945 |
| 399 | $2\times10^7$ | 0·21715 | 0·19104 | 0·17874 | 0·19382 | 0·22013 | 0·17742 | 0·19195 | 0·19325 | 0·22013 |
| 568 | $2\times10^7$ | 0·40492 | 0·33964 | 0·40721 | 0·41453 | 0·40533 | 0·40268 | 0·32783 | 0·38887 | 0·41453 |
| 367 | $2\times10^7$ | 0·67825 | 0·76769 | 0·86676 | 0·71588 | 0·83100 | 0·91104 | 1·10401 | 1·00716 | 1·10401 |
| 503 | $2\times10^7$ | 0·60903 | 0·61471 | 0·60862 | 0·59995 | 0·57088 | 0·59285 | 0·57041 | 0·51246 | 0·61471 |
| 501 | $2\times10^7$ | 0·37235 | 0·34997 | 0·30293 | 0·31378 | 0·32367 | 0·43029 | 0·47048 | 0·86470 | 0·86470 |
| 751 | $2\times10^7$ | 0·75400 | 0·88735 | 0·86670 | 0·89115 | 0·82179 | 0·76395 | 0·88678 | 1·03062 | 1·03062 |
| 546 | $2\times10^7$ | 0·41955 | 0·45362 | 0·37177 | 0·38223 | 0·35298 | 0·35871 | 0·33912 | 0·42371 | 0·45362 |
| 945 | $2\times10^7$ | 0·38023 | 0·34236 | 0·38313 | 0·37264 | 0·36149 | 0·38123 | 0·42467 | 0·24335 | 0·42467 |
| 802 | $2\times10^7$ | 0·34366 | 0·34924 | 0·31054 | 0·35459 | 0·36660 | | | 0·27517 | 0·36660 |
| 960 | $2\times10^7$ | 0·70849 | 0·61720 | 0·80730 | 13·97742 | 12·04183 | 3·57727 | 2·01608 | 0·68418 | 13·97742 |
| 375 | $2\times10^7$ | 0·47757 | 0·41334 | 0·67544 | 0·57535 | 0·51190 | 0·66177 | 0·51668 | 0·49993 | 0·67544 |
| 350 | $2\times10^7$ | 0·75435 | 0·78285 | 0·76999 | 0·70849 | 0·74592 | 0·67595 | 0·68245 | 0·92422 | 0·92422 |
| 1111 | $2\times10^7$ | 6·08685 | 9·64602 | 7·55574 | 7·11440 | 4·65524 | 5·40654 | 4·96146 | 3·33096 | 9·64602 |
| 576 | $2\times10^7$ | 1·19481 | 1·21558 | 1·43162 | 1·01571 | 1·26495 | 1·15264 | 1·62050 | 1·36859 | 1·62050 |
| 520 | $2\times10^7$ | 2·35531 | 2·68713 | 2·73759 | 2·18203 | 2·64126 | 2·74288 | 4·17388 | 3·67335 | 4·17388 |

| Boost fold change CT-B IgM | | | | | | | | | |
|---|---|---|---|---|---|---|---|---|---|
| Dose (CFU) | D1 | D4 | D7 | D15 | D29 | D57 | D85 | D180 | max FC |
| $2\times10^7$ | 1·00 | 1·15 | 1·38 | 0·88 | 0·68 | 0·67 | 0·53 | 0·56 | 1·38 |
| $2\times10^7$ | 1·00 | 1·03 | 1·37 | 6·54 | 2·16 | 1·02 | 1·22 | 1·18 | 6·54 |
| $2\times10^7$ | 1·00 | 1·10 | 0·91 | 0·87 | 0·98 | 1·05 | 1·41 | 1·44 | 1·44 |
| $2\times10^7$ | 1·00 | 1·00 | 1·01 | 0·93 | 0·98 | 1·11 | 1·80 | 1·28 | 1·80 |
| $2\times10^7$ | 1·00 | 1·11 | 1·06 | 1·03 | 1·10 | 1·03 | 1·28 | 1·17 | 1·28 |
| $2\times10^7$ | 1·00 | 0·88 | 0·82 | 0·89 | 1·01 | 0·82 | 0·88 | 0·89 | 1·01 |
| $2\times10^7$ | 1·00 | 0·84 | 1·01 | 1·02 | 1·00 | 0·99 | 0·81 | 0·96 | 1·02 |
| $2\times10^7$ | 1·00 | 1·13 | 1·28 | 1·06 | 1·23 | 1·34 | 1·63 | 1·48 | 1·63 |
| $2\times10^7$ | 1·00 | 1·01 | 1·00 | 0·99 | 0·94 | 0·97 | 0·94 | 0·84 | 1·01 |
| $2\times10^7$ | 1·00 | 0·94 | 0·81 | 0·84 | 0·87 | 1·16 | 1·26 | 2·32 | 2·32 |
| $2\times10^7$ | 1·00 | 1·18 | 1·15 | 1·18 | 1·09 | 1·01 | 1·18 | 1·37 | 1·37 |
| $2\times10^7$ | 1·00 | 1·08 | 0·89 | 0·91 | 0·84 | 0·85 | 0·81 | 1·01 | 1·08 |
| $2\times10^7$ | 1·00 | 0·90 | 1·01 | 0·98 | 0·95 | 1·00 | 1·12 | 0·64 | 1·12 |
| $2\times10^7$ | 1·00 | 1·02 | 0·90 | 1·03 | 1·07 | | | 0·80 | 1·07 |
| $2\times10^7$ | 1·00 | 0·87 | 1·14 | 19·73 | 17·00 | 5·05 | 2·85 | 0·97 | 19·73 |
| $2\times10^7$ | 1·00 | 0·87 | 1·41 | 1·20 | 1·07 | 1·39 | 1·08 | 1·05 | 1·41 |
| $2\times10^7$ | 1·00 | 1·04 | 1·02 | 0·94 | 0·99 | 0·90 | 0·90 | 1·23 | 1·23 |
| $2\times10^7$ | 1·00 | 1·58 | 1·24 | 1·17 | 0·76 | 0·89 | 0·82 | 0·55 | 1·58 |
| $2\times10^7$ | 1·00 | 1·02 | 1·20 | 0·85 | 1·06 | 0·96 | 1·36 | 1·15 | 1·36 |
| $2\times10^7$ | 1·00 | 1·14 | 1·16 | 0·93 | 1·12 | 1·16 | 1·77 | 1·56 | 1·77 |

| Primary CT-B IgM (RAU) | | | | | | | | | | |
|---|---|---|---|---|---|---|---|---|---|---|
| Dose interval (days) | Dose (CFU) | D1 | D4 | D7 | D15 | D29 | D57 | D85 | D180 | max |
| 389 | $10^8$ | 0·96334 | 1·18608 | 1·06117 | 2·37578 | 2·77874 | 1·52005 | 1·67208 | 1·18590 | 2·77874 |
| 405 | $10^8$ | 0·27511 | 0·24392 | 0·25143 | 0·22992 | 0·30127 | 0·33581 | 0·46460 | 0·38846 | 0·46460 |
| 322 | $10^7$ | 6·44998 | 7·06733 | 7·67717 | 7·03415 | 7·04519 | 7·11180 | 6·96820 | 7·17896 | 7·67717 |
| 418 | $10^7$ | 0·32641 | 0·37977 | 0·35367 | 0·33007 | 0·33519 | 0·60213 | 0·46752 | 0·48941 | 0·60213 |
| 378 | $10^8$ | 0·32580 | 0·38272 | 0·31469 | 0·27476 | 0·22739 | 0·24538 | 0·26581 | 0·24868 | 0·38272 |
| 399 | $10^8$ | 0·51993 | 0·46006 | 0·48530 | 0·38354 | 0·49144 | 0·42367 | 0·41658 | 0·45310 | 0·51993 |
| 568 | $10^7$ | 0·56271 | 0·55908 | 0·56401 | 0·43040 | 0·41883 | 0·44226 | 0·58814 | 0·43649 | 0·58814 |
| 367 | $10^8$ | 1·25256 | 1·56546 | 1·62095 | 1·70904 | 1·58279 | 1·57410 | 1·63285 | 1·69356 | 1·70904 |
| 503 | $10^7$ | 0·98241 | 0·93126 | 1·13472 | 1·22269 | 1·04226 | 0·84987 | 0·92273 | 0·81421 | 1·22269 |
| 501 | $10^8$ | 0·71517 | 0·71885 | 0·72182 | 0·75208 | 0·65330 | 0·61216 | 0·79833 | 0·75534 | 0·79833 |
| 751 | $10^{10}$ | 1·07689 | 1·01323 | 1·30508 | 2·43296 | 1·14982 | 0·85427 | 0·94011 | 0·80897 | 2·43296 |
| 546 | $10^8$ | 1·17598 | 1·20657 | 1·07723 | 0·64975 | 0·75173 | 0·90996 | 0·88401 | 0·65548 | 1·20657 |
| 945 | $10^8$ | 0·67071 | 0·59575 | 0·47675 | 0·40417 | 0·43163 | 0·48582 | 0·50863 | 0·41777 | 0·67071 |
| 802 | $10^{10}$ | 0·87717 | 1·06467 | 0·70027 | 0·72599 | | | 0·67480 | 0·73569 | 1·06467 |
| 960 | $10^8$ | 0·90076 | 0·82534 | 0·84373 | 0·93846 | 0·82047 | 0·92457 | 0·35076 | 0·37795 | 0·93846 |
| 375 | $10^5$ | 0·43177 | 0·42725 | 0·48235 | 0·48335 | 0·88920 | 0·65285 | 0·85346 | 0·40795 | 0·88920 |
| 350 | $10^5$ | 0·95358 | 0·88401 | 1·08047 | 0·94074 | 0·94989 | 0·88229 | 0·95727 | 0·74169 | 1·08047 |
| 1111 | $10^6$ | 4·16655 | 5·64419 | 6·28863 | 4·18381 | 2·92414 | 4·17635 | 3·66352 | 3·84355 | 6·28863 |
| 576 | $10^7$ | 0·32106 | 0·35431 | 0·35207 | 1·09079 | 0·79098 | 1·13101 | 0·77351 | 4·15786 | 4·15786 |
| 520 | $10^7$ | 3·94900 | 4·01532 | 5·53359 | 3·89665 | 4·04886 | 4·35495 | 4·75105 | 4·62821 | 5·53359 |

| Primary fold change CT-B IgM | | | | | | | | | |
|---|---|---|---|---|---|---|---|---|---|
| Dose (CFU) | D1 | D4 | D7 | D15 | D29 | D57 | D85 | D180 | max FC |
| $10^8$ | 1·00 | 1·23 | 1·10 | 2·47 | 2·88 | 1·58 | 1·74 | 1·23 | 2·88 |
| $10^8$ | 1·00 | 0·89 | 0·91 | 0·84 | 1·10 | 1·22 | 1·69 | 1·41 | 1·69 |
| $10^7$ | 1·00 | 1·10 | 1·19 | 1·09 | 1·09 | 1·10 | 1·08 | 1·11 | 1·19 |
| $10^7$ | 1·00 | 1·16 | 1·08 | 1·01 | 1·03 | 1·84 | 1·43 | 1·50 | 1·84 |
| $10^8$ | 1·00 | 1·17 | 0·97 | 0·84 | 0·70 | 0·75 | 0·82 | 0·76 | 1·17 |
| $10^8$ | 1·00 | 0·88 | 0·93 | 0·74 | 0·95 | 0·81 | 0·80 | 0·87 | 0·95 |
| $10^7$ | 1·00 | 0·99 | 1·00 | 0·76 | 0·74 | 0·79 | 1·05 | 0·78 | 1·05 |
| $10^8$ | 1·00 | 1·25 | 1·29 | 1·36 | 1·26 | 1·26 | 1·30 | 1·35 | 1·36 |
| $10^7$ | 1·00 | 0·95 | 1·16 | 1·24 | 1·06 | 0·87 | 0·94 | 0·83 | 1·24 |
| $10^8$ | 1·00 | 1·01 | 1·01 | 1·05 | 0·91 | 0·86 | 1·12 | 1·06 | 1·12 |
| $10^{10}$ | 1·00 | 0·94 | 1·21 | 2·26 | 1·07 | 0·79 | 0·87 | 0·75 | 2·26 |
| $10^8$ | 1·00 | 1·03 | 0·92 | 0·55 | 0·64 | 0·77 | 0·75 | 0·56 | 1·03 |
| $10^8$ | 1·00 | 0·89 | 0·71 | 0·60 | 0·64 | 0·72 | 0·76 | 0·62 | 0·89 |
| $10^{10}$ | 1·00 | 1·21 | 0·80 | 0·83 | | | 0·77 | 0·84 | 1·21 |
| $10^8$ | 1·00 | 0·92 | 0·94 | 1·04 | 0·91 | 1·03 | 0·39 | 0·42 | 1·04 |
| $10^5$ | 1·00 | 0·99 | 1·12 | 1·12 | 2·06 | 1·51 | 1·98 | 0·94 | 2·06 |
| $10^5$ | 1·00 | 0·93 | 1·13 | 0·99 | 1·00 | 0·93 | 1·00 | 0·78 | 1·13 |
| $10^6$ | 1·00 | 1·35 | 1·51 | 1·00 | 0·70 | 1·00 | 0·88 | 0·92 | 1·51 |
| $10^7$ | 1·00 | 1·10 | 1·10 | 3·40 | 2·46 | 3·52 | 2·41 | 12·95 | 12·95 |
| $10^7$ | 1·00 | 1·02 | 1·40 | 0·99 | 1·03 | 1·10 | 1·20 | 1·17 | 1·40 |

| Boost CT-B IgG (RAU) | | | | | | | | | | |
|---|---|---|---|---|---|---|---|---|---|---|
| Dose interval (days) | Dose (CFU) | D1 | D4 | D7 | D15 | D29 | D57 | D85 | D180 | max |
| 389 | $2\times10^{7}$ | 0·35639 | 0·27398 | 0·44867 | 0·30828 | 0·29358 | 0·36175 | 0·14078 | 0·13232 | 0·44867 |
| 405 | $2\times10^{7}$ | 0·02104 | 0·01968 | 0·02841 | 0·50498 | 0·25677 | 0·09928 | 0·05351 | 0·03551 | 0·50498 |
| 322 | $2\times10^{7}$ | 0·01303 | 0·01169 | 0·01317 | 0·01163 | 0·01284 | 0·01395 | 0·00835 | 0·00748 | 0·01395 |
| 418 | $2\times10^{7}$ | 0·00792 | 0·00723 | 0·00756 | 0·00680 | 0·00702 | 0·00644 | 0·00642 | 0·00444 | 0·00792 |
| 378 | $2\times10^{7}$ | 0·02509 | 0·02689 | 0·02290 | 0·05900 | 0·05733 | 0·05121 | 0·03193 | 0·02278 | 0·05900 |
| 399 | $2\times10^{7}$ | 0·01292 | 0·01085 | 0·01100 | 0·01148 | 0·01234 | 0·00931 | 0·00840 | 0·00819 | 0·01292 |
| 568 | $2\times10^{7}$ | 0·01879 | 0·01664 | 0·03868 | 0·36874 | 0·21516 | 0·25850 | 0·10366 | 0·03558 | 0·36874 |
| 367 | $2\times10^{7}$ | 0·03285 | 0·03443 | 0·07102 | 0·06617 | 0·05737 | 0·03883 | 0·03238 | 0·02936 | 0·07102 |
| 503 | $2\times10^{7}$ | 0·01865 | 0·02025 | 0·01787 | 0·01913 | 0·01765 | 0·01461 | 0·01465 | 0·02805 | 0·02805 |
| 501 | $2\times10^{7}$ | 0·02572 | 0·02684 | 0·02642 | 0·03342 | 0·03918 | 0·02575 | 0·02803 | 0·03686 | 0·03918 |
| 751 | $2\times10^{7}$ | 0·09503 | 0·10374 | 0·10683 | 0·17087 | 0·15454 | 0·13217 | 0·15294 | 0·10759 | 0·17087 |
| 546 | $2\times10^{7}$ | 0·04112 | 0·03208 | 0·03271 | 0·03472 | 0·03885 | 0·03247 | 0·03442 | 0·04694 | 0·04694 |
| 945 | $2\times10^{7}$ | 0·02057 | 0·01944 | 0·03191 | 0·14959 | 0·05380 | 0·05714 | 0·06396 | 0·05177 | 0·14959 |
| 802 | $2\times10^{7}$ | 0·05312 | 0·05269 | 0·09728 | 0·11105 | 0·10705 | | | 0·09621 | 0·11105 |
| 960 | $2\times10^{7}$ | 0·02822 | 0·02639 | 0·09863 | 0·52164 | 0·41575 | 0·26238 | 0·19624 | 0·15116 | 0·52164 |
| 375 | $2\times10^{7}$ | 0·01486 | 0·01373 | 0·12053 | 0·10770 | 0·07627 | 0·05030 | 0·03213 | 0·04843 | 0·12053 |
| 350 | $2\times10^{7}$ | 0·02151 | 0·01750 | 0·01847 | 0·02024 | 0·01916 | 0·01889 | 0·01675 | 0·03394 | 0·03394 |
| 1111 | $2\times10^{7}$ | 0·02215 | 0·02300 | 0·02069 | 0·02061 | 0·01970 | 0·01760 | 0·02011 | 0·01958 | 0·02300 |
| 576 | $2\times10^{7}$ | 0·03480 | 0·03593 | 0·08215 | 0·11754 | 0·11173 | 0·08079 | 0·08854 | 0·06279 | 0·11754 |
| 520 | $2\times10^{7}$ | 0·02405 | 0·02831 | 0·03845 | 0·03472 | 0·03341 | 0·03102 | 0·03525 | 0·02811 | 0·03845 |

| Boost fold change CT-B IgG | | | | | | | | | |
|---|---|---|---|---|---|---|---|---|---|
| Dose (CFU) | D1 | D4 | D7 | D15 | D29 | D57 | D85 | D180 | max FC |
| $2\times10^{7}$ | 1·00 | 0·77 | 1·26 | 0·86 | 0·82 | 1·02 | 0·40 | 0·37 | 1·26 |
| $2\times10^{7}$ | 1·00 | 0·94 | 1·35 | 24·00 | 12·20 | 4·72 | 2·54 | 1·69 | 24·00 |
| $2\times10^{7}$ | 1·00 | 0·90 | 1·01 | 0·89 | 0·98 | 1·07 | 0·64 | 0·57 | 1·07 |
| $2\times10^{7}$ | 1·00 | 0·91 | 0·95 | 0·86 | 0·89 | 0·81 | 0·81 | 0·56 | 0·95 |
| $2\times10^{7}$ | 1·00 | 1·07 | 0·91 | 2·35 | 2·29 | 2·04 | 1·27 | 0·91 | 2·35 |
| $2\times10^{7}$ | 1·00 | 0·84 | 0·85 | 0·89 | 0·96 | 0·72 | 0·65 | 0·63 | 0·96 |
| $2\times10^{7}$ | 1·00 | 0·89 | 2·06 | 19·62 | 11·45 | 13·76 | 5·52 | 1·89 | 19·62 |
| $2\times10^{7}$ | 1·00 | 1·05 | 2·16 | 2·01 | 1·75 | 1·18 | 0·99 | 0·89 | 2·16 |
| $2\times10^{7}$ | 1·00 | 1·09 | 0·96 | 1·03 | 0·95 | 0·78 | 0·79 | 1·50 | 1·50 |
| $2\times10^{7}$ | 1·00 | 1·04 | 1·03 | 1·30 | 1·52 | 1·00 | 1·09 | 1·43 | 1·52 |
| $2\times10^{7}$ | 1·00 | 1·09 | 1·12 | 1·80 | 1·63 | 1·39 | 1·61 | 1·13 | 1·80 |
| $2\times10^{7}$ | 1·00 | 0·78 | 0·80 | 0·84 | 0·94 | 0·79 | 0·84 | 1·14 | 1·14 |
| $2\times10^{7}$ | 1·00 | 0·95 | 1·55 | 7·27 | 2·62 | 2·78 | 3·11 | 2·52 | 7·27 |
| $2\times10^{7}$ | 1·00 | 0·99 | 1·83 | 2·09 | 2·02 | | | 1·81 | 2·09 |
| $2\times10^{7}$ | 1·00 | 0·94 | 3·50 | 18·49 | 14·73 | 9·30 | 6·95 | 5·36 | 18·49 |
| $2\times10^{7}$ | 1·00 | 0·92 | 8·11 | 7·25 | 5·13 | 3·38 | 2·16 | 3·26 | 8·11 |
| $2\times10^{7}$ | 1·00 | 0·81 | 0·86 | 0·94 | 0·89 | 0·88 | 0·78 | 1·58 | 1·58 |
| $2\times10^{7}$ | 1·00 | 1·04 | 0·93 | 0·93 | 0·89 | 0·79 | 0·91 | 0·88 | 1·04 |
| $2\times10^{7}$ | 1·00 | 1·03 | 2·36 | 3·38 | 3·21 | 2·32 | 2·54 | 1·80 | 3·38 |
| $2\times10^{7}$ | 1·00 | 1·18 | 1·60 | 1·44 | 1·39 | 1·29 | 1·47 | 1·17 | 1·60 |

| Primary CT-B IgG (RAU) | | | | | | | | | | |
|---|---|---|---|---|---|---|---|---|---|---|
| Dose interval (days) | Dose (CFU) | D1 | D4 | D7 | D15 | D29 | D57 | D85 | D180 | max |
| 389 | $10^{8}$ | 0·06990 | 0·08113 | 0·11032 | 3·68765 | 3·89655 | 3·46795 | 1·76769 | 0·19164 | 3·89655 |
| 405 | $10^{8}$ | 0·00700 | 0·00573 | 0·00662 | 0·01361 | 0·02172 | 0·02038 | 0·02489 | 0·01916 | 0·02489 |
| 322 | $10^{7}$ | 0·00412 | 0·00526 | 0·00512 | 0·00488 | 0·00577 | 0·00539 | 0·00516 | 0·00577 | 0·00577 |
| 418 | $10^{7}$ | 0·00452 | 0·00457 | 0·00434 | 0·00481 | 0·00755 | 0·00584 | 0·00480 | 0·00463 | 0·00755 |
| 378 | $10^{8}$ | 0·01108 | 0·01329 | 0·03141 | 0·32014 | 0·18031 | 0·04703 | 0·02616 | 0·01984 | 0·32014 |
| 399 | $10^{8}$ | 0·00327 | 0·00243 | 0·00240 | 0·00396 | 0·01132 | 0·01535 | 0·01436 | 0·01348 | 0·01535 |
| 568 | $10^{7}$ | 0·01626 | 0·01855 | 0·02626 | 0·27783 | 0·17611 | 0·12094 | 0·08473 | 0·04446 | 0·27783 |
| 367 | $10^{8}$ | 0·02612 | 0·02495 | 0·02437 | 0·02646 | 0·02605 | 0·02694 | 0·02644 | 0·02891 | 0·02891 |
| 503 | $10^{7}$ | 0·01200 | 0·01141 | 0·01027 | 0·03015 | 0·03656 | 0·03686 | 0·03377 | 0·01353 | 0·03686 |
| 501 | $10^{8}$ | 0·03011 | 0·03274 | 0·05401 | 0·08919 | 0·06974 | 0·05313 | 0·02221 | 0·02277 | 0·08919 |
| 751 | $10^{10}$ | 0·01006 | 0·01022 | 0·02122 | 1·67337 | 1·23423 | 0·82762 | 0·49335 | 0·26583 | 1·67337 |
| 546 | $10^{8}$ | 0·02948 | 0·02736 | 0·04662 | 0·34480 | 0·32304 | 0·17476 | 0·09586 | 0·03630 | 0·34480 |
| 945 | $10^{8}$ | 0·01487 | 0·01438 | 0·01523 | 0·04715 | 0·07052 | 0·05336 | 0·07113 | 0·04438 | 0·07113 |
| 802 | $10^{10}$ | 0·04259 | 0·04580 | 0·04714 | 0·22594 | | | 0·09633 | 0·07051 | 0·22594 |
| 960 | $10^{8}$ | 0·02166 | 0·02025 | 0·02221 | 0·10638 | 0·10757 | 0·10950 | 0·05941 | 0·04704 | 0·10950 |
| 375 | $10^{5}$ | 0·00809 | 0·00862 | 0·01033 | 0·01042 | 0·01077 | 0·01244 | 0·01463 | 0·01373 | 0·01463 |
| 350 | $10^{5}$ | 0·01009 | 0·00964 | 0·00995 | 0·01234 | 0·02839 | 0·02199 | 0·02409 | 0·02284 | 0·02839 |
| 1111 | $10^{6}$ | 0·02036 | 0·02579 | 0·03829 | 0·20417 | 0·11996 | 0·07454 | 0·04340 | 0·03820 | 0·20417 |
| 576 | $10^{7}$ | 0·02336 | 0·02500 | 0·03214 | 0·80040 | 0·64070 | 0·33133 | 0·12467 | 0·04366 | 0·80040 |
| 520 | $10^{7}$ | 0·01236 | 0·01224 | 0·01628 | 0·03996 | 0·04890 | 0·03307 | 0·03532 | 0·02204 | 0·04890 |

| Primary fold change CT-B IgG | | | | | | | | | |
|---|---|---|---|---|---|---|---|---|---|
| Dose (CFU) | D1 | D4 | D7 | D15 | D29 | D57 | D85 | D180 | max FC |
| $10^{8}$ | 1·00 | 1·16 | 1·58 | 52·76 | 55·74 | 49·61 | 25·29 | 2·74 | 55·74 |
| $10^{8}$ | 1·00 | 0·82 | 0·95 | 1·94 | 3·10 | 2·91 | 3·56 | 2·74 | 3·56 |
| $10^{7}$ | 1·00 | 1·28 | 1·24 | 1·18 | 1·40 | 1·31 | 1·25 | 1·40 | 1·40 |
| $10^{7}$ | 1·00 | 1·01 | 0·96 | 1·06 | 1·67 | 1·29 | 1·06 | 1·02 | 1·67 |
| $10^{8}$ | 1·00 | 1·20 | 2·83 | 28·89 | 16·27 | 4·24 | 2·36 | 1·79 | 28·89 |
| $10^{8}$ | 1·00 | 0·74 | 0·73 | 1·21 | 3·46 | 4·69 | 4·39 | 4·12 | 4·69 |
| $10^{7}$ | 1·00 | 1·14 | 1·62 | 17·09 | 10·83 | 7·44 | 5·21 | 2·73 | 17·09 |
| $10^{8}$ | 1·00 | 0·96 | 0·93 | 1·01 | 1·00 | 1·03 | 1·01 | 1·11 | 1·11 |
| $10^{7}$ | 1·00 | 0·95 | 0·86 | 2·51 | 3·05 | 3·07 | 2·81 | 1·13 | 3·07 |
| $10^{8}$ | 1·00 | 1·09 | 1·79 | 2·96 | 2·32 | 1·76 | 0·74 | 0·76 | 2·96 |
| $10^{10}$ | 1·00 | 1·02 | 2·11 | 166·34 | 122·69 | 82·27 | 49·04 | 26·42 | 166·34 |
| $10^{8}$ | 1·00 | 0·93 | 1·58 | 11·70 | 10·96 | 5·93 | 3·25 | 1·23 | 11·70 |
| $10^{8}$ | 1·00 | 0·97 | 1·02 | 3·17 | 4·74 | 3·59 | 4·78 | 2·98 | 4·78 |
| $10^{10}$ | 1·00 | 1·08 | 1·11 | 5·31 | | | 2·26 | 1·66 | 5·31 |
| $10^{8}$ | 1·00 | 0·93 | 1·03 | 4·91 | 4·97 | 5·06 | 2·74 | 2·17 | 5·06 |
| $10^{5}$ | 1·00 | 1·07 | 1·28 | 1·29 | 1·33 | 1·54 | 1·81 | 1·70 | 1·81 |
| $10^{5}$ | 1·00 | 0·96 | 0·99 | 1·22 | 2·81 | 2·18 | 2·39 | 2·26 | 2·81 |
| $10^{6}$ | 1·00 | 1·27 | 1·88 | 10·03 | 5·89 | 3·66 | 2·13 | 1·88 | 10·03 |
| $10^{7}$ | 1·00 | 1·07 | 1·38 | 34·26 | 27·43 | 14·18 | 5·34 | 1·87 | 34·26 |
| $10^{7}$ | 1·00 | 0·99 | 1·32 | 3·23 | 3·96 | 2·68 | 2·86 | 1·78 | 3·96 |

| Boost CT-B IgA (RAU) | | | | | | | | | | |
|---|---|---|---|---|---|---|---|---|---|---|
| Dose interval (days) | Dose (CFU) | D1 | D4 | D7 | D15 | D29 | D57 | D85 | D180 | max |
| 389 | $2\times10^{7}$ | 0·02240 | 0·02385 | 0·11509 | 0·07999 | 0·02252 | 0·02749 | 0·01781 | 0·01707 | 0·11509 |
| 405 | $2\times10^{7}$ | 0·03166 | 0·02719 | 0·05604 | 0·58326 | 0·10472 | 0·05186 | 0·04990 | 0·04175 | 0·58326 |
| 322 | $2\times10^{7}$ | 0·01986 | 0·01778 | 0·01345 | 0·01253 | 0·02008 | 0·01790 | 0·01551 | 0·01540 | 0·02008 |
| 418 | $2\times10^{7}$ | 0·02528 | 0·02528 | 0·02729 | 0·02810 | 0·02858 | 0·03420 | 0·04909 | 0·02979 | 0·04909 |
| 378 | $2\times10^{7}$ | 0·03626 | 0·03671 | 0·04160 | 0·14978 | 0·06570 | 0·04647 | 0·05045 | 0·06312 | 0·14978 |
| 399 | $2\times10^{7}$ | 0·03274 | 0·02415 | 0·02647 | 0·02845 | 0·03257 | 0·03854 | 0·03980 | 0·03842 | 0·03980 |
| 568 | $2\times10^{7}$ | 0·01811 | 0·01248 | 0·06522 | 0·14330 | 0·03984 | 0·02775 | 0·02098 | 0·01577 | 0·14330 |
| 367 | $2\times10^{7}$ | 0·02512 | 0·02411 | 0·07804 | 0·05554 | 0·03354 | 0·03491 | 0·04522 | 0·03457 | 0·07804 |
| 503 | $2\times10^{7}$ | 0·01338 | 0·01496 | 0·01447 | 0·01625 | 0·01250 | 0·01563 | 0·01614 | 0·02748 | 0·02748 |
| 501 | $2\times10^{7}$ | 0·02186 | 0·02131 | 0·02679 | 0·02426 | 0·03610 | 0·03046 | 0·03349 | 0·03939 | 0·03939 |
| 751 | $2\times10^{7}$ | 0·00895 | 0·00846 | 0·01294 | 0·02572 | 0·01345 | 0·01044 | 0·01164 | 0·01075 | 0·02572 |
| 546 | $2\times10^{7}$ | 0·05331 | 0·04521 | 0·04310 | 0·04616 | 0·04569 | 0·03613 | 0·03691 | 0·04976 | 0·05331 |
| 945 | $2\times10^{7}$ | 0·21079 | 0·19232 | 0·33500 | 0·84468 | 0·58536 | 0·42554 | 0·39124 | 0·40399 | 0·84468 |
| 802 | $2\times10^{7}$ | 0·15088 | 0·15328 | 0·24891 | 0·24377 | 0·19623 | | | 0·14329 | 0·24891 |
| 960 | $2\times10^{7}$ | 0·04862 | 0·04542 | 0·16397 | 0·24837 | 0·12272 | 0·06545 | 0·05629 | 0·07044 | 0·24837 |
| 375 | $2\times10^{7}$ | 0·05108 | 0·04550 | 0·52688 | 0·38149 | 0·16942 | 0·08677 | 0·06308 | 0·10005 | 0·52688 |
| 350 | $2\times10^{7}$ | 0·02557 | 0·02335 | 0·02610 | 0·02592 | 0·02344 | 0·02230 | 0·02214 | 0·03827 | 0·03827 |
| 1111 | $2\times10^{7}$ | 0·13220 | 0·15766 | 0·16894 | 0·12626 | 0·12761 | 0·12137 | 0·14753 | 0·10739 | 0·16894 |
| 576 | $2\times10^{7}$ | 0·04837 | 0·05392 | 0·39431 | 0·24719 | 0·14752 | 0·07671 | 0·08479 | 0·07970 | 0·39431 |
| 520 | $2\times10^{7}$ | 0·06006 | 0·07031 | 0·09723 | 0·07595 | 0·06796 | 0·06838 | 0·08316 | 0·06332 | 0·09723 |

| Boost fold change CT-B IgA | | | | | | | | | |
|---|---|---|---|---|---|---|---|---|---|
| Dose (CFU) | D1 | D4 | D7 | D15 | D29 | D57 | D85 | D180 | max FC |
| $2\times10^{7}$ | 1·00 | 1·06 | 5·14 | 3·57 | 1·01 | 1·23 | 0·79 | 0·76 | 5·14 |
| $2\times10^{7}$ | 1·00 | 0·86 | 1·77 | 18·42 | 3·31 | 1·64 | 1·58 | 1·32 | 18·42 |
| $2\times10^{7}$ | 1·00 | 0·90 | 0·68 | 0·63 | 1·01 | 0·90 | 0·78 | 0·78 | 1·01 |
| $2\times10^{7}$ | 1·00 | 1·00 | 1·08 | 1·11 | 1·13 | 1·35 | 1·94 | 1·18 | 1·94 |
| $2\times10^{7}$ | 1·00 | 1·01 | 1·15 | 4·13 | 1·81 | 1·28 | 1·39 | 1·74 | 4·13 |
| $2\times10^{7}$ | 1·00 | 0·74 | 0·81 | 0·87 | 0·99 | 1·18 | 1·22 | 1·17 | 1·22 |
| $2\times10^{7}$ | 1·00 | 0·69 | 3·60 | 7·91 | 2·20 | 1·53 | 1·16 | 0·87 | 7·91 |
| $2\times10^{7}$ | 1·00 | 0·96 | 3·11 | 2·21 | 1·34 | 1·39 | 1·80 | 1·38 | 3·11 |
| $2\times10^{7}$ | 1·00 | 1·12 | 1·08 | 1·21 | 0·93 | 1·17 | 1·21 | 2·05 | 2·05 |
| $2\times10^{7}$ | 1·00 | 0·97 | 1·23 | 1·11 | 1·65 | 1·39 | 1·53 | 1·80 | 1·80 |
| $2\times10^{7}$ | 1·00 | 0·95 | 1·45 | 2·87 | 1·50 | 1·17 | 1·30 | 1·20 | 2·87 |
| $2\times10^{7}$ | 1·00 | 0·85 | 0·81 | 0·87 | 0·86 | 0·68 | 0·69 | 0·93 | 0·93 |
| $2\times10^{7}$ | 1·00 | 0·91 | 1·59 | 4·01 | 2·78 | 2·02 | 1·86 | 1·92 | 4·01 |
| $2\times10^{7}$ | 1·00 | 1·02 | 1·65 | 1·62 | 1·30 | | | 0·95 | 1·65 |
| $2\times10^{7}$ | 1·00 | 0·93 | 3·37 | 5·11 | 2·52 | 1·35 | 1·16 | 1·45 | 5·11 |
| $2\times10^{7}$ | 1·00 | 0·89 | 10·31 | 7·47 | 3·32 | 1·70 | 1·23 | 1·96 | 10·31 |
| $2\times10^{7}$ | 1·00 | 0·91 | 1·02 | 1·01 | 0·92 | 0·87 | 0·87 | 1·50 | 1·50 |
| $2\times10^{7}$ | 1·00 | 1·19 | 1·28 | 0·96 | 0·97 | 0·92 | 1·12 | 0·81 | 1·28 |
| $2\times10^{7}$ | 1·00 | 1·11 | 8·15 | 5·11 | 3·05 | 1·59 | 1·75 | 1·65 | 8·15 |
| $2\times10^{7}$ | 1·00 | 1·17 | 1·62 | 1·26 | 1·13 | 1·14 | 1·38 | 1·05 | 1·62 |

| Primary CT-B IgA (RAU) | | | | | | | | | | |
|---|---|---|---|---|---|---|---|---|---|---|
| Dose interval (days) | Dose (CFU) | D1 | D4 | D7 | D15 | D29 | D57 | D85 | D180 | max |
| 389 | $10^{8}$ | 0·01314 | 0·01533 | 0·05163 | 0·34261 | 0·05408 | 0·03038 | 0·02163 | 0·01358 | 0·34261 |
| 405 | $10^{8}$ | 0·01947 | 0·01907 | 0·01785 | 0·02697 | 0·02964 | 0·02451 | 0·02574 | 0·02815 | 0·02964 |
| 322 | $10^{7}$ | 0·00909 | 0·00769 | 0·01418 | 0·01846 | 0·01701 | 0·01403 | 0·01116 | 0·00972 | 0·01846 |
| 418 | $10^{7}$ | 0·01568 | 0·01670 | 0·01982 | 0·02319 | 0·02881 | 0·01776 | 0·01876 | 0·02153 | 0·02881 |
| 378 | $10^{8}$ | 0·01867 | 0·02081 | 0·29560 | 1·47954 | 0·42018 | 0·04436 | 0·03394 | 0·02601 | 1·47954 |
| 399 | $10^{8}$ | 0·05973 | 0·06631 | 0·05716 | 0·03010 | 0·11680 | 0·07038 | 0·05513 | 0·03865 | 0·11680 |
| 568 | $10^{7}$ | 0·01141 | 0·01226 | 0·01998 | 0·09696 | 0·04559 | 0·02034 | 0·01781 | 0·01269 | 0·09696 |
| 367 | $10^{8}$ | 0·02455 | 0·02626 | 0·02397 | 0·02603 | 0·02124 | 0·02594 | 0·02133 | 0·02298 | 0·02626 |
| 503 | $10^{7}$ | 0·03069 | 0·02525 | 0·02950 | 0·07328 | 0·05152 | 0·03268 | 0·02505 | 0·00800 | 0·07328 |
| 501 | $10^{8}$ | 0·02841 | 0·02746 | 0·05777 | 0·07595 | 0·04710 | 0·03310 | 0·01541 | 0·02079 | 0·07595 |
| 751 | $10^{10}$ | 0·00146 | 0·00158 | 0·00614 | 0·12045 | 0·03742 | 0·02164 | 0·01506 | 0·01038 | 0·12045 |
| 546 | $10^{8}$ | 0·02052 | 0·01901 | 0·09017 | 0·29997 | 0·20807 | 0·09570 | 0·05812 | 0·02518 | 0·29997 |
| 945 | $10^{8}$ | 0·06446 | 0·07413 | 0·06444 | 1·60204 | 1·05840 | 0·51290 | 0·57179 | 0·44060 | 1·60204 |
| 802 | $10^{10}$ | 0·03379 | 0·03811 | 0·08379 | 0·29181 | | | 0·06284 | 0·05550 | 0·29181 |
| 960 | $10^{8}$ | 0·01603 | 0·01373 | 0·01451 | 0·05641 | 0·05567 | 0·02918 | 0·01614 | 0·01586 | 0·05641 |
| 375 | $10^{5}$ | 0·02759 | 0·02602 | 0·02710 | 0·02134 | 0·02405 | 0·02090 | 0·02694 | 0·03981 | 0·03981 |
| 350 | $10^{5}$ | 0·01653 | 0·01839 | 0·01947 | 0·02181 | 0·03444 | 0·02170 | 0·01917 | 0·02551 | 0·03444 |
| 1111 | $10^{6}$ | 0·08621 | 0·06931 | 0·18598 | 0·59258 | 0·16387 | 0·08949 | 0·08354 | 0·07132 | 0·59258 |
| 576 | $10^{7}$ | 0·02049 | 0·02349 | 0·06504 | 0·83940 | 0·35016 | 0·11330 | 0·07455 | 0·04749 | 0·83940 |
| 520 | $10^{7}$ | 0·02642 | 0·02891 | 0·05162 | 0·17751 | 0·10531 | 0·05679 | 0·07933 | 0·05750 | 0·17751 |

| Primary fold change CT-B IgA | | | | | | | | | |
|---|---|---|---|---|---|---|---|---|---|
| Dose (CFU) | D1 | D4 | D7 | D15 | D29 | D57 | D85 | D180 | max FC |
| $10^{8}$ | 1·00 | 1·17 | 3·93 | 26·07 | 4·12 | 2·31 | 1·65 | 1·03 | 26·07 |
| $10^{8}$ | 1·00 | 0·98 | 0·92 | 1·39 | 1·52 | 1·26 | 1·32 | 1·45 | 1·52 |
| $10^{7}$ | 1·00 | 0·85 | 1·56 | 2·03 | 1·87 | 1·54 | 1·23 | 1·07 | 2·03 |
| $10^{7}$ | 1·00 | 1·07 | 1·26 | 1·48 | 1·84 | 1·13 | 1·20 | 1·37 | 1·84 |
| $10^{8}$ | 1·00 | 1·11 | 15·83 | 79·25 | 22·51 | 2·38 | 1·82 | 1·39 | 79·25 |
| $10^{8}$ | 1·00 | 1·11 | 0·96 | 0·50 | 1·96 | 1·18 | 0·92 | 0·65 | 1·96 |
| $10^{7}$ | 1·00 | 1·07 | 1·75 | 8·50 | 4·00 | 1·78 | 1·56 | 1·11 | 8·50 |
| $10^{8}$ | 1·00 | 1·07 | 0·98 | 1·06 | 0·87 | 1·06 | 0·87 | 0·94 | 1·07 |
| $10^{7}$ | 1·00 | 0·82 | 0·96 | 2·39 | 1·68 | 1·06 | 0·82 | 0·26 | 2·39 |
| $10^{8}$ | 1·00 | 0·97 | 2·03 | 2·67 | 1·66 | 1·17 | 0·54 | 0·73 | 2·67 |
| $10^{10}$ | 1·00 | 1·08 | 4·21 | 82·50 | 25·63 | 14·82 | 10·32 | 7·11 | 82·50 |
| $10^{8}$ | 1·00 | 0·93 | 4·39 | 14·62 | 10·14 | 4·66 | 2·83 | 1·23 | 14·62 |
| $10^{8}$ | 1·00 | 1·15 | 1·00 | 24·85 | 16·42 | 7·96 | 8·87 | 6·84 | 24·85 |
| $10^{10}$ | 1·00 | 1·13 | 2·48 | 8·64 | | | 1·86 | 1·64 | 8·64 |
| $10^{8}$ | 1·00 | 0·86 | 0·91 | 3·52 | 3·47 | 1·82 | 1·01 | 0·99 | 3·52 |
| $10^{5}$ | 1·00 | 0·94 | 0·98 | 0·77 | 0·87 | 0·76 | 0·98 | 1·44 | 1·44 |
| $10^{5}$ | 1·00 | 1·11 | 1·18 | 1·32 | 2·08 | 1·31 | 1·16 | 1·54 | 2·08 |
| $10^{6}$ | 1·00 | 0·80 | 2·16 | 6·87 | 1·90 | 1·04 | 0·97 | 0·83 | 6·87 |
| $10^{7}$ | 1·00 | 1·15 | 3·17 | 40·97 | 17·09 | 5·53 | 3·64 | 2·32 | 40·97 |
| $10^{7}$ | 1·00 | 1·09 | 1·95 | 6·72 | 3·99 | 2·15 | 3·00 | 2·18 | 6·72 |

| Boost TcpA IgM (RAU) | | | | | | | | | | |
|---|---|---|---|---|---|---|---|---|---|---|
| Dose interval (days) | Dose (CFU) | D1 | D4 | D7 | D15 | D29 | D57 | D85 | D180 | max |
| 389 | $2\times10^7$ | 0·73322 | 0·79170 | 1·19122 | 0·64649 | 0·39480 | 0·44525 | 0·33950 | 0·35372 | 1·19122 |
| 405 | $2\times10^7$ | 0·05644 | 0·09753 | 0·11702 | 0·31891 | 0·18954 | 0·12081 | 0·12515 | 0·10955 | 0·31891 |
| 322 | $2\times10^7$ | 0·26453 | 0·24222 | 0·25393 | 0·25925 | 0·26663 | 1·28998 | 0·72363 | 0·52099 | 1·28998 |
| 418 | $2\times10^7$ | 0·16317 | 0·17302 | 0·15849 | 0·16377 | 0·18729 | 0·20356 | 0·18515 | 0·15354 | 0·20356 |
| 378 | $2\times10^7$ | 0·12209 | 0·13571 | 0·19181 | 0·29994 | 0·18671 | 0·17013 | 0·12689 | 0·10658 | 0·29994 |
| 399 | $2\times10^7$ | 0·31071 | 0·29320 | 0·27185 | 0·25180 | 0·27867 | 0·18738 | 0·18377 | 0·19016 | 0·31071 |
| 568 | $2\times10^7$ | 0·22385 | 0·17476 | 0·63045 | 2·31000 | 1·08155 | 0·43727 | 0·24176 | 0·21822 | 2·31000 |
| 367 | $2\times10^7$ | 0·34431 | 0·37894 | 0·68593 | 0·53906 | 0·41863 | 0·25210 | 0·25456 | 0·24256 | 0·68593 |
| 503 | $2\times10^7$ | 0·28700 | 0·27916 | 0·26241 | 0·25712 | 0·31017 | 0·26112 | 0·26898 | 0·15495 | 0·31017 |
| 501 | $2\times10^7$ | 0·32194 | 0·33672 | 0·37791 | 0·32145 | 0·28023 | 0·21715 | 0·34988 | 0·23965 | 0·37791 |
| 751 | $2\times10^7$ | 0·25184 | 0·29998 | 0·38967 | 0·40285 | 0·31006 | 0·25049 | 0·30219 | 0·11899 | 0·40285 |
| 546 | $2\times10^7$ | 0·36054 | 0·43611 | 0·34527 | 0·34388 | 0·33271 | 0·32427 | 0·30245 | 0·16914 | 0·43611 |
| 945 | $2\times10^7$ | 0·09906 | 0·10026 | 0·14216 | 0·17042 | 0·11961 | 0·11726 | 0·12048 | 0·07246 | 0·17042 |
| 802 | $2\times10^7$ | 0·26654 | 0·27850 | 1·11590 | 0·65933 | 0·47414 | | | 0·23064 | 1·11590 |
| 960 | $2\times10^7$ | 0·24286 | 0·22372 | 0·61655 | 0·43833 | 0·33979 | 0·27304 | 0·25429 | 0·13749 | 0·61655 |
| 375 | $2\times10^7$ | 0·23466 | 0·20586 | 0·51641 | 0·34802 | 0·27659 | 0·35946 | 0·27740 | 0·12921 | 0·51641 |
| 350 | $2\times10^7$ | 0·30571 | 0·30488 | 0·33813 | 0·29185 | 0·29672 | 0·29185 | 0·29020 | 0·23742 | 0·33813 |
| 1111 | $2\times10^7$ | 0·49176 | 0·59008 | 0·45111 | 0·47605 | 0·40700 | 0·35892 | 0·33664 | 0·31293 | 0·59008 |
| 576 | $2\times10^7$ | 0·12341 | 0·13586 | 0·21715 | 0·15070 | 0·18127 | 0·13671 | 0·11291 | 0·10158 | 0·21715 |
| 520 | $2\times10^7$ | 0·23631 | 0·28586 | 0·37812 | 0·30707 | 0·32045 | 0·32835 | 0·18060 | 0·15141 | 0·37812 |

| Boost fold change TcpA IgM | | | | | | | | | |
|---|---|---|---|---|---|---|---|---|---|
| Dose (CFU) | D1 | D4 | D7 | D15 | D29 | D57 | D85 | D180 | max FC |
| $2\times10^7$ | 1·00 | 1·08 | 1·62 | 0·88 | 0·54 | 0·61 | 0·46 | 0·48 | 1·62 |
| $2\times10^7$ | 1·00 | 1·73 | 2·07 | 5·65 | 3·36 | 2·14 | 2·22 | 1·94 | 5·65 |
| $2\times10^7$ | 1·00 | 0·92 | 0·96 | 0·98 | 1·01 | 4·88 | 2·74 | 1·97 | 4·88 |
| $2\times10^7$ | 1·00 | 1·06 | 0·97 | 1·00 | 1·15 | 1·25 | 1·13 | 0·94 | 1·25 |
| $2\times10^7$ | 1·00 | 1·11 | 1·57 | 2·46 | 1·53 | 1·39 | 1·04 | 0·87 | 2·46 |
| $2\times10^7$ | 1·00 | 0·94 | 0·87 | 0·81 | 0·90 | 0·60 | 0·59 | 0·61 | 0·94 |
| $2\times10^7$ | 1·00 | 0·78 | 2·82 | 10·32 | 4·83 | 1·95 | 1·08 | 0·97 | 10·32 |
| $2\times10^7$ | 1·00 | 1·10 | 1·99 | 1·57 | 1·22 | 0·73 | 0·74 | 0·70 | 1·99 |
| $2\times10^7$ | 1·00 | 0·97 | 0·91 | 0·90 | 1·08 | 0·91 | 0·94 | 0·54 | 1·08 |
| $2\times10^7$ | 1·00 | 1·05 | 1·17 | 1·00 | 0·87 | 0·67 | 1·09 | 0·74 | 1·17 |
| $2\times10^7$ | 1·00 | 1·19 | 1·55 | 1·60 | 1·23 | 0·99 | 1·20 | 0·47 | 1·60 |
| $2\times10^7$ | 1·00 | 1·21 | 0·96 | 0·95 | 0·92 | 0·90 | 0·84 | 0·47 | 1·21 |
| $2\times10^7$ | 1·00 | 1·01 | 1·44 | 1·72 | 1·21 | 1·18 | 1·22 | 0·73 | 1·72 |
| $2\times10^7$ | 1·00 | 1·04 | 4·19 | 2·47 | 1·78 | | | 0·87 | 4·19 |
| $2\times10^7$ | 1·00 | 0·92 | 2·54 | 1·80 | 1·40 | 1·12 | 1·05 | 0·57 | 2·54 |
| $2\times10^7$ | 1·00 | 0·88 | 2·20 | 1·48 | 1·18 | 1·53 | 1·18 | 0·55 | 2·20 |
| $2\times10^7$ | 1·00 | 1·00 | 1·11 | 0·95 | 0·97 | 0·95 | 0·95 | 0·78 | 1·11 |
| $2\times10^7$ | 1·00 | 1·20 | 0·92 | 0·97 | 0·83 | 0·73 | 0·68 | 0·64 | 1·20 |
| $2\times10^7$ | 1·00 | 1·10 | 1·76 | 1·22 | 1·47 | 1·11 | 0·91 | 0·82 | 1·76 |
| $2\times10^7$ | 1·00 | 1·21 | 1·60 | 1·30 | 1·36 | 1·39 | 0·76 | 0·64 | 1·60 |

| Primary TcpA IgM (RAU) | | | | | | | | | | |
|---|---|---|---|---|---|---|---|---|---|---|
| Dose interval (days) | Dose (CFU) | D1 | D4 | D7 | D15 | D29 | D57 | D85 | D180 | max |
| 389 | $10^8$ | 0·92444 | 1·12108 | 1·14131 | 1·34008 | 0·74836 | 0·56072 | 0·78195 | 0·64960 | 1·34008 |
| 405 | $10^8$ | 0·17272 | 0·14152 | 0·14350 | 0·19144 | 0·22752 | 0·23925 | 0·25204 | 0·24717 | 0·25204 |
| 322 | $10^7$ | 0·33894 | 0·36862 | 0·68709 | 0·93325 | 0·59666 | 0·39657 | 0·37405 | 0·34252 | 0·93325 |
| 418 | $10^7$ | 0·44836 | 0·49713 | 0·44036 | 0·42005 | 0·33160 | 0·35053 | 0·21967 | 0·41001 | 0·49713 |
| 378 | $10^8$ | 0·25754 | 0·27236 | 0·23769 | 0·22015 | 0·18087 | 0·18754 | 0·18501 | 0·20329 | 0·27236 |
| 399 | $10^8$ | 0·54132 | 0·49910 | 0·51376 | 0·80185 | 0·97375 | 0·76576 | 0·75622 | 0·83623 | 0·97375 |
| 568 | $10^7$ | 0·24685 | 0·23664 | 0·30269 | 2·40990 | 1·17831 | 0·43129 | 0·49516 | 0·32105 | 2·40990 |
| 367 | $10^8$ | 0·70153 | 0·93721 | 1·00539 | 1·00539 | 0·94120 | 0·77868 | 0·87036 | 0·94721 | 1·00539 |
| 503 | $10^7$ | 0·54446 | 0·45112 | 0·75461 | 2·46897 | 1·09158 | 0·47250 | 0·42854 | 0·45963 | 2·46897 |
| 501 | $10^8$ | 0·55276 | 0·54818 | 0·63820 | 0·77113 | 0·48171 | 0·81840 | 0·58201 | 0·51831 | 0·81840 |
| 751 | $10^{10}$ | 0·20740 | 0·23989 | 0·29772 | 1·47887 | 0·68902 | 0·34249 | 0·71388 | 0·33300 | 1·47887 |
| 546 | $10^8$ | 1·58581 | 1·72279 | 1·58841 | 0·86662 | 1·14503 | 1·16470 | 0·92930 | 0·66738 | 1·72279 |
| 945 | $10^8$ | 0·20165 | 0·19573 | 0·13347 | 0·14026 | 0·16218 | 0·17654 | 0·17934 | 0·08548 | 0·20165 |
| 802 | $10^{10}$ | 0·37240 | 0·48458 | 0·26105 | 0·32828 | | | 0·40211 | 0·32429 | 0·48458 |
| 960 | $10^8$ | 0·83273 | 0·76748 | 0·77831 | 1·10424 | 0·81857 | 0·78788 | 0·34066 | 0·39152 | 1·10424 |
| 375 | $10^5$ | 0·32086 | 0·33258 | 0·39082 | 0·50454 | 0·36554 | 0·23154 | 0·33398 | 0·15468 | 0·50454 |
| 350 | $10^5$ | 0·39082 | 0·36478 | 0·46058 | 0·79350 | 0·58322 | 0·42920 | 0·40156 | 0·23056 | 0·79350 |
| 1111 | $10^6$ | 1·01334 | 1·18599 | 1·42736 | 1·80176 | 1·01001 | 1·13339 | 1·01667 | 0·74940 | 1·80176 |
| 576 | $10^7$ | 0·22952 | 0·24660 | 0·22749 | 0·30269 | 0·28109 | 0·39082 | 0·31951 | 0·30110 | 0·39082 |
| 520 | $10^7$ | 0·48608 | 0·47219 | 0·67575 | 0·67857 | 0·64557 | 0·66877 | 0·64156 | 0·53687 | 0·67857 |

| Primary fold change TcpA IgM | | | | | | | | | |
|---|---|---|---|---|---|---|---|---|---|
| Dose (CFU) | D1 | D4 | D7 | D15 | D29 | D57 | D85 | D180 | max FC |
| $10^8$ | 1·00 | 1·21 | 1·23 | 1·45 | 0·81 | 0·61 | 0·85 | 0·70 | 1·45 |
| $10^8$ | 1·00 | 0·82 | 0·83 | 1·11 | 1·32 | 1·39 | 1·46 | 1·43 | 1·46 |
| $10^7$ | 1·00 | 1·09 | 2·03 | 2·75 | 1·76 | 1·17 | 1·10 | 1·01 | 2·75 |
| $10^7$ | 1·00 | 1·11 | 0·98 | 0·94 | 0·74 | 0·78 | 0·49 | 0·91 | 1·11 |
| $10^8$ | 1·00 | 1·06 | 0·92 | 0·85 | 0·70 | 0·73 | 0·72 | 0·79 | 1·06 |
| $10^8$ | 1·00 | 0·92 | 0·95 | 1·48 | 1·80 | 1·41 | 1·40 | 1·54 | 1·80 |
| $10^7$ | 1·00 | 0·96 | 1·23 | 9·76 | 4·77 | 1·75 | 2·01 | 1·30 | 9·76 |
| $10^8$ | 1·00 | 1·34 | 1·43 | 1·43 | 1·34 | 1·11 | 1·24 | 1·35 | 1·43 |
| $10^7$ | 1·00 | 0·83 | 1·39 | 4·53 | 2·00 | 0·87 | 0·79 | 0·84 | 4·53 |
| $10^8$ | 1·00 | 0·99 | 1·15 | 1·40 | 0·87 | 1·48 | 1·05 | 0·94 | 1·48 |
| $10^{10}$ | 1·00 | 1·16 | 1·44 | 7·13 | 3·32 | 1·65 | 3·44 | 1·61 | 7·13 |
| $10^8$ | 1·00 | 1·09 | 1·00 | 0·55 | 0·72 | 0·73 | 0·59 | 0·42 | 1·09 |
| $10^8$ | 1·00 | 0·97 | 0·66 | 0·70 | 0·80 | 0·88 | 0·89 | 0·42 | 0·97 |
| $10^{10}$ | 1·00 | 1·30 | 0·70 | 0·88 | | | 1·08 | 0·87 | 1·30 |
| $10^8$ | 1·00 | 0·92 | 0·93 | 1·33 | 0·98 | 0·95 | 0·41 | 0·47 | 1·33 |
| $10^5$ | 1·00 | 1·04 | 1·22 | 1·57 | 1·14 | 0·72 | 1·04 | 0·48 | 1·57 |
| $10^5$ | 1·00 | 0·93 | 1·18 | 2·03 | 1·49 | 1·10 | 1·03 | 0·59 | 2·03 |
| $10^6$ | 1·00 | 1·17 | 1·41 | 1·78 | 1·00 | 1·12 | 1·00 | 0·74 | 1·78 |
| $10^7$ | 1·00 | 1·07 | 0·99 | 1·32 | 1·22 | 1·70 | 1·39 | 1·31 | 1·70 |
| $10^7$ | 1·00 | 0·97 | 1·39 | 1·40 | 1·33 | 1·38 | 1·32 | 1·10 | 1·40 |

| Boost TcpA IgG (RAU) | | | | | | | | | | |
|---|---|---|---|---|---|---|---|---|---|---|
| Dose interval (days) | Dose (CFU) | D1 | D4 | D7 | D15 | D29 | D57 | D85 | D180 | max |
| 389 | $2\times10^7$ | 0·02808 | 0·02026 | 0·92943 | 1·93251 | 0·50953 | 0·96364 | 0·12191 | 0·04168 | 1·93251 |
| 405 | $2\times10^7$ | 0·00213 | 0·00398 | 0·01924 | 0·30197 | 0·13110 | 0·04383 | 0·01371 | 0·00474 | 0·30197 |
| 322 | $2\times10^7$ | 0·01101 | 0·00875 | 0·01014 | 0·01022 | 0·01158 | 0·01256 | 0·00413 | 0·00364 | 0·01256 |
| 418 | $2\times10^7$ | 0·00731 | 0·00956 | 0·01200 | 0·02424 | 0·02215 | 0·01470 | 0·01208 | 0·00686 | 0·02424 |
| 378 | $2\times10^7$ | 0·02014 | 0·02020 | 0·20931 | 0·96215 | 0·60456 | 0·37644 | 0·14994 | 0·06504 | 0·96215 |
| 399 | $2\times10^7$ | 0·00685 | 0·00591 | 0·00654 | 0·00630 | 0·00544 | 0·00583 | 0·00571 | 0·00587 | 0·00685 |
| 568 | $2\times10^7$ | 0·01108 | 0·00971 | 0·10318 | 2·32999 | 1·24183 | 0·86346 | 0·33560 | 0·02298 | 2·32999 |
| 367 | $2\times10^7$ | 0·00544 | 0·00669 | 0·04266 | 0·03295 | 0·02001 | 0·00921 | 0·00630 | 0·00783 | 0·04266 |
| 503 | $2\times10^7$ | 0·04596 | 0·05111 | 0·04659 | 0·04925 | 0·04006 | 0·03766 | 0·03555 | 0·03533 | 0·05111 |
| 501 | $2\times10^7$ | 0·01577 | 0·01604 | 0·17628 | 0·24865 | 0·20684 | 0·06269 | 0·04066 | 0·02568 | 0·24865 |
| 751 | $2\times10^7$ | 0·09382 | 0·08991 | 0·25507 | 1·16708 | 0·71792 | 0·36335 | 0·35408 | 0·04157 | 1·16708 |
| 546 | $2\times10^7$ | 0·02628 | 0·01785 | 0·08081 | 0·09318 | 0·10521 | 0·06937 | 0·07455 | 0·05905 | 0·10521 |
| 945 | $2\times10^7$ | 0·00552 | 0·00509 | 0·03160 | 0·66403 | 0·16206 | 0·07920 | 0·05198 | 0·01737 | 0·66403 |
| 802 | $2\times10^7$ | 0·00716 | 0·00730 | 0·44704 | 0·28289 | 0·21675 | | | 0·02317 | 0·44704 |
| 960 | $2\times10^7$ | 0·00819 | 0·00641 | 0·04814 | 0·08623 | 0·04843 | 0·02183 | 0·01400 | 0·01371 | 0·08623 |
| 375 | $2\times10^7$ | 0·00916 | 0·00980 | 0·84875 | 0·65554 | 0·43832 | 0·22357 | 0·12030 | 0·04992 | 0·84875 |
| 350 | $2\times10^7$ | 0·15110 | 0·11866 | 0·26789 | 0·28064 | 0·20694 | 0·14951 | 0·12226 | 0·20956 | 0·28064 |
| 1111 | $2\times10^7$ | 0·00393 | 0·00448 | 0·00399 | 0·00478 | 0·00466 | 0·00364 | 0·00695 | 0·00759 | 0·00759 |
| 576 | $2\times10^7$ | 0·00421 | 0·00444 | 0·05503 | 0·09669 | 0·08462 | 0·04863 | 0·03043 | 0·01050 | 0·09669 |
| 520 | $2\times10^7$ | 0·01214 | 0·01285 | 0·12646 | 0·10156 | 0·06504 | 0·04046 | 0·03090 | 0·01868 | 0·12646 |

| Boost fold change TcpA IgG | | | | | | | | | |
|---|---|---|---|---|---|---|---|---|---|
| Dose (CFU) | D1 | D4 | D7 | D15 | D29 | D57 | D85 | D180 | max FC |
| $2\times10^7$ | 1·00 | 0·72 | 33·10 | 68·83 | 18·15 | 34·32 | 4·34 | 1·48 | 68·83 |
| $2\times10^7$ | 1·00 | 1·86 | 9·01 | 141·47 | 61·42 | 20·53 | 6·42 | 2·22 | 141·47 |
| $2\times10^7$ | 1·00 | 0·79 | 0·92 | 0·93 | 1·05 | 1·14 | 0·38 | 0·33 | 1·14 |
| $2\times10^7$ | 1·00 | 1·31 | 1·64 | 3·32 | 3·03 | 2·01 | 1·65 | 0·94 | 3·32 |
| $2\times10^7$ | 1·00 | 1·00 | 10·40 | 47·78 | 30·02 | 18·70 | 7·45 | 3·23 | 47·78 |
| $2\times10^7$ | 1·00 | 0·86 | 0·95 | 0·92 | 0·79 | 0·85 | 0·83 | 0·86 | 0·95 |
| $2\times10^7$ | 1·00 | 0·88 | 9·31 | 210·29 | 112·08 | 77·93 | 30·29 | 2·07 | 210·29 |
| $2\times10^7$ | 1·00 | 1·23 | 7·85 | 6·06 | 3·68 | 1·69 | 1·16 | 1·44 | 7·85 |
| $2\times10^7$ | 1·00 | 1·11 | 1·01 | 1·07 | 0·87 | 0·82 | 0·77 | 0·77 | 1·11 |
| $2\times10^7$ | 1·00 | 1·02 | 11·18 | 15·76 | 13·11 | 3·97 | 2·58 | 1·63 | 15·76 |
| $2\times10^7$ | 1·00 | 0·96 | 2·72 | 12·44 | 7·65 | 3·87 | 3·77 | 0·44 | 12·44 |
| $2\times10^7$ | 1·00 | 0·68 | 3·08 | 3·55 | 4·00 | 2·64 | 2·84 | 2·25 | 4·00 |
| $2\times10^7$ | 1·00 | 0·92 | 5·72 | 120·29 | 29·36 | 14·35 | 9·41 | 3·15 | 120·29 |
| $2\times10^7$ | 1·00 | 1·02 | 62·41 | 39·49 | 30·26 | | | 3·24 | 62·41 |
| $2\times10^7$ | 1·00 | 0·78 | 5·87 | 10·52 | 5·91 | 2·66 | 1·71 | 1·67 | 10·52 |
| $2\times10^7$ | 1·00 | 1·07 | 92·68 | 71·58 | 47·86 | 24·41 | 13·14 | 5·45 | 92·68 |
| $2\times10^7$ | 1·00 | 0·79 | 1·77 | 1·86 | 1·37 | 0·99 | 0·81 | 1·39 | 1·86 |
| $2\times10^7$ | 1·00 | 1·14 | 1·01 | 1·22 | 1·18 | 0·92 | 1·77 | 1·93 | 1·93 |
| $2\times10^7$ | 1·00 | 1·06 | 13·08 | 22·97 | 20·11 | 11·55 | 7·23 | 2·49 | 22·97 |
| $2\times10^7$ | 1·00 | 1·06 | 10·42 | 8·37 | 5·36 | 3·33 | 2·55 | 1·54 | 10·42 |

| Primary TcpA IgG (RAU) | | | | | | | | | | |
|---|---|---|---|---|---|---|---|---|---|---|
| Dose interval (days) | Dose (CFU) | D1 | D4 | D7 | D15 | D29 | D57 | D85 | D180 | max |
| 389 | $10^8$ | 0·01495 | 0·01545 | 0·01673 | 0·02134 | 0·01476 | 0·01836 | 0·02456 | 0·01463 | 0·02456 |
| 405 | $10^8$ | 0·00426 | 0·00297 | 0·00468 | 0·00568 | 0·00066 | 0·00043 | 0·00078 | 0·00066 | 0·00568 |
| 322 | $10^7$ | 0·00270 | 0·00388 | 0·00442 | 0·00344 | 0·00457 | 0·00353 | 0·00491 | 0·00447 | 0·00491 |
| 418 | $10^7$ | 0·01502 | 0·01268 | 0·01296 | 0·01407 | 0·01282 | 0·00235 | 0·00442 | 0·00329 | 0·01502 |
| 378 | $10^8$ | 0·00718 | 0·01004 | 0·00812 | 0·00904 | 0·01337 | 0·02250 | 0·02015 | 0·01632 | 0·02250 |
| 399 | $10^8$ | 0·01012 | 0·01072 | 0·00935 | 0·00462 | 0·00353 | 0·00398 | 0·00521 | 0·00319 | 0·01072 |
| 568 | $10^7$ | 0·01578 | 0·01637 | 0·01522 | 0·01893 | 0·01709 | 0·01699 | 0·01858 | 0·01676 | 0·01893 |
| 367 | $10^8$ | 0·00314 | 0·00270 | 0·00211 | 0·00280 | 0·00226 | 0·00240 | 0·00211 | 0·00245 | 0·00314 |
| 503 | $10^7$ | 0·01608 | 0·01641 | 0·01427 | 0·02482 | 0·02657 | 0·05390 | 0·05512 | 0·04582 | 0·05512 |
| 501 | $10^8$ | 0·00997 | 0·01275 | 0·01050 | 0·01390 | 0·01502 | 0·02287 | 0·02182 | 0·01780 | 0·02287 |
| 751 | $10^{10}$ | 0·01506 | 0·01613 | 0·01519 | 0·01280 | 0·02421 | 0·03881 | 0·03073 | 0·02502 | 0·03881 |
| 546 | $10^8$ | 0·01562 | 0·01588 | 0·01558 | 0·01410 | 0·02042 | 0·07168 | 0·07542 | 0·02353 | 0·07542 |
| 945 | $10^8$ | 0·02136 | 0·02364 | 0·02546 | 0·02265 | 0·02698 | 0·01417 | 0·01754 | 0·00652 | 0·02698 |
| 802 | $10^{10}$ | 0·00264 | 0·00312 | 0·00312 | 0·00386 | | | 0·00373 | 0·00361 | 0·00386 |
| 960 | $10^8$ | 0·01146 | 0·01064 | 0·00760 | 0·00742 | 0·00742 | 0·00846 | 0·00652 | 0·00716 | 0·01146 |
| 375 | $10^5$ | 0·00358 | 0·00349 | 0·00737 | 0·00675 | 0·00506 | 0·01151 | 0·01684 | 0·01151 | 0·01684 |
| 350 | $10^5$ | 0·01062 | 0·01195 | 0·00995 | 0·01595 | 0·02076 | 0·05916 | 0·09098 | 0·10921 | 0·10921 |
| 1111 | $10^6$ | 0·00565 | 0·00583 | 0·00565 | 0·00931 | 0·00777 | 0·00964 | 0·00511 | 0·00981 | 0·00981 |
| 576 | $10^7$ | 0·00993 | 0·01027 | 0·00997 | 0·01345 | 0·01400 | 0·00718 | 0·00378 | 0·00427 | 0·01400 |
| 520 | $10^7$ | 0·00477 | 0·00467 | 0·00349 | 0·00555 | 0·00755 | 0·00774 | 0·00858 | 0·00486 | 0·00858 |

| Primary fold change TcpA IgG | | | | | | | | | |
|---|---|---|---|---|---|---|---|---|---|
| Dose (CFU) | D1 | D4 | D7 | D15 | D29 | D57 | D85 | D180 | max FC |
| $10^8$ | 1·00 | 1·03 | 1·12 | 1·43 | 0·99 | 1·23 | 1·64 | 0·98 | 1·64 |
| $10^8$ | 1·00 | 0·70 | 1·10 | 1·33 | 0·15 | 0·10 | 0·18 | 0·15 | 1·33 |
| $10^7$ | 1·00 | 1·44 | 1·64 | 1·27 | 1·69 | 1·31 | 1·82 | 1·66 | 1·82 |
| $10^7$ | 1·00 | 0·84 | 0·86 | 0·94 | 0·85 | 0·16 | 0·29 | 0·22 | 0·94 |
| $10^8$ | 1·00 | 1·40 | 1·13 | 1·26 | 1·86 | 3·13 | 2·81 | 2·27 | 3·13 |
| $10^8$ | 1·00 | 1·06 | 0·92 | 0·46 | 0·35 | 0·39 | 0·51 | 0·32 | 1·06 |
| $10^7$ | 1·00 | 1·04 | 0·96 | 1·20 | 1·08 | 1·08 | 1·18 | 1·06 | 1·20 |
| $10^8$ | 1·00 | 0·86 | 0·67 | 0·89 | 0·72 | 0·76 | 0·67 | 0·78 | 0·89 |
| $10^7$ | 1·00 | 1·02 | 0·89 | 1·54 | 1·65 | 3·35 | 3·43 | 2·85 | 3·43 |
| $10^8$ | 1·00 | 1·28 | 1·05 | 1·39 | 1·51 | 2·29 | 2·19 | 1·79 | 2·29 |
| $10^{10}$ | 1·00 | 1·07 | 1·01 | 0·85 | 1·61 | 2·58 | 2·04 | 1·66 | 2·58 |
| $10^8$ | 1·00 | 1·02 | 1·00 | 0·90 | 1·31 | 4·59 | 4·83 | 1·51 | 4·83 |
| $10^8$ | 1·00 | 1·11 | 1·19 | 1·06 | 1·26 | 0·66 | 0·82 | 0·31 | 1·26 |
| $10^{10}$ | 1·00 | 1·18 | 1·18 | 1·46 | | | 1·41 | 1·37 | 1·46 |
| $10^8$ | 1·00 | 0·93 | 0·66 | 0·65 | 0·65 | 0·74 | 0·57 | 0·62 | 0·93 |
| $10^5$ | 1·00 | 0·97 | 2·06 | 1·89 | 1·41 | 3·22 | 4·70 | 3·21 | 4·70 |
| $10^5$ | 1·00 | 1·13 | 0·94 | 1·50 | 1·95 | 5·57 | 8·57 | 10·28 | 10·28 |
| $10^6$ | 1·00 | 1·03 | 1·00 | 1·65 | 1·38 | 1·71 | 0·90 | 1·74 | 1·74 |
| $10^7$ | 1·00 | 1·03 | 1·00 | 1·35 | 1·41 | 0·72 | 0·38 | 0·43 | 1·41 |
| $10^7$ | 1·00 | 0·98 | 0·73 | 1·16 | 1·58 | 1·62 | 1·80 | 1·02 | 1·80 |

| Boost TcpA IgA (RAU) | | | | | | | | | | |
|---|---|---|---|---|---|---|---|---|---|---|
| Dose interval (days) | Dose (CFU) | D1 | D4 | D7 | D15 | D29 | D57 | D85 | D180 | max |
| 389 | $2\times10^7$ | 0·01435 | 0·01371 | 0·76732 | 0·40571 | 0·03483 | 0·05680 | 0·01562 | 0·01079 | 0·76732 |
| 405 | $2\times10^7$ | 0·01103 | 0·00976 | 0·08503 | 0·48930 | 0·06533 | 0·02174 | 0·01355 | 0·01190 | 0·48930 |
| 322 | $2\times10^7$ | 0·00717 | 0·00682 | 0·00711 | 0·00658 | 0·00787 | 0·01037 | 0·00686 | 0·00649 | 0·01037 |
| 418 | $2\times10^7$ | 0·01155 | 0·01171 | 0·01853 | 0·02554 | 0·01654 | 0·01308 | 0·01568 | 0·01219 | 0·02554 |
| 378 | $2\times10^7$ | 0·02835 | 0·02746 | 0·44197 | 1·06264 | 0·21545 | 0·06452 | 0·03781 | 0·04312 | 1·06264 |
| 399 | $2\times10^7$ | 0·01480 | 0·01617 | 0·01474 | 0·01517 | 0·01578 | 0·01541 | 0·01497 | 0·01472 | 0·01617 |
| 568 | $2\times10^7$ | 0·01077 | 0·00810 | 0·16651 | 1·20324 | 0·18499 | 0·03327 | 0·01526 | 0·01033 | 1·20324 |
| 367 | $2\times10^7$ | 0·00897 | 0·00930 | 0·14057 | 0·06001 | 0·01898 | 0·01340 | 0·01133 | 0·01036 | 0·14057 |
| 503 | $2\times10^7$ | 0·02230 | 0·02401 | 0·02233 | 0·02938 | 0·02235 | 0·02102 | 0·02270 | 0·05548 | 0·05548 |
| 501 | $2\times10^7$ | 0·01007 | 0·01019 | 0·19773 | 0·09141 | 0·03500 | 0·01494 | 0·01317 | 0·03404 | 0·19773 |
| 751 | $2\times10^7$ | 0·01016 | 0·01035 | 0·10398 | 0·20037 | 0·04399 | 0·01912 | 0·01894 | 0·01690 | 0·20037 |
| 546 | $2\times10^7$ | 0·03059 | 0·02886 | 0·17577 | 0·19461 | 0·10636 | 0·06269 | 0·05363 | 0·07864 | 0·19461 |
| 945 | $2\times10^7$ | 0·01545 | 0·01698 | 0·05580 | 0·16224 | 0·05293 | 0·03150 | 0·02946 | 0·07066 | 0·16224 |
| 802 | $2\times10^7$ | 0·01908 | 0·01861 | 0·49879 | 0·15010 | 0·06338 | | | 0·06833 | 0·49879 |
| 960 | $2\times10^7$ | 0·01821 | 0·01745 | 0·15886 | 0·11018 | 0·03286 | 0·01920 | 0·01806 | 0·03288 | 0·15886 |
| 375 | $2\times10^7$ | 0·02096 | 0·01802 | 1·49279 | 0·68082 | 0·17903 | 0·05702 | 0·03956 | 0·09386 | 1·49279 |
| 350 | $2\times10^7$ | 0·09901 | 0·08970 | 0·48547 | 0·27455 | 0·12651 | 0·11250 | 0·08530 | 0·19162 | 0·48547 |
| 1111 | $2\times10^7$ | 0·02827 | 0·03340 | 0·03652 | 0·02873 | 0·02843 | 0·02546 | 0·06265 | 0·06162 | 0·06265 |
| 576 | $2\times10^7$ | 0·00969 | 0·00990 | 0·11704 | 0·06569 | 0·02833 | 0·01364 | 0·02467 | 0·02187 | 0·11704 |
| 520 | $2\times10^7$ | 0·01591 | 0·01732 | 0·09889 | 0·05616 | 0·02199 | 0·01804 | 0·04235 | 0·03361 | 0·09889 |

| Boost fold change TcpA IgA | | | | | | | | | |
|---|---|---|---|---|---|---|---|---|---|
| Dose (CFU) | D1 | D4 | D7 | D15 | D29 | D57 | D85 | D180 | max FC |
| $2\times10^7$ | 1·00 | 0·96 | 53·49 | 28·28 | 2·43 | 3·96 | 1·09 | 0·75 | 53·49 |
| $2\times10^7$ | 1·00 | 0·89 | 7·71 | 44·37 | 5·92 | 1·97 | 1·23 | 1·08 | 44·37 |
| $2\times10^7$ | 1·00 | 0·95 | 0·99 | 0·92 | 1·10 | 1·44 | 0·96 | 0·90 | 1·44 |
| $2\times10^7$ | 1·00 | 1·01 | 1·60 | 2·21 | 1·43 | 1·13 | 1·36 | 1·06 | 2·21 |
| $2\times10^7$ | 1·00 | 0·97 | 15·59 | 37·48 | 7·60 | 2·28 | 1·33 | 1·52 | 37·48 |
| $2\times10^7$ | 1·00 | 1·09 | 1·00 | 1·03 | 1·07 | 1·04 | 1·01 | 0·99 | 1·09 |
| $2\times10^7$ | 1·00 | 0·75 | 15·46 | 111·73 | 17·18 | 3·09 | 1·42 | 0·96 | 111·73 |
| $2\times10^7$ | 1·00 | 1·04 | 15·67 | 6·69 | 2·11 | 1·49 | 1·26 | 1·16 | 15·67 |
| $2\times10^7$ | 1·00 | 1·08 | 1·00 | 1·32 | 1·00 | 0·94 | 1·02 | 2·49 | 2·49 |
| $2\times10^7$ | 1·00 | 1·01 | 19·64 | 9·08 | 3·48 | 1·48 | 1·31 | 3·38 | 19·64 |
| $2\times10^7$ | 1·00 | 1·02 | 10·23 | 19·72 | 4·33 | 1·88 | 1·86 | 1·66 | 19·72 |
| $2\times10^7$ | 1·00 | 0·94 | 5·75 | 6·36 | 3·48 | 2·05 | 1·75 | 2·57 | 6·36 |
| $2\times10^7$ | 1·00 | 1·10 | 3·61 | 10·50 | 3·43 | 2·04 | 1·91 | 4·57 | 10·50 |
| $2\times10^7$ | 1·00 | 0·98 | 26·15 | 7·87 | 3·32 | | | 3·58 | 26·15 |
| $2\times10^7$ | 1·00 | 0·96 | 8·72 | 6·05 | 1·80 | 1·05 | 0·99 | 1·81 | 8·72 |
| $2\times10^7$ | 1·00 | 0·86 | 71·20 | 32·47 | 8·54 | 2·72 | 1·89 | 4·48 | 71·20 |
| $2\times10^7$ | 1·00 | 0·91 | 4·90 | 2·77 | 1·28 | 1·14 | 0·86 | 1·94 | 4·90 |
| $2\times10^7$ | 1·00 | 1·18 | 1·29 | 1·02 | 1·01 | 0·90 | 2·22 | 2·18 | 2·22 |
| $2\times10^7$ | 1·00 | 1·02 | 12·08 | 6·78 | 2·93 | 1·41 | 2·55 | 2·26 | 12·08 |
| $2\times10^7$ | 1·00 | 1·09 | 6·21 | 3·53 | 1·38 | 1·13 | 2·66 | 2·11 | 6·21 |

| Primary TcpA IgA (RAU) | | | | | | | | | | |
|---|---|---|---|---|---|---|---|---|---|---|
| Dose interval (days) | Dose (CFU) | D1 | D4 | D7 | D15 | D29 | D57 | D85 | D180 | max |
| 389 | $10^8$ | 0·00546 | 0·00702 | 0·00726 | 0·01681 | 0·00880 | 0·00830 | 0·00875 | 0·00650 | 0·01681 |
| 405 | $10^8$ | 0·01088 | 0·00816 | 0·00893 | 0·00875 | 0·00604 | 0·00774 | 0·00774 | 0·00832 | 0·01088 |
| 322 | $10^7$ | 0·00200 | 0·00213 | 0·00288 | 0·00458 | 0·00392 | 0·00299 | 0·00332 | 0·00174 | 0·00458 |
| 418 | $10^7$ | 0·00896 | 0·00746 | 0·00898 | 0·01108 | 0·01002 | 0·01013 | 0·00997 | 0·01203 | 0·01203 |
| 378 | $10^8$ | 0·02204 | 0·02212 | 0·02345 | 0·02227 | 0·02840 | 0·02271 | 0·02103 | 0·01800 | 0·02840 |
| 399 | $10^8$ | 0·01068 | 0·00898 | 0·01007 | 0·00863 | 0·01234 | 0·01134 | 0·01120 | 0·00985 | 0·01234 |
| 568 | $10^7$ | 0·00431 | 0·00562 | 0·00626 | 0·00757 | 0·00655 | 0·00728 | 0·01123 | 0·00490 | 0·01123 |
| 367 | $10^8$ | 0·00517 | 0·00602 | 0·00588 | 0·00656 | 0·00544 | 0·00552 | 0·00546 | 0·00531 | 0·00656 |
| 503 | $10^7$ | 0·01788 | 0·01457 | 0·02023 | 0·03098 | 0·03022 | 0·02548 | 0·02053 | 0·01279 | 0·03098 |
| 501 | $10^8$ | 0·00471 | 0·00482 | 0·00565 | 0·00637 | 0·00741 | 0·00837 | 0·00463 | 0·00597 | 0·00837 |
| 751 | $10^{10}$ | 0·00722 | 0·00807 | 0·00709 | 0·01268 | 0·01114 | 0·00939 | 0·01042 | 0·00924 | 0·01268 |
| 546 | $10^8$ | 0·00967 | 0·00901 | 0·01000 | 0·00952 | 0·01374 | 0·01877 | 0·01196 | 0·01157 | 0·01877 |
| 945 | $10^8$ | 0·01601 | 0·01628 | 0·01381 | 0·01474 | 0·01442 | 0·01430 | 0·01704 | 0·01051 | 0·01704 |
| 802 | $10^{10}$ | 0·01187 | 0·01227 | 0·01124 | 0·01123 | | | 0·01357 | 0·01256 | 0·01357 |
| 960 | $10^8$ | 0·01614 | 0·01229 | 0·01416 | 0·02266 | 0·02323 | 0·01906 | 0·01161 | 0·01334 | 0·02323 |
| 375 | $10^5$ | 0·01054 | 0·00950 | 0·01079 | 0·01215 | 0·01005 | 0·00975 | 0·01166 | 0·01532 | 0·01532 |
| 350 | $10^5$ | 0·00891 | 0·00921 | 0·00863 | 0·00905 | 0·01686 | 0·03347 | 0·02929 | 0·05878 | 0·05878 |
| 1111 | $10^6$ | 0·02641 | 0·02250 | 0·01791 | 0·01732 | 0·01640 | 0·01876 | 0·01739 | 0·01833 | 0·02641 |
| 576 | $10^7$ | 0·00514 | 0·00634 | 0·00947 | 0·01428 | 0·01195 | 0·00761 | 0·00626 | 0·00605 | 0·01428 |
| 520 | $10^7$ | 0·00822 | 0·00927 | 0·01357 | 0·00974 | 0·00946 | 0·00835 | 0·01208 | 0·01136 | 0·01357 |

| Primary fold change TcpA IgA | | | | | | | | | |
|---|---|---|---|---|---|---|---|---|---|
| Dose (CFU) | D1 | D4 | D7 | D15 | D29 | D57 | D85 | D180 | max FC |
| $10^8$ | 1·00 | 1·29 | 1·33 | 3·08 | 1·61 | 1·52 | 1·60 | 1·19 | 3·08 |
| $10^8$ | 1·00 | 0·75 | 0·82 | 0·80 | 0·56 | 0·71 | 0·71 | 0·76 | 0·82 |
| $10^7$ | 1·00 | 1·06 | 1·44 | 2·29 | 1·96 | 1·49 | 1·66 | 0·87 | 2·29 |
| $10^7$ | 1·00 | 0·83 | 1·00 | 1·24 | 1·12 | 1·13 | 1·11 | 1·34 | 1·34 |
| $10^8$ | 1·00 | 1·00 | 1·06 | 1·01 | 1·29 | 1·03 | 0·95 | 0·82 | 1·29 |
| $10^8$ | 1·00 | 0·84 | 0·94 | 0·81 | 1·16 | 1·06 | 1·05 | 0·92 | 1·16 |
| $10^7$ | 1·00 | 1·30 | 1·45 | 1·76 | 1·52 | 1·69 | 2·61 | 1·14 | 2·61 |
| $10^8$ | 1·00 | 1·16 | 1·14 | 1·27 | 1·05 | 1·07 | 1·06 | 1·03 | 1·27 |
| $10^7$ | 1·00 | 0·81 | 1·13 | 1·73 | 1·69 | 1·43 | 1·15 | 0·72 | 1·73 |
| $10^8$ | 1·00 | 1·02 | 1·20 | 1·35 | 1·57 | 1·78 | 0·98 | 1·27 | 1·78 |
| $10^{10}$ | 1·00 | 1·12 | 0·98 | 1·76 | 1·54 | 1·30 | 1·44 | 1·28 | 1·76 |
| $10^8$ | 1·00 | 0·93 | 1·03 | 0·98 | 1·42 | 1·94 | 1·24 | 1·20 | 1·94 |
| $10^8$ | 1·00 | 1·02 | 0·86 | 0·92 | 0·90 | 0·89 | 1·06 | 0·66 | 1·06 |
| $10^{10}$ | 1·00 | 1·03 | 0·95 | 0·95 | | | 1·14 | 1·06 | 1·14 |
| $10^8$ | 1·00 | 0·76 | 0·88 | 1·40 | 1·44 | 1·18 | 0·72 | 0·83 | 1·44 |
| $10^5$ | 1·00 | 0·90 | 1·02 | 1·15 | 0·95 | 0·93 | 1·11 | 1·45 | 1·45 |
| $10^5$ | 1·00 | 1·03 | 0·97 | 1·02 | 1·89 | 3·76 | 3·29 | 6·60 | 6·60 |
| $10^6$ | 1·00 | 0·85 | 0·68 | 0·66 | 0·62 | 0·71 | 0·66 | 0·69 | 0·85 |
| $10^7$ | 1·00 | 1·23 | 1·84 | 2·78 | 2·32 | 1·48 | 1·22 | 1·18 | 2·78 |
| $10^7$ | 1·00 | 1·13 | 1·65 | 1·18 | 1·15 | 1·02 | 1·47 | 1·38 | 1·65 |

**Table S6: Isotype- and antigen-specific immune responses in lymphocyte supernatants after primary and booster vaccination.**
Blue: no serum sample; light grey: below limit of detection.

| Boost Inaba OSP IgM (RAU) | | | | | | | | |
|---|---|---|---|---|---|---|---|---|
| **Dose interval (days)** | **Dose (CFU)** | **D1** | **D4** | **D7** | **D15** | **D29** | **D180** | **max** |
| 389 | $2\times10^7$ | | 2·73E-06 | 4·58E-04 | 1·64E-07 | | 7·58E-07 | 4·58E-04 |
| 405 | $2\times10^7$ | | | 1·96E-03 | 6·42E-07 | | 4·40E-07 | 1·96E-03 |
| 322 | $2\times10^7$ | | | 8·15E-04 | | 2·19E-06 | 5·81E-07 | 8·15E-04 |
| 418 | $2\times10^7$ | | | 3·81E-04 | | | 5·81E-07 | 3·81E-04 |
| 378 | $2\times10^7$ | | | 4·79E-03 | 8·18E-06 | 3·73E-06 | 1·18E-06 | 4·79E-03 |
| 399 | $2\times10^7$ | | | 5·51E-06 | | | 1·97E-07 | 5·51E-06 |
| 568 | $2\times10^7$ | | | 1·13E-03 | | | 4·05E-07 | 1·13E-03 |
| 367 | $2\times10^7$ | | | 3·75E-05 | | | 4·05E-07 | 3·75E-05 |
| 503 | $2\times10^7$ | | | 5·23E-06 | | | 1·26E-06 | 5·23E-06 |
| 501 | $2\times10^7$ | | | 4·09E-03 | | | 4·90E-07 | 4·09E-03 |
| 751 | $2\times10^7$ | 7·58E-07 | 4·05E-07 | 2·25E-03 | 3·64E-06 | 1·40E-06 | 6·63E-07 | 2·25E-03 |
| 546 | $2\times10^7$ | 4·75E-07 | 7·58E-07 | 5·36E-05 | 5·46E-06 | 4·15E-06 | 1·08E-06 | 5·36E-05 |
| 945 | $2\times10^7$ | 2·66E-07 | 6·16E-07 | 4·24E-03 | 9·71E-07 | 6·87E-07 | 5·58E-07 | 4·24E-03 |
| 802 | $2\times10^7$ | 7·58E-07 | 3·70E-07 | 2·91E-05 | 6·16E-07 | 1·29E-06 | 3·93E-07 | 2·91E-05 |
| 960 | $2\times10^7$ | 5·92E-07 | 4·75E-07 | 5·82E-03 | 7·06E-06 | 2·16E-06 | 8·48E-07 | 5·82E-03 |
| 375 | $2\times10^7$ | 4·40E-07 | 4·75E-07 | 6·35E-04 | 2·63E-06 | 5·46E-07 | 1·04E-06 | 6·35E-04 |
| 350 | $2\times10^7$ | 6·51E-07 | 4·75E-07 | 5·12E-05 | 8·64E-07 | 6·87E-07 | 2·94E-06 | 5·12E-05 |
| 1111 | $2\times10^7$ | | 1·97E-07 | 3·01E-07 | 1·29E-07 | 3·71E-06 | 7·73E-07 | 3·71E-06 |
| 576 | $2\times10^7$ | 3·01E-07 | 5·53E-06 | 4·49E-03 | 3·70E-07 | 4·75E-07 | 1·00E-06 | 4·49E-03 |
| 520 | $2\times10^7$ | 3·30E-04 | 1·79E-05 | 9·71E-07 | 2·66E-06 | 1·54E-05 | 6·99E-07 | 3·30E-04 |

| Primary Inaba OSP IgM (RAU) | | | | | | | |
|---|---|---|---|---|---|---|---|
| **Dose interval (days)** | **Dose (CFU)** | **D1** | **D7** | **D15** | **D29** | **D180** | **max** |
| 389 | $10^8$ | 1·37E-07 | 3·79E-06 | 3·71E-06 | 7·41E-06 | | 7·41E-06 |
| 405 | $10^8$ | 1·05E-07 | 1·97E-06 | 1·77E-06 | 6·28E-07 | | 1·97E-06 |
| 322 | $10^7$ | 3·14E-07 | 9·34E-03 | 7·79E-06 | 3·39E-06 | 1·62E-06 | 9·34E-03 |
| 418 | $10^7$ | 5·24E-07 | 1·24E-02 | 2·84E-05 | 7·25E-05 | 9·74E-08 | 1·24E-02 |
| 378 | $10^8$ | | 3·65E-03 | 2·15E-06 | 5·60E-05 | | 3·65E-03 |
| 399 | $10^8$ | 1·02E-06 | 1·76E-01 | 4·79E-04 | 2·22E-05 | 4·63E-05 | 1·76E-01 |
| 568 | $10^7$ | 1·78E-04 | 1·14E-02 | 7·82E-04 | 6·22E-04 | 3·11E-04 | 1·14E-02 |
| 367 | $10^8$ | 1·68E-06 | 4·46E-05 | 3·70E-05 | 5·80E-06 | 2·76E-06 | 4·46E-05 |
| 503 | $10^7$ | 2·91E-07 | 1·78E-03 | 1·71E-05 | 8·33E-05 | | 1·78E-03 |
| 501 | $10^8$ | 6·28E-07 | 4·45E-03 | 1·93E-05 | 4·23E-06 | 1·24E-06 | 4·45E-03 |
| 751 | $10^{10}$ | | 2·59E-02 | 1·64E-05 | 8·76E-05 | 5·30E-08 | 2·59E-02 |
| 546 | $10^8$ | 6·81E-07 | 1·43E-04 | 9·77E-05 | 1·92E-05 | | 1·43E-04 |
| 945 | $10^8$ | | 2·02E-05 | | 1·63E-07 | | 2·02E-05 |
| 802 | $10^{10}$ | | 1·13E-01 | 2·52E-05 | | | 1·13E-01 |
| 960 | $10^8$ | | 4·18E-03 | 2·60E-06 | 3·73E-06 | 1·45E-06 | 4·18E-03 |
| 375 | $10^5$ | | 3·89E-05 | 1·55E-05 | 4·24E-06 | 1·01E-06 | 3·89E-05 |
| 350 | $10^5$ | 6·55E-07 | 9·26E-04 | 5·88E-06 | 1·55E-06 | 2·01E-06 | 9·26E-04 |
| 1111 | $10^6$ | | 9·23E-05 | | | | 9·23E-05 |
| 576 | $10^7$ | 1·32E-06 | 1·48E-03 | | 8·46E-07 | 1·28E-07 | 1·48E-03 |
| 520 | $10^7$ | | 3·07E-04 | 4·77E-04 | 6·37E-06 | 7·08E-07 | 4·77E-04 |

| | |
|---|---|
| | no serum sample |
| | below limit of detection |

| Boost Inaba IgG (RAU) | | | | | | | | |
|---|---|---|---|---|---|---|---|---|
| **Dose interval (days)** | **Dose (CFU)** | **D1** | **D4** | **D7** | **D15** | **D29** | **D180** | **max** |
| 389 | $2\times10^{7}$ | 1·78E-07 | 5·74E-06 | | 2·64E-05 | 2·10E-05 | 1·27E-06 | 2·64E-05 |
| 405 | $2\times10^{7}$ | 3·86E-06 | | 7·10E-03 | 1·41E-05 | 1·10E-05 | | 7·10E-03 |
| 322 | $2\times10^{7}$ | 5·74E-06 | 2·34E-06 | 7·78E-04 | 7·65E-06 | | 1·91E-06 | 7·78E-04 |
| 418 | $2\times10^{7}$ | | | 2·43E-03 | 1·78E-07 | 8·33E-06 | 5·69E-06 | 2·43E-03 |
| 378 | $2\times10^{7}$ | | 3·57E-06 | 9·24E-03 | 4·70E-06 | 2·66E-06 | 1·49E-06 | 9·24E-03 |
| 399 | $2\times10^{7}$ | | 2·66E-06 | 3·57E-06 | 5·74E-06 | 3·86E-06 | 3·69E-06 | 5·74E-06 |
| 568 | $2\times10^{7}$ | 1·78E-07 | | | 8·17E-05 | 4·77E-05 | 4·25E-06 | 8·17E-05 |
| 367 | $2\times10^{7}$ | 3·86E-06 | 3·28E-05 | 3·68E-04 | 1·22E-05 | 8·55E-06 | 3·68E-05 | 3·68E-04 |
| 503 | $2\times10^{7}$ | 3·86E-06 | 1·66E-06 | 3·67E-05 | 4·70E-06 | | 3·50E-06 | 3·67E-05 |
| 501 | $2\times10^{7}$ | 2·97E-06 | 2·01E-05 | 1·65E-02 | 1·18E-05 | 1·58E-05 | 1·35E-05 | 1·65E-02 |
| 751 | $2\times10^{7}$ | 5·49E-05 | 1·96E-04 | 3·96E-01 | 1·28E-04 | 1·11E-04 | 2·05E-05 | 3·96E-01 |
| 546 | $2\times10^{7}$ | 4·79E-06 | 8·08E-06 | 3·37E-04 | 1·07E-05 | 6·04E-06 | 5·45E-06 | 3·37E-04 |
| 945 | $2\times10^{7}$ | 5·33E-06 | 3·87E-05 | 8·63E-02 | 1·21E-04 | 5·50E-05 | 4·25E-05 | 8·63E-02 |
| 802 | $2\times10^{7}$ | 8·12E-07 | | 1·01E-04 | 1·36E-05 | 1·35E-05 | 1·39E-05 | 1·01E-04 |
| 960 | $2\times10^{7}$ | 3·28E-06 | 3·12E-06 | 8·77E-03 | 5·95E-05 | 1·00E-05 | 2·95E-05 | 8·77E-03 |
| 375 | $2\times10^{7}$ | 1·49E-06 | 3·69E-06 | 1·37E-03 | 1·49E-05 | 4·61E-06 | 1·97E-05 | 1·37E-03 |
| 350 | $2\times10^{7}$ | 7·75E-06 | 5·29E-05 | 6·56E-03 | 1·50E-05 | 9·39E-06 | 3·06E-05 | 6·56E-03 |
| 1111 | $2\times10^{7}$ | | 3·09E-07 | 1·33E-04 | 1·91E-06 | 7·07E-06 | 1·35E-05 | 1·33E-04 |
| 576 | $2\times10^{7}$ | 5·69E-07 | 5·69E-07 | 3·94E-03 | 2·92E-06 | 4·25E-06 | 1·22E-05 | 3·94E-03 |
| 520 | $2\times10^{7}$ | 3·09E-04 | 2·10E-05 | 9·72E-06 | 3·68E-05 | 1·47E-05 | 2·76E-05 | 3·09E-04 |

| Primary Inaba OSP IgG (RAU) | | | | | | | |
|---|---|---|---|---|---|---|---|
| **Dose interval (days)** | **Dose (CFU)** | **D1** | **D7** | **D15** | **D29** | **D180** | **max** |
| 389 | $10^{8}$ | 1·07E-05 | 2·93E-06 | 2·93E-06 | 1·24E-05 | | 1·24E-05 |
| 405 | $10^{8}$ | 6·43E-06 | | 6·43E-06 | 1·29E-05 | | 1·29E-05 |
| 322 | $10^{7}$ | | | | | | |
| 418 | $10^{7}$ | 1·73E-05 | 2·04E-05 | 8·10E-06 | 1·42E-05 | | 2·04E-05 |
| 378 | $10^{8}$ | | | | | | |
| 399 | $10^{8}$ | | 1·11E-04 | 2·58E-06 | 1·92E-06 | 9·73E-05 | 1·11E-04 |
| 568 | $10^{7}$ | 5·12E-04 | 7·11E-03 | 5·12E-04 | 1·25E-04 | 9·02E-04 | 7·11E-03 |
| 367 | $10^{8}$ | | | | | 1·68E-04 | 1·68E-04 |
| 503 | $10^{7}$ | 8·95E-06 | 1·68E-05 | 1·51E-05 | 2·93E-06 | 4·40E-05 | 4·40E-05 |
| 501 | $10^{8}$ | 1·92E-06 | 2·70E-05 | | 1·31E-06 | | 2·70E-05 |
| 751 | $10^{10}$ | 1·01E-05 | 8·85E-06 | 1·01E-05 | | 2·83E-05 | 2·83E-05 |
| 546 | $10^{8}$ | 1·02E-05 | 1·59E-05 | 1·46E-05 | 3·48E-05 | 7·87E-06 | 3·48E-05 |
| 945 | $10^{8}$ | | | 1·27E-05 | | 4·26E-05 | 4·26E-05 |
| 802 | $10^{10}$ | 3·34E-06 | 1·69E-06 | | | 4·85E-06 | 4·85E-06 |
| 960 | $10^{8}$ | | 2·64E-04 | 5·84E-06 | | | 2·64E-04 |
| 375 | $10^{5}$ | | | 7·33E-05 | | 2·45E-05 | 7·33E-05 |
| 350 | $10^{5}$ | | | 7·87E-06 | | 4·94E-05 | 4·94E-05 |
| 1111 | $10^{6}$ | 1·27E-05 | 3·76E-05 | | | | 3·76E-05 |
| 576 | $10^{7}$ | 4·04E-06 | 1·29E-05 | 6·43E-06 | 4·43E-06 | | 1·29E-05 |
| 520 | $10^{7}$ | | | | | | |

| Boost Inaba IgA (RAU) | | | | | | | | |
|---|---|---|---|---|---|---|---|---|
| **Dose interval (days)** | **Dose (CFU)** | **D1** | **D4** | **D7** | **D15** | **D29** | **D180** | **max** |
| 389 | $2\times10^{7}$ | 1·46E-07 | | 1·09E-01 | 2·12E-05 | | 2·83E-06 | 1·09E-01 |
| 405 | $2\times10^{7}$ | | 1·83E-07 | 7·17E-02 | 2·82E-06 | 1·34E-06 | 4·70E-06 | 7·17E-02 |
| 322 | $2\times10^{7}$ | 2·13E-05 | | 5·02E-02 | 3·53E-06 | 3·13E-07 | 3·10E-06 | 5·02E-02 |
| 418 | $2\times10^{7}$ | | | 4·64E-03 | | 3·86E-07 | 4·04E-06 | 4·64E-03 |
| 378 | $2\times10^{7}$ | 5·75E-05 | 4·34E-05 | 8·48E-01 | 1·44E-04 | 3·58E-05 | 3·78E-05 | 8·48E-01 |
| 399 | $2\times10^{7}$ | | 2·67E-07 | 7·13E-06 | 1·78E-06 | 1·83E-07 | 6·02E-06 | 7·13E-06 |
| 568 | $2\times10^{7}$ | | | 1·72E-02 | 1·78E-06 | | 5·60E-06 | 1·72E-02 |
| 367 | $2\times10^{7}$ | 2·45E-06 | 1·06E-05 | 2·96E-04 | 4·77E-06 | 2·25E-06 | 6·41E-06 | 2·96E-04 |
| 503 | $2\times10^{7}$ | 1·25E-05 | 4·12E-07 | 1·16E-04 | 2·36E-05 | 2·03E-07 | 2·93E-05 | 1·16E-04 |
| 501 | $2\times10^{7}$ | 2·03E-07 | | 3·61E-02 | 2·53E-05 | | 1·06E-05 | 3·61E-02 |
| 751 | $2\times10^{7}$ | 3·30E-06 | 2·16E-05 | 3·33E-02 | 1·42E-04 | 5·30E-05 | 1·40E-05 | 3·33E-02 |
| 546 | $2\times10^{7}$ | 2·18E-05 | 5·25E-05 | 1·21E-03 | 9·14E-05 | 6·80E-06 | | 1·21E-03 |
| 945 | $2\times10^{7}$ | 6·87E-06 | 1·83E-05 | 6·75E-02 | 3·28E-05 | 9·10E-06 | 4·14E-06 | 6·75E-02 |
| 802 | $2\times10^{7}$ | 2·09E-06 | 9·88E-07 | 5·34E-04 | 2·09E-06 | 1·90E-06 | 2·60E-06 | 5·34E-04 |
| 960 | $2\times10^{7}$ | 4·99E-06 | 1·40E-05 | 2·10E-02 | 2·38E-05 | 2·60E-06 | 3·12E-05 | 2·10E-02 |
| 375 | $2\times10^{7}$ | 9·13E-07 | 7·07E-06 | 7·59E-03 | 3·34E-05 | 2·40E-05 | 1·69E-05 | 7·59E-03 |
| 350 | $2\times10^{7}$ | 5·09E-06 | 8·85E-05 | 2·88E-03 | 3·92E-06 | 1·29E-05 | 2·02E-05 | 2·88E-03 |
| 1111 | $2\times10^{7}$ | 9·50E-07 | 1·86E-06 | 7·02E-05 | 2·21E-06 | 2·68E-06 | 1·31E-05 | 7·02E-05 |
| 576 | $2\times10^{7}$ | 1·21E-06 | 4·35E-06 | 7·70E-02 | 4·23E-06 | 3·46E-06 | 4·05E-06 | 7·70E-02 |
| 520 | $2\times10^{7}$ | 1·17E-03 | 2·11E-04 | 2·63E-05 | 2·58E-05 | 2·19E-04 | 2·20E-05 | 1·17E-03 |

| Primary Inaba OSP IgA (RAU) | | | | | | | |
|---|---|---|---|---|---|---|---|
| **Dose interval (days)** | **Dose (CFU)** | **D1** | **D7** | **D15** | **D29** | **D180** | **max** |
| 389 | $10^{8}$ | 4·32E-06 | 7·38E-07 | 7·19E-06 | 5·64E-06 | | 7·19E-06 |
| 405 | $10^{8}$ | 2·43E-06 | 1·63E-06 | | 2·43E-06 | | 2·43E-06 |
| 322 | $10^{7}$ | 3·71E-07 | 1·71E-03 | 4·01E-06 | 2·28E-06 | 1·81E-06 | 1·71E-03 |
| 418 | $10^{7}$ | 2·16E-06 | 1·26E-04 | 4·50E-05 | 3·20E-04 | 1·93E-05 | 3·20E-04 |
| 378 | $10^{8}$ | 1·04E-06 | 8·40E-05 | 2·19E-06 | 1·84E-05 | 3·45E-06 | 8·40E-05 |
| 399 | $10^{8}$ | 1·44E-06 | 1·12E-03 | 4·58E-05 | 2·22E-06 | 2·36E-05 | 1·12E-03 |
| 568 | $10^{7}$ | | 2·06E-02 | 5·60E-03 | 8·28E-03 | 9·81E-06 | 2·06E-02 |
| 367 | $10^{8}$ | 1·10E-06 | 4·23E-07 | 2·28E-06 | 1·45E-06 | 7·46E-06 | 7·46E-06 |
| 503 | $10^{7}$ | 1·57E-06 | 1·75E-04 | 1·42E-05 | 3·83E-05 | 2·64E-05 | 1·75E-04 |
| 501 | $10^{8}$ | 1·10E-06 | 9·67E-06 | 7·51E-06 | 9·18E-06 | 8·38E-06 | 9·67E-06 |
| 751 | $10^{10}$ | 2·16E-07 | 4·97E-04 | 1·08E-04 | 2·56E-04 | 1·54E-04 | 4·97E-04 |
| 546 | $10^{8}$ | 1·26E-06 | 4·89E-04 | 2·50E-05 | 7·04E-05 | 1·18E-05 | 4·89E-04 |
| 945 | $10^{8}$ | 9·06E-07 | 2·90E-06 | 1·41E-06 | 2·45E-06 | 5·26E-06 | 5·26E-06 |
| 802 | $10^{10}$ | 5·97E-07 | 3·87E-06 | 2·52E-05 | | 2·57E-05 | 2·57E-05 |
| 960 | $10^{8}$ | 1·72E-06 | 9·70E-05 | 1·28E-06 | 1·22E-06 | 9·29E-06 | 9·70E-05 |
| 375 | $10^{5}$ | 1·57E-06 | 3·10E-06 | 2·25E-05 | 5·78E-06 | 5·45E-05 | 5·45E-05 |
| 350 | $10^{5}$ | 2·28E-06 | 7·36E-06 | 3·50E-06 | 1·28E-05 | 8·93E-05 | 8·93E-05 |
| 1111 | $10^{6}$ | 6·58E-07 | 1·14E-02 | 1·28E-05 | 5·57E-06 | 2·21E-06 | 1·14E-02 |
| 576 | $10^{7}$ | 8·66E-07 | 3·62E-06 | 1·81E-06 | 2·16E-06 | 5·84E-07 | 3·62E-06 |
| 520 | $10^{7}$ | 2·93E-06 | 6·73E-06 | 2·40E-06 | 1·19E-05 | 2·09E-05 | 2·09E-05 |

| Boost Ogawa IgM (RAU) | | | | | | | | |
|---|---|---|---|---|---|---|---|---|
| Dose interval (days) | Dose (CFU) | D1 | D4 | D7 | D15 | D29 | D180 | max |
| 389 | $2\times10^{7}$ | 1·79E-06 | | 2·20E-03 | 2·83E-06 | 1·31E-06 | 1·63E-06 | 2·20E-03 |
| 405 | $2\times10^{7}$ | 3·69E-06 | 1·08E-07 | 4·59E-03 | 4·91E-05 | 9·44E-07 | 9·23E-07 | 4·59E-03 |
| 322 | $2\times10^{7}$ | | 2·04E-07 | 2·52E-04 | 1·89E-06 | 1·08E-07 | 1·86E-06 | 2·52E-04 |
| 418 | $2\times10^{7}$ | 1·50E-06 | | 1·83E-04 | | 1·53E-07 | 1·63E-06 | 1·83E-04 |
| 378 | $2\times10^{7}$ | | 1·53E-07 | 3·14E-03 | 4·24E-06 | 2·51E-06 | 2·91E-06 | 3·14E-03 |
| 399 | $2\times10^{7}$ | 1·08E-07 | 4·79E-06 | 1·01E-05 | 6·90E-07 | 1·50E-06 | 1·47E-06 | 1·01E-05 |
| 568 | $2\times10^{7}$ | 1·53E-07 | 8·57E-07 | 1·53E-03 | 1·89E-06 | 5·56E-06 | 3·23E-06 | 1·53E-03 |
| 367 | $2\times10^{7}$ | 2·04E-07 | 1·53E-07 | 2·51E-05 | | | 1·63E-06 | 2·51E-05 |
| 503 | $2\times10^{7}$ | | 9·44E-07 | 4·02E-06 | 1·53E-07 | 2·61E-07 | 1·00E-06 | 4·02E-06 |
| 501 | $2\times10^{7}$ | | | 3·86E-03 | 1·21E-06 | 3·04E-06 | 1·49E-06 | 3·86E-03 |
| 751 | $2\times10^{7}$ | 1·94E-06 | 2·58E-06 | 1·84E-03 | 7·41E-06 | 2·02E-06 | 3·70E-06 | 1·84E-03 |
| 546 | $2\times10^{7}$ | 1·78E-06 | 2·99E-06 | 1·42E-04 | 2·18E-05 | 2·99E-05 | 4·01E-05 | 1·42E-04 |
| 945 | $2\times10^{7}$ | 2·10E-06 | 1·15E-06 | 8·14E-03 | 2·50E-06 | 2·58E-06 | 1·48E-05 | 8·14E-03 |
| 802 | $2\times10^{7}$ | 1·08E-06 | 1·94E-06 | 2·32E-05 | 1·71E-06 | 3·24E-06 | 7·46E-07 | 2·32E-05 |
| 960 | $2\times10^{7}$ | 1·94E-06 | 1·63E-06 | 2·79E-03 | 2·50E-06 | 2·02E-06 | 2·39E-06 | 2·79E-03 |
| 375 | $2\times10^{7}$ | 1·63E-06 | 2·10E-06 | 1·05E-03 | 4·45E-06 | 1·47E-06 | 2·20E-06 | 1·05E-03 |
| 350 | $2\times10^{7}$ | 7·70E-07 | 1·23E-06 | 1·83E-05 | 2·75E-06 | 2·42E-06 | 1·13E-05 | 1·83E-05 |
| 1111 | $2\times10^{7}$ | 3·24E-07 | 1·00E-06 | 1·15E-06 | 2·10E-06 | 1·00E-06 | 3·81E-06 | 3·81E-06 |
| 576 | $2\times10^{7}$ | 2·58E-06 | 2·42E-06 | 2·11E-03 | 6·19E-07 | 9·56E-06 | 3·08E-06 | 2·11E-03 |
| 520 | $2\times10^{7}$ | 2·34E-04 | 6·92E-06 | 1·63E-06 | 2·26E-06 | 6·92E-06 | 1·84E-06 | 2·34E-04 |

| Primary Ogawa OSP IgM (RAU) | | | | | | | |
|---|---|---|---|---|---|---|---|
| Dose interval (days) | Dose (CFU) | D1 | D7 | D15 | D29 | D180 | max |
| 389 | $10^{8}$ | 2·75E-07 | 1·48E-04 | 1·45E-05 | 4·86E-08 | | 1·48E-04 |
| 405 | $10^{8}$ | | 2·18E-04 | 1·62E-06 | 2·75E-07 | | 2·18E-04 |
| 322 | $10^{7}$ | | 7·81E-05 | | 9·66E-08 | | 7·81E-05 |
| 418 | $10^{7}$ | 1·01E-06 | 5·80E-06 | | 1·81E-07 | | 5·80E-06 |
| 378 | $10^{8}$ | | 1·58E-02 | 2·51E-05 | 5·85E-05 | | 1·58E-02 |
| 399 | $10^{8}$ | 7·43E-08 | 2·15E-01 | 5·79E-04 | 3·05E-05 | 5·55E-05 | 2·15E-01 |
| 568 | $10^{7}$ | | 9·57E-01 | 6·64E-04 | | 5·24E-04 | 9·57E-01 |
| 367 | $10^{8}$ | | 3·82E-03 | 8·24E-05 | 1·63E-06 | | 3·82E-03 |
| 503 | $10^{7}$ | | 1·75E-03 | 2·22E-05 | 2·59E-06 | | 1·75E-03 |
| 501 | $10^{8}$ | 1·05E-07 | 1·34E-03 | 2·75E-07 | 1·01E-06 | | 1·34E-03 |
| 751 | $10^{10}$ | | 3·09E-02 | 1·30E-05 | 1·41E-05 | 1·25E-07 | 3·09E-02 |
| 546 | $10^{8}$ | 4·43E-07 | 7·74E-03 | 2·42E-05 | 6·15E-06 | | 7·74E-03 |
| 945 | $10^{8}$ | 5·88E-08 | 3·29E-04 | 1·25E-05 | 2·96E-06 | | 3·29E-04 |
| 802 | $10^{10}$ | | 2·40E-03 | 8·05E-06 | | 5·25E-06 | 2·40E-03 |
| 960 | $10^{8}$ | 1·37E-06 | 8·91E-04 | 2·23E-05 | 1·23E-05 | 3·11E-07 | 8·91E-04 |
| 375 | $10^{5}$ | | 8·01E-06 | | 5·27E-05 | 1·31E-06 | 5·27E-05 |
| 350 | $10^{5}$ | | | | | 1·71E-06 | 1·71E-06 |
| 1111 | $10^{6}$ | 7·88E-08 | 1·61E-04 | | | | 1·61E-04 |
| 576 | $10^{7}$ | 1·81E-07 | 1·81E-03 | 8·67E-06 | | | 1·81E-03 |
| 520 | $10^{7}$ | | 2·24E-04 | 3·66E-05 | 8·47E-06 | | 2·24E-04 |

| Boost Ogawa IgG (RAU) | | | | | | | | |
|---|---|---|---|---|---|---|---|---|
| **Dose interval (days)** | **Dose (CFU)** | **D1** | **D4** | **D7** | **D15** | **D29** | **D180** | **max** |
| 389 | $2\times10^7$ | 2·96E-06 | | 7·13E-03 | 3·74E-06 | | 1·61E-06 | 7·13E-03 |
| 405 | $2\times10^7$ | 2·19E-06 | 5·86E-06 | 2·06E-04 | 3·35E-06 | 2·58E-06 | 2·66E-06 | 2·06E-04 |
| 322 | $2\times10^7$ | 6·81E-06 | 2·58E-06 | 1·07E-04 | 2·00E-06 | 2·00E-06 | 1·96E-06 | 1·07E-04 |
| 418 | $2\times10^7$ | 3·55E-06 | 2·19E-06 | 4·24E-05 | 3·74E-06 | 3·55E-06 | 3·02E-06 | 4·24E-05 |
| 378 | $2\times10^7$ | | | 2·58E-04 | 4·71E-06 | 2·39E-06 | 2·84E-06 | 2·58E-04 |
| 399 | $2\times10^7$ | 4·90E-06 | 3·35E-06 | 4·13E-06 | 2·19E-06 | 4·71E-06 | 1·79E-06 | 4·90E-06 |
| 568 | $2\times10^7$ | | 5·67E-06 | 3·16E-03 | 7·18E-06 | 7·00E-06 | 1·96E-06 | 3·16E-03 |
| 367 | $2\times10^7$ | 5·09E-06 | 2·59E-05 | 3·37E-04 | 7·93E-06 | 5·09E-06 | 2·06E-05 | 3·37E-04 |
| 503 | $2\times10^7$ | | | 3·55E-06 | 7·42E-07 | 2·77E-06 | 3·19E-06 | 3·55E-06 |
| 501 | $2\times10^7$ | 1·63E-06 | 1·81E-06 | 1·45E-03 | 1·44E-06 | 1·44E-06 | 9·10E-07 | 1·45E-03 |
| 751 | $2\times10^7$ | 7·11E-06 | 2·78E-05 | 1·85E-03 | 3·58E-05 | 3·55E-06 | 2·51E-06 | 1·85E-03 |
| 546 | $2\times10^7$ | 2·49E-06 | 6·93E-06 | 2·89E-05 | 2·84E-06 | 5·68E-06 | 2·19E-06 | 2·89E-05 |
| 945 | $2\times10^7$ | 4·52E-06 | 4·97E-06 | 1·26E-02 | 1·27E-05 | 6·48E-06 | 7·82E-07 | 1·26E-02 |
| 802 | $2\times10^7$ | 2·31E-06 | 1·26E-06 | 7·29E-06 | 2·75E-06 | 2·31E-06 | 5·78E-07 | 7·29E-06 |
| 960 | $2\times10^7$ | 3·90E-06 | 1·96E-06 | 5·13E-04 | 8·01E-06 | 4·70E-06 | 1·97E-06 | 5·13E-04 |
| 375 | $2\times10^7$ | 3·19E-06 | 4·26E-06 | 1·00E-03 | 8·90E-06 | 2·49E-06 | 8·94E-07 | 1·00E-03 |
| 350 | $2\times10^7$ | 2·93E-06 | 1·98E-05 | 9·32E-04 | 6·75E-06 | 5·23E-06 | 1·41E-06 | 9·32E-04 |
| 1111 | $2\times10^7$ | 2·75E-06 | 2·58E-06 | 2·84E-06 | 1·61E-06 | 2·31E-06 | 7·66E-07 | 2·84E-06 |
| 576 | $2\times10^7$ | 1·18E-06 | 3·90E-06 | 2·15E-04 | 2·66E-06 | 1·35E-06 | 9·74E-07 | 2·15E-04 |
| 520 | $2\times10^7$ | 6·99E-05 | 3·72E-06 | 4·08E-06 | 1·87E-06 | 7·02E-06 | 2·14E-06 | 6·99E-05 |

| Primary Ogawa OSP IgG (RAU) | | | | | | | |
|---|---|---|---|---|---|---|---|
| **Dose interval (days)** | **Dose (CFU)** | **D1** | **D7** | **D15** | **D29** | **D180** | **max** |
| 389 | $10^8$ | 3·40E-06 | 7·37E-06 | | 1·82E-05 | | 1·82E-05 |
| 405 | $10^8$ | | | | | | |
| 322 | $10^7$ | | | 2·10E-06 | | | 2·10E-06 |
| 418 | $10^7$ | | 7·71E-06 | | 5·57E-06 | | 7·71E-06 |
| 378 | $10^8$ | | 5·31E-05 | | 4·63E-07 | | 5·31E-05 |
| 399 | $10^8$ | | 5·14E-05 | 7·03E-06 | | 4·72E-05 | 5·14E-05 |
| 568 | $10^7$ | | 7·46E-02 | 4·72E-04 | | | 7·46E-02 |
| 367 | $10^8$ | 2·91E-07 | 2·53E-06 | 3·27E-06 | 6·58E-07 | 9·16E-05 | 9·16E-05 |
| 503 | $10^7$ | 1·30E-05 | 1·45E-05 | 2·86E-06 | 7·71E-06 | 1·10E-06 | 1·45E-05 |
| 501 | $10^8$ | 1·05E-05 | 1·08E-05 | 5·95E-06 | | | 1·08E-05 |
| 751 | $10^{10}$ | 4·34E-06 | 2·53E-06 | 1·33E-06 | | 2·53E-06 | 4·34E-06 |
| 546 | $10^8$ | | 2·14E-05 | 6·68E-06 | 1·27E-05 | | 2·14E-05 |
| 945 | $10^8$ | 3·50E-06 | | 9·92E-08 | 3·50E-06 | | 3·50E-06 |
| 802 | $10^{10}$ | | 3·13E-06 | 1·92E-06 | | | 3·13E-06 |
| 960 | $10^8$ | 5·69E-06 | 1·19E-05 | 2·02E-06 | 2·02E-06 | 1·33E-06 | 1·19E-05 |
| 375 | $10^5$ | | | | | 1·51E-05 | 1·51E-05 |
| 350 | $10^5$ | | | | | 1·14E-05 | 1·14E-05 |
| 1111 | $10^6$ | 6·26E-07 | 4·24E-06 | 1·30E-06 | | 4·97E-06 | 4·97E-06 |
| 576 | $10^7$ | 1·19E-05 | | 8·36E-06 | | | 1·19E-05 |
| 520 | $10^7$ | | 2·10E-06 | | | | 2·10E-06 |

| Boost Ogawa IgA (RAU) | | | | | | | | |
|---|---|---|---|---|---|---|---|---|
| Dose interval (days) | Dose (CFU) | D1 | D4 | D7 | D15 | D29 | D180 | max |
| 389 | $2\times10^7$ | 1·00E-05 | 6·95E-06 | 6·44E-02 | 4·62E-05 | 5·17E-06 | 9·87E-06 | 6·44E-02 |
| 405 | $2\times10^7$ | 7·61E-06 | 5·61E-06 | 2·11E-02 | 3·06E-05 | 1·41E-05 | 2·89E-05 | 2·11E-02 |
| 322 | $2\times10^7$ | 1·24E-05 | 8·39E-06 | 3·53E-03 | 9·04E-06 | 8·60E-06 | 2·01E-05 | 3·53E-03 |
| 418 | $2\times10^7$ | 1·36E-06 | 5·06E-06 | 2·35E-03 | 7·61E-06 | 8·05E-06 | 1·88E-05 | 2·35E-03 |
| 378 | $2\times10^7$ | 5·18E-05 | 5·51E-05 | 3·67E-01 | 1·16E-04 | 2·92E-05 | 2·75E-05 | 3·67E-01 |
| 399 | $2\times10^7$ | 7·78E-07 | 1·47E-05 | 2·32E-05 | 4·72E-06 | 4·95E-06 | 1·85E-05 | 2·32E-05 |
| 568 | $2\times10^7$ | 2·62E-06 | | 3·00E-03 | 7·72E-06 | 6·28E-06 | 3·09E-05 | 3·00E-03 |
| 367 | $2\times10^7$ | 1·02E-05 | 8·17E-06 | 1·13E-04 | 2·62E-06 | 7·83E-06 | 4·43E-05 | 1·13E-04 |
| 503 | $2\times10^7$ | 5·73E-06 | 7·17E-06 | 5·12E-05 | 9·59E-06 | 6·95E-06 | 4·70E-05 | 5·12E-05 |
| 501 | $2\times10^7$ | 3·83E-06 | 2·73E-06 | 1·42E-02 | 6·06E-06 | 2·95E-06 | 4·55E-05 | 1·42E-02 |
| 751 | $2\times10^7$ | 2·01E-05 | 6·06E-05 | 2·34E-02 | 1·55E-04 | 1·09E-04 | 4·79E-05 | 2·34E-02 |
| 546 | $2\times10^7$ | 2·35E-05 | 1·18E-04 | 2·73E-04 | 5·18E-05 | 3·66E-05 | 1·73E-05 | 2·73E-04 |
| 945 | $2\times10^7$ | 3·61E-05 | 6·00E-05 | 1·23E-01 | 4·92E-05 | 3·67E-05 | 2·38E-05 | 1·23E-01 |
| 802 | $2\times10^7$ | 1·15E-05 | 6·93E-06 | 4·07E-04 | 1·27E-05 | 1·30E-05 | 2·89E-05 | 4·07E-04 |
| 960 | $2\times10^7$ | 6·17E-05 | 6·26E-05 | 7·98E-03 | 7·30E-05 | 1·61E-05 | 1·40E-04 | 7·98E-03 |
| 375 | $2\times10^7$ | 9·18E-06 | 4·31E-05 | 7·52E-03 | 1·19E-04 | 4·90E-05 | 8·68E-05 | 7·52E-03 |
| 350 | $2\times10^7$ | 1·79E-05 | 7·30E-05 | 1·91E-03 | 2·03E-05 | 2·16E-05 | 8·76E-05 | 1·91E-03 |
| 1111 | $2\times10^7$ | 9·28E-06 | 1·31E-05 | 3·40E-05 | 1·47E-05 | 1·34E-05 | 6·24E-05 | 6·24E-05 |
| 576 | $2\times10^7$ | 5·58E-06 | 2·12E-05 | 5·37E-03 | 2·20E-05 | 1·52E-05 | 3·10E-05 | 5·37E-03 |
| 520 | $2\times10^7$ | 3·06E-04 | 5·64E-05 | 3·00E-05 | 2·48E-05 | 5·81E-05 | 9·04E-05 | 3·06E-04 |

| Primary Ogawa OSP IgA (RAU) | | | | | | | |
|---|---|---|---|---|---|---|---|
| Dose interval (days) | Dose (CFU) | D1 | D7 | D15 | D29 | D180 | max |
| 389 | $10^8$ | 7·41E-06 | 1·84E-05 | 1·19E-05 | 9·05E-06 | | 1·84E-05 |
| 405 | $10^8$ | 3·57E-06 | 1·74E-05 | 3·07E-06 | 1·53E-05 | 1·47E-05 | 1·74E-05 |
| 322 | $10^7$ | 3·27E-06 | 8·07E-06 | 1·04E-05 | 5·04E-06 | 4·26E-06 | 1·04E-05 |
| 418 | $10^7$ | 3·73E-06 | 6·18E-06 | 6·59E-06 | 2·53E-06 | 6·54E-07 | 6·59E-06 |
| 378 | $10^8$ | 1·60E-06 | 1·32E-01 | 1·68E-04 | 4·79E-04 | 2·32E-06 | 1·32E-01 |
| 399 | $10^8$ | | 8·95E-04 | 5·53E-05 | 3·33E-06 | 3·69E-05 | 8·95E-04 |
| 568 | $10^7$ | 3·25E-06 | 2·94E-05 | 3·65E-06 | 3·25E-06 | 3·25E-06 | 2·94E-05 |
| 367 | $10^8$ | 6·54E-07 | 5·74E-05 | 1·46E-07 | 6·56E-06 | | 5·74E-05 |
| 503 | $10^7$ | 3·48E-07 | 4·91E-04 | 1·60E-05 | 9·47E-06 | 1·52E-05 | 4·91E-04 |
| 501 | $10^8$ | 2·26E-06 | 7·14E-06 | 4·36E-06 | 9·26E-06 | 1·33E-05 | 1·33E-05 |
| 751 | $10^{10}$ | | 6·15E-05 | 3·15E-05 | 7·41E-05 | 1·82E-04 | 1·82E-04 |
| 546 | $10^8$ | 1·37E-06 | 1·00E-02 | 2·48E-05 | 2·93E-06 | | 1·00E-02 |
| 945 | $10^8$ | 5·85E-07 | 3·31E-06 | 2·30E-06 | 2·83E-06 | 2·84E-06 | 3·31E-06 |
| 802 | $10^{10}$ | 5·02E-07 | 5·37E-05 | 3·09E-05 | | 1·09E-05 | 5·37E-05 |
| 960 | $10^8$ | 4·51E-06 | 7·84E-04 | 6·16E-06 | 3·41E-06 | 3·57E-06 | 7·84E-04 |
| 375 | $10^5$ | 3·07E-06 | 2·66E-06 | 7·04E-06 | 8·53E-06 | 7·13E-05 | 7·13E-05 |
| 350 | $10^5$ | 5·62E-06 | 6·95E-06 | 5·04E-06 | 5·04E-06 | 3·12E-05 | 3·12E-05 |
| 1111 | $10^6$ | 1·88E-06 | 1·39E-03 | 3·49E-06 | 5·79E-06 | 4·66E-06 | 1·39E-03 |
| 576 | $10^7$ | 2·86E-06 | 2·71E-03 | 2·76E-06 | 2·23E-05 | 1·23E-06 | 2·71E-03 |
| 520 | $10^7$ | 4·06E-06 | 6·03E-05 | 8·71E-06 | 4·07E-05 | 7·32E-06 | 6·03E-05 |

| Boost CT-B IgM (RAU) | | | | | | | | |
|---|---|---|---|---|---|---|---|---|
| Dose interval (days) | Dose (CFU) | D1 | D4 | D7 | D15 | D29 | D180 | max |
| 389 | $2\times10^{7}$ | 6·68E-05 | 1·69E-05 | 1·33E-05 | 1·61E-04 | 1·33E-05 | 5·04E-05 | 1·61E-04 |
| 405 | $2\times10^{7}$ | 1·83E-04 | 6·18E-05 | 1·17E-03 | 8·21E-05 | 1·45E-04 | 8·23E-05 | 1·17E-03 |
| 322 | $2\times10^{7}$ | 6·82E-04 | 8·68E-04 | 1·90E-03 | 4·38E-04 | 3·99E-04 | 1·14E-03 | 1·90E-03 |
| 418 | $2\times10^{7}$ | 1·40E-04 | 4·25E-05 | 1·14E-04 | | 5·69E-05 | 7·13E-05 | 1·40E-04 |
| 378 | $2\times10^{7}$ | 1·29E-04 | 1·29E-04 | 2·83E-04 | 9·01E-07 | 1·08E-04 | 3·74E-05 | 2·83E-04 |
| 399 | $2\times10^{7}$ | 2·25E-04 | 1·35E-04 | 1·19E-04 | 1·45E-04 | 3·78E-05 | 2·06E-04 | 2·25E-04 |
| 568 | $2\times10^{7}$ | 1·08E-04 | 2·07E-05 | 4·13E-04 | 6·18E-05 | 1·72E-04 | 4·37E-04 | 4·37E-04 |
| 367 | $2\times10^{7}$ | 1·88E-04 | 2·77E-04 | 2·25E-04 | 1·69E-05 | 1·24E-04 | 5·59E-04 | 5·59E-04 |
| 503 | $2\times10^{7}$ | 1·03E-04 | 1·14E-04 | 4·72E-05 | 1·24E-04 | | 1·56E-04 | 1·56E-04 |
| 501 | $2\times10^{7}$ | 9·77E-05 | 1·03E-04 | | 1·61E-04 | 4·72E-05 | 2·34E-04 | 2·34E-04 |
| 751 | $2\times10^{7}$ | 2·35E-04 | 8·98E-05 | 1·26E-03 | 3·15E-04 | 3·10E-04 | 3·72E-04 | 1·26E-03 |
| 546 | $2\times10^{7}$ | 1·81E-04 | 5·13E-04 | 1·36E-04 | 3·36E-04 | 2·94E-04 | 3·30E-04 | 5·13E-04 |
| 945 | $2\times10^{7}$ | 1·40E-04 | 1·56E-04 | 5·55E-04 | 2·26E-04 | 2·85E-04 | 1·79E-04 | 5·55E-04 |
| 802 | $2\times10^{7}$ | 5·38E-05 | 1·89E-04 | 2·00E-05 | 2·83E-05 | 6·11E-05 | 1·13E-04 | 1·89E-04 |
| 960 | $2\times10^{7}$ | 3·17E-04 | 1·28E-04 | 1·73E-03 | 5·72E-05 | 8·98E-05 | 1·57E-04 | 1·73E-03 |
| 375 | $2\times10^{7}$ | 8·14E-04 | 1·28E-04 | 2·61E-03 | 6·38E-04 | 7·86E-05 | 2·49E-04 | 2·61E-03 |
| 350 | $2\times10^{7}$ | 7·13E-05 | 2·54E-05 | 1·73E-04 | 4·79E-04 | 1·17E-04 | 9·09E-04 | 9·09E-04 |
| 1111 | $2\times10^{7}$ | 2·83E-05 | 7·50E-05 | 8·23E-05 | 6·77E-05 | 8·98E-05 | 3·30E-04 | 3·30E-04 |
| 576 | $2\times10^{7}$ | 7·86E-05 | 5·80E-04 | 5·00E-04 | 2·83E-05 | 7·13E-05 | 3·64E-04 | 5·80E-04 |
| 520 | $2\times10^{7}$ | 5·84E-04 | 2·10E-04 | 2·26E-04 | 1·93E-04 | 8·54E-04 | 4·74E-04 | 8·54E-04 |

| Primary CT-B IgM (RAU) | | | | | | | |
|---|---|---|---|---|---|---|---|
| Dose interval (days) | Dose (CFU) | D1 | D7 | D15 | D29 | D180 | max |
| 389 | $10^{8}$ | | 6·19E-04 | | 1·91E-04 | 7·19E-05 | 6·19E-04 |
| 405 | $10^{8}$ | | 1·54E-05 | 5·17E-05 | | | 5·17E-05 |
| 322 | $10^{7}$ | 7·12E-07 | | 1·98E-04 | 3·13E-06 | 1·73E-05 | 1·98E-04 |
| 418 | $10^{7}$ | | | 5·17E-05 | 5·17E-05 | | 5·17E-05 |
| 378 | $10^{8}$ | | 3·48E-04 | | | | 3·48E-04 |
| 399 | $10^{8}$ | | 2·69E-04 | 2·95E-04 | 1·14E-03 | 5·14E-04 | 1·14E-03 |
| 568 | $10^{7}$ | | | | | | |
| 367 | $10^{8}$ | 3·13E-06 | 2·53E-04 | 1·42E-04 | | | 2·53E-04 |
| 503 | $10^{7}$ | 1·54E-05 | 1·01E-03 | 1·73E-04 | | | 1·01E-03 |
| 501 | $10^{8}$ | | 2·69E-04 | | 2·49E-04 | | 2·69E-04 |
| 751 | $10^{10}$ | | 8·99E-04 | | | 6·17E-06 | 8·99E-04 |
| 546 | $10^{8}$ | | 4·37E-04 | 1·60E-04 | | | 4·37E-04 |
| 945 | $10^{8}$ | | 4·25E-04 | 1·20E-05 | | | 4·25E-04 |
| 802 | $10^{10}$ | | 3·10E-05 | | | | 3·10E-05 |
| 960 | $10^{8}$ | 5·68E-03 | 2·34E-03 | 2·21E-02 | 1·62E-02 | 3·10E-05 | 2·21E-02 |
| 375 | $10^{5}$ | | | | 4·50E-03 | 5·38E-05 | 4·50E-03 |
| 350 | $10^{5}$ | 5·75E-05 | 1·19E-04 | 3·13E-06 | 9·17E-06 | 6·71E-04 | 6·71E-04 |
| 1111 | $10^{6}$ | | 3·60E-04 | | 3·10E-05 | | 3·60E-04 |
| 576 | $10^{7}$ | | 2·04E-04 | | 3·08E-04 | | 3·08E-04 |
| 520 | $10^{7}$ | | 1·17E-03 | 1·29E-03 | 9·18E-05 | | 1·29E-03 |

| Boost CT-B IgG (RAU) | | | | | | | | |
|---|---|---|---|---|---|---|---|---|
| **Dose interval (days)** | **Dose (CFU)** | **D1** | **D4** | **D7** | **D15** | **D29** | **D180** | **max** |
| 389 | $2\times10^7$ | | 1·34E-06 | 1·16E-03 | 4·12E-07 | 2·45E-07 | 1·66E-06 | 1·16E-03 |
| 405 | $2\times10^7$ | | | 2·28E-03 | 8·92E-05 | 1·16E-05 | 1·10E-06 | 2·28E-03 |
| 322 | $2\times10^7$ | | | | | | 6·44E-07 | 6·44E-07 |
| 418 | $2\times10^7$ | | | | | | 7·78E-07 | 7·78E-07 |
| 378 | $2\times10^7$ | | | 1·30E-03 | 3·36E-05 | 8·49E-06 | 8·98E-07 | 1·30E-03 |
| 399 | $2\times10^7$ | | | | | | 3·54E-07 | 3·54E-07 |
| 568 | $2\times10^7$ | | | 3·59E-03 | 2·85E-05 | | 1·75E-06 | 3·59E-03 |
| 367 | $2\times10^7$ | | 3·26E-06 | 4·68E-05 | | | 2·06E-06 | 4·68E-05 |
| 503 | $2\times10^7$ | | | 1·61E-05 | | | 1·14E-06 | 1·61E-05 |
| 501 | $2\times10^7$ | | | 1·79E-04 | 1·73E-05 | | 9·10E-07 | 1·79E-04 |
| 751 | $2\times10^7$ | 2·45E-06 | 2·91E-06 | 8·76E-04 | 4·79E-05 | 4·42E-06 | 2·51E-06 | 8·76E-04 |
| 546 | $2\times10^7$ | 9·92E-07 | 4·25E-06 | 2·72E-05 | 3·96E-06 | 2·86E-06 | 2·19E-06 | 2·72E-05 |
| 945 | $2\times10^7$ | 1·29E-06 | 1·75E-06 | 4·38E-04 | 4·47E-05 | 2·76E-06 | 7·82E-07 | 4·38E-04 |
| 802 | $2\times10^7$ | 7·78E-07 | 1·75E-06 | 6·02E-05 | 4·98E-06 | 2·09E-06 | 5·78E-07 | 6·02E-05 |
| 960 | $2\times10^7$ | 8·68E-07 | 7·78E-07 | 1·69E-03 | 2·58E-05 | 1·33E-05 | 1·97E-06 | 1·69E-03 |
| 375 | $2\times10^7$ | 1·02E-06 | 1·06E-06 | 1·01E-03 | 1·38E-05 | 2·38E-06 | 8·94E-07 | 1·01E-03 |
| 350 | $2\times10^7$ | 6·58E-07 | 1·11E-06 | 2·09E-05 | 5·48E-06 | 2·05E-06 | 1·41E-06 | 2·09E-05 |
| 1111 | $2\times10^7$ | 5·12E-07 | 4·72E-07 | 9·25E-07 | 9·12E-07 | 6·84E-07 | 7·66E-07 | 9·25E-07 |
| 576 | $2\times10^7$ | 5·65E-07 | 3·82E-06 | 2·70E-03 | 8·19E-06 | 3·70E-06 | 9·74E-07 | 2·70E-03 |
| 520 | $2\times10^7$ | 1·08E-04 | 1·67E-06 | 1·15E-06 | 1·06E-06 | 1·34E-06 | 2·14E-06 | 1·08E-04 |

| Primary CT-B IgG (RAU) | | | | | | | |
|---|---|---|---|---|---|---|---|
| **Dose interval (days)** | **Dose (CFU)** | **D1** | **D7** | **D15** | **D29** | **D180** | **max** |
| 389 | $10^8$ | 1·43E-06 | 5·76E-03 | 1·17E-04 | 9·93E-06 | | 5·76E-03 |
| 405 | $10^8$ | | 5·73E-06 | 1·10E-04 | 8·14E-06 | | 1·10E-04 |
| 322 | $10^7$ | | | | | | |
| 418 | $10^7$ | | | 2·91E-06 | 8·88E-06 | | 8·88E-06 |
| 378 | $10^8$ | 2·54E-07 | 7·28E-03 | 5·05E-05 | 5·81E-05 | | 7·28E-03 |
| 399 | $10^8$ | | | 5·88E-06 | 7·30E-05 | 4·85E-05 | 7·30E-05 |
| 568 | $10^7$ | | | | | | |
| 367 | $10^8$ | | | 1·86E-06 | 4·95E-07 | | 1·86E-06 |
| 503 | $10^7$ | | 2·67E-05 | 1·13E-05 | 1·87E-06 | | 2·67E-05 |
| 501 | $10^8$ | 5·85E-07 | 1·10E-03 | 3·31E-05 | | | 1·10E-03 |
| 751 | $10^{10}$ | | 4·75E-03 | 1·26E-04 | 9·00E-06 | | 4·75E-03 |
| 546 | $10^8$ | 1·28E-06 | 6·44E-03 | 2·31E-05 | 5·20E-07 | 1·09E-06 | 6·44E-03 |
| 945 | $10^8$ | | 4·20E-06 | 8·64E-05 | 5·53E-06 | | 8·64E-05 |
| 802 | $10^{10}$ | | 6·86E-04 | 1·98E-05 | | | 6·86E-04 |
| 960 | $10^8$ | | 1·34E-03 | 8·52E-05 | 4·68E-07 | | 1·34E-03 |
| 375 | $10^5$ | | | | 2·49E-05 | 1·15E-06 | 2·49E-05 |
| 350 | $10^5$ | | 3·60E-05 | 6·83E-05 | 8·26E-06 | 6·98E-07 | 6·83E-05 |
| 1111 | $10^6$ | | 3·46E-03 | 1·57E-07 | | | 3·46E-03 |
| 576 | $10^7$ | | 1·22E-03 | 1·35E-05 | 1·22E-05 | | 1·22E-03 |
| 520 | $10^7$ | | 4·69E-04 | 1·95E-04 | 9·07E-06 | | 4·69E-04 |

| Boost CT-B IgA (RAU) | | | | | | | | |
|---|---|---|---|---|---|---|---|---|
| **Dose interval (days)** | **Dose (CFU)** | **D1** | **D4** | **D7** | **D15** | **D29** | **D180** | **max** |
| 389 | $2\times10^{7}$ | 1·19E-04 | 2·59E-05 | 4·05E-03 | 1·41E-04 | 6·96E-05 | 7·43E-06 | 4·05E-03 |
| 405 | $2\times10^{7}$ | 2·50E-05 | 1·35E-05 | 2·12E-01 | 1·46E-03 | 7·54E-05 | 3·47E-05 | 2·12E-01 |
| 322 | $2\times10^{7}$ | 2·06E-05 | 1·15E-05 | 2·09E-06 | 1·39E-05 | 1·32E-05 | 2·91E-05 | 2·91E-05 |
| 418 | $2\times10^{7}$ | 4·68E-06 | 9·99E-06 | 1·75E-05 | 1·25E-05 | 2·41E-05 | 2·87E-05 | 2·87E-05 |
| 378 | $2\times10^{7}$ | 2·24E-05 | 5·79E-05 | 7·42E-03 | 3·83E-04 | 3·55E-05 | 3·11E-05 | 7·42E-03 |
| 399 | $2\times10^{7}$ | 3·24E-05 | 2·02E-05 | 2·40E-05 | 1·76E-05 | 4·67E-05 | 3·06E-05 | 4·67E-05 |
| 568 | $2\times10^{7}$ | 9·90E-06 | 1·20E-05 | 1·91E-02 | 7·67E-05 | 2·12E-05 | 2·22E-05 | 1·91E-02 |
| 367 | $2\times10^{7}$ | 3·40E-05 | 2·54E-05 | 6·63E-05 | 1·67E-05 | 4·71E-05 | 1·75E-04 | 1·75E-04 |
| 503 | $2\times10^{7}$ | 1·24E-05 | 1·96E-05 | 4·93E-05 | 1·62E-05 | 1·52E-05 | 3·69E-05 | 4·93E-05 |
| 501 | $2\times10^{7}$ | 4·46E-05 | 5·59E-05 | 7·01E-04 | 1·37E-05 | 5·92E-05 | | 7·01E-04 |
| 751 | $2\times10^{7}$ | 1·90E-05 | 3·53E-05 | 1·27E-03 | 1·13E-04 | 4·42E-05 | | 1·27E-03 |
| 546 | $2\times10^{7}$ | 2·87E-05 | 5·78E-04 | 3·34E-04 | 4·17E-05 | 4·36E-05 | | 5·78E-04 |
| 945 | $2\times10^{7}$ | 3·78E-05 | 7·18E-05 | 4·80E-02 | 9·20E-04 | 1·11E-04 | | 4·80E-02 |
| 802 | $2\times10^{7}$ | 7·61E-06 | 1·45E-05 | 3·45E-04 | 1·50E-04 | 1·60E-05 | | 3·45E-04 |
| 960 | $2\times10^{7}$ | 2·46E-05 | 4·33E-05 | 6·40E-03 | 6·93E-05 | 1·49E-05 | | 6·40E-03 |
| 375 | $2\times10^{7}$ | 4·54E-05 | 1·02E-04 | 1·26E-02 | 8·55E-04 | 2·34E-04 | | 1·26E-02 |
| 350 | $2\times10^{7}$ | 3·15E-05 | 3·14E-05 | 1·00E-04 | 4·54E-05 | 1·76E-05 | | 1·00E-04 |
| 1111 | $2\times10^{7}$ | 8·22E-06 | 1·13E-05 | 1·87E-05 | 2·41E-05 | 1·22E-05 | | 2·41E-05 |
| 576 | $2\times10^{7}$ | 3·84E-05 | 2·13E-05 | 1·10E-01 | 3·47E-05 | 3·73E-05 | | 1·10E-01 |
| 520 | $2\times10^{7}$ | 8·22E-04 | 2·15E-04 | 3·76E-05 | 8·27E-05 | 3·26E-05 | | 8·22E-04 |

| Primary CT-B IgA (RAU) | | | | | | | |
|---|---|---|---|---|---|---|---|
| **Dose interval (days)** | **Dose (CFU)** | **D1** | **D7** | **D15** | **D29** | **D180** | **max** |
| 389 | $10^{8}$ | 1·80E-05 | 1·13E-02 | 1·92E-04 | 1·69E-05 | 8·33E-06 | 1·13E-02 |
| 405 | $10^{8}$ | 9·38E-06 | 2·93E-06 | 3·55E-04 | 1·04E-04 | 2·37E-05 | 3·55E-04 |
| 322 | $10^{7}$ | 5·39E-07 | 1·70E-05 | 9·62E-06 | 7·46E-06 | 5·29E-06 | 1·70E-05 |
| 418 | $10^{7}$ | 2·00E-05 | 1·52E-05 | 1·02E-04 | 1·70E-04 | 1·53E-05 | 1·70E-04 |
| 378 | $10^{8}$ | 9·23E-06 | 9·34E-01 | 1·14E-04 | 2·17E-04 | 5·06E-06 | 9·34E-01 |
| 399 | $10^{8}$ | 2·02E-06 | 1·39E-04 | 2·29E-03 | 1·24E-03 | 1·40E-05 | 2·29E-03 |
| 568 | $10^{7}$ | 6·56E-06 | 9·23E-04 | 5·80E-05 | 3·16E-05 | 7·08E-06 | 9·23E-04 |
| 367 | $10^{8}$ | 5·39E-07 | 2·58E-06 | 8·38E-06 | 1·16E-05 | 1·73E-04 | 1·73E-04 |
| 503 | $10^{7}$ | 2·87E-06 | 4·06E-04 | 7·86E-05 | 1·58E-04 | 1·75E-06 | 4·06E-04 |
| 501 | $10^{8}$ | 1·42E-05 | 3·49E-03 | 6·67E-05 | 1·25E-04 | 1·14E-05 | 3·49E-03 |
| 751 | $10^{10}$ | | 3·65E-03 | 1·27E-04 | 4·77E-05 | 3·03E-06 | 3·65E-03 |
| 546 | $10^{8}$ | 7·30E-06 | 6·24E-02 | 1·03E-04 | 4·54E-05 | 1·69E-05 | 6·24E-02 |
| 945 | $10^{8}$ | 5·02E-06 | 1·24E-03 | 4·44E-03 | 1·01E-04 | 1·29E-06 | 4·44E-03 |
| 802 | $10^{10}$ | 1·60E-06 | 1·71E-02 | 1·06E-04 | | 5·35E-07 | 1·71E-02 |
| 960 | $10^{8}$ | 3·22E-05 | 1·86E-03 | 3·19E-04 | 1·23E-05 | 4·07E-06 | 1·86E-03 |
| 375 | $10^{5}$ | 2·96E-05 | 1·43E-06 | 1·33E-05 | 6·85E-05 | 1·02E-04 | 1·02E-04 |
| 350 | $10^{5}$ | 2·45E-06 | 2·56E-05 | 2·07E-04 | 1·69E-05 | 1·62E-05 | 2·07E-04 |
| 1111 | $10^{6}$ | 4·12E-05 | 4·13E-01 | 4·17E-04 | 5·17E-05 | 6·13E-06 | 4·13E-01 |
| 576 | $10^{7}$ | | 2·63E-02 | 3·45E-04 | 6·79E-06 | 2·25E-07 | 2·63E-02 |
| 520 | $10^{7}$ | 1·87E-05 | 8·05E-03 | 1·46E-03 | 7·99E-05 | 1·03E-05 | 8·05E-03 |

| Boost TcpA IgM (RAU) | | | | | | | | |
|---|---|---|---|---|---|---|---|---|
| Dose interval (days) | Dose (CFU) | D1 | D4 | D7 | D15 | D29 | D180 | max |
| 389 | $2\times10^7$ | | 1·14E-04 | 1·07E-02 | 1·70E-04 | | 1·49E-04 | 1·07E-02 |
| 405 | $2\times10^7$ | | | 1·00E-02 | | | 3·97E-05 | 1·00E-02 |
| 322 | $2\times10^7$ | | | 8·96E-03 | | | 5·06E-05 | 8·96E-03 |
| 418 | $2\times10^7$ | | | 1·74E-03 | | | 5·64E-05 | 1·74E-03 |
| 378 | $2\times10^7$ | | | 1·20E-02 | 6·61E-05 | | 6·53E-05 | 1·20E-02 |
| 399 | $2\times10^7$ | | | | | | 1·07E-04 | 1·07E-04 |
| 568 | $2\times10^7$ | | | 6·93E-02 | | | 1·86E-04 | 6·93E-02 |
| 367 | $2\times10^7$ | | | | | | 1·07E-04 | 1·07E-04 |
| 503 | $2\times10^7$ | 1·61E-04 | | | | | 1·21E-04 | 1·61E-04 |
| 501 | $2\times10^7$ | | | 2·16E-03 | | | 6·91E-04 | 2·16E-03 |
| 751 | $2\times10^7$ | 1·56E-04 | 5·06E-05 | 3·31E-02 | 1·93E-04 | 1·28E-04 | 4·93E-04 | 3·31E-02 |
| 546 | $2\times10^7$ | 4·51E-05 | 1·93E-04 | 1·24E-04 | 1·24E-04 | 2·94E-04 | 8·12E-04 | 8·12E-04 |
| 945 | $2\times10^7$ | 7·45E-05 | 1·00E-04 | 1·23E-02 | 2·08E-04 | 9·37E-05 | 1·88E-03 | 1·23E-02 |
| 802 | $2\times10^7$ | 1·04E-04 | 1·00E-04 | 8·08E-05 | 2·95E-05 | 1·58E-04 | 5·92E-04 | 5·92E-04 |
| 960 | $2\times10^7$ | 2·82E-05 | 9·04E-05 | 1·40E-02 | 4·51E-05 | 8·72E-05 | 1·18E-03 | 1·40E-02 |
| 375 | $2\times10^7$ | 3·97E-05 | 1·14E-04 | 2·09E-02 | 9·37E-05 | 5·93E-05 | 8·78E-04 | 2·09E-02 |
| 350 | $2\times10^7$ | 1·28E-04 | 6·83E-05 | 1·11E-03 | 1·75E-04 | 7·14E-05 | 3·04E-03 | 3·04E-03 |
| 1111 | $2\times10^7$ | 6·23E-05 | 5·64E-05 | 4·23E-05 | 4·51E-05 | 1·28E-04 | 6·80E-04 | 6·80E-04 |
| 576 | $2\times10^7$ | 2·95E-05 | 1·14E-04 | 9·41E-03 | 1·07E-05 | 8·40E-05 | 7·68E-04 | 9·41E-03 |
| 520 | $2\times10^7$ | 5·15E-04 | 1·07E-04 | 2·71E-05 | 8·40E-05 | 9·04E-05 | 1·12E-03 | 1·12E-03 |

| Primary TcpA IgM (RAU) | | | | | | | |
|---|---|---|---|---|---|---|---|
| Dose interval (days) | Dose (CFU) | D1 | D7 | D15 | D29 | D180 | max |
| 389 | $10^8$ | | 2·24E-05 | 1·01E-04 | 9·48E-06 | 5·93E-05 | 1·01E-04 |
| 405 | $10^8$ | 5·24E-05 | 2·40E-06 | 1·76E-06 | 2·24E-05 | | 5·24E-05 |
| 322 | $10^7$ | 3·85E-06 | 3·09E-02 | 1·70E-04 | 9·27E-06 | | 3·09E-02 |
| 418 | $10^7$ | | 1·38E-05 | 4·90E-06 | 8·49E-05 | 1·40E-06 | 8·49E-05 |
| 378 | $10^8$ | | 4·70E-05 | 1·42E-07 | | 3·36E-05 | 4·70E-05 |
| 399 | $10^8$ | | 3·06E-05 | 1·38E-05 | 7·80E-07 | 7·85E-04 | 7·85E-04 |
| 568 | $10^7$ | | 6·06E-02 | | | 9·55E-03 | 6·06E-02 |
| 367 | $10^8$ | | 4·92E-04 | 6·66E-05 | | 8·53E-05 | 4·92E-04 |
| 503 | $10^7$ | 1·76E-06 | 1·76E-03 | 7·80E-07 | 6·07E-05 | 7·32E-05 | 1·76E-03 |
| 501 | $10^8$ | 4·42E-07 | 2·62E-04 | 1·76E-06 | 1·54E-05 | 4·02E-05 | 2·62E-04 |
| 751 | $10^{10}$ | | 3·10E-03 | | 3·24E-05 | | 3·10E-03 |
| 546 | $10^8$ | 6·07E-05 | 3·74E-05 | 8·21E-06 | 9·81E-05 | 1·63E-05 | 9·81E-05 |
| 945 | $10^8$ | 1·05E-04 | | | | | 1·05E-04 |
| 802 | $10^{10}$ | | 2·27E-04 | | | 4·38E-05 | 2·27E-04 |
| 960 | $10^8$ | 1·15E-02 | 1·67E-03 | 5·36E-01 | 3·18E-01 | 1·64E-04 | 5·36E-01 |
| 375 | $10^5$ | | 1·68E-03 | 5·57E-04 | | 6·53E-05 | 1·68E-03 |
| 350 | $10^5$ | 1·40E-06 | 1·23E-03 | 2·44E-04 | 1·63E-05 | 7·45E-05 | 1·23E-03 |
| 1111 | $10^6$ | | 1·92E-03 | | 2·46E-04 | | 1·92E-03 |
| 576 | $10^7$ | 2·40E-06 | 6·07E-05 | 1·70E-05 | | 6·17E-05 | 6·17E-05 |
| 520 | $10^7$ | 4·02E-05 | 2·97E-04 | 2·16E-05 | 5·46E-06 | 2·49E-06 | 2·97E-04 |

| Boost TcpA IgG (RAU) | | | | | | | | |
|---|---|---|---|---|---|---|---|---|
| **Dose interval (days)** | **Dose (CFU)** | **D1** | **D4** | **D7** | **D15** | **D29** | **D180** | **max** |
| 389 | $2\times10^{7}$ | 1·71E-05 | 9·47E-06 | 1·60E-02 | 6·62E-05 | 3·57E-05 | 1·13E-05 | 1·60E-02 |
| 405 | $2\times10^{7}$ | 6·97E-07 | | 2·90E-03 | 1·45E-05 | 1·45E-05 | 8·77E-06 | 2·90E-03 |
| 322 | $2\times10^{7}$ | 1·81E-06 | 9·47E-06 | | | 1·81E-06 | 6·23E-06 | 9·47E-06 |
| 418 | $2\times10^{7}$ | | 7·82E-06 | 6·46E-05 | | 1·28E-05 | 5·98E-06 | 6·46E-05 |
| 378 | $2\times10^{7}$ | | 1·62E-05 | 2·75E-02 | 7·54E-05 | 3·37E-04 | 4·78E-05 | 2·75E-02 |
| 399 | $2\times10^{7}$ | | | | 1·21E-06 | nd | 8·77E-06 | 8·77E-06 |
| 568 | $2\times10^{7}$ | | 1·11E-05 | 1·37E-02 | 4·79E-05 | 1·88E-05 | 5·73E-06 | 1·37E-02 |
| 367 | $2\times10^{7}$ | | | 1·81E-06 | 2·13E-05 | 2·93E-07 | 1·92E-05 | 2·13E-05 |
| 503 | $2\times10^{7}$ | | 6·20E-06 | 4·79E-05 | | 1·79E-05 | 1·13E-05 | 4·79E-05 |
| 501 | $2\times10^{7}$ | 7·82E-06 | | 5·33E-03 | 1·88E-05 | 1·11E-05 | 4·40E-05 | 5·33E-03 |
| 751 | $2\times10^{7}$ | 1·44E-05 | 1·18E-05 | 6·77E-02 | 3·18E-04 | 2·78E-05 | 2·82E-05 | 6·77E-02 |
| 546 | $2\times10^{7}$ | 1·13E-05 | 3·82E-05 | 2·66E-04 | 2·22E-05 | 2·61E-05 | 4·79E-05 | 2·66E-04 |
| 945 | $2\times10^{7}$ | 1·13E-05 | 1·13E-05 | 9·30E-03 | 3·06E-05 | 1·94E-05 | 3·93E-05 | 9·30E-03 |
| 802 | $2\times10^{7}$ | 1·11E-05 | 5·98E-06 | 1·18E-04 | 9·79E-06 | 1·31E-05 | 3·83E-05 | 1·18E-04 |
| 960 | $2\times10^{7}$ | 8·71E-06 | 9·02E-06 | 1·98E-03 | 2·33E-05 | 9·53E-06 | 5·57E-05 | 1·98E-03 |
| 375 | $2\times10^{7}$ | 4·71E-06 | 1·54E-05 | 2·57E-03 | 6·15E-05 | 1·84E-05 | 5·47E-05 | 2·57E-03 |
| 350 | $2\times10^{7}$ | 4·18E-05 | 1·13E-05 | 1·31E-03 | 2·99E-05 | 3·39E-05 | 5·67E-05 | 1·31E-03 |
| 1111 | $2\times10^{7}$ | 5·98E-06 | 6·23E-06 | 6·99E-06 | 1·39E-05 | 5·22E-06 | 2·91E-05 | 2·91E-05 |
| 576 | $2\times10^{7}$ | 4·46E-06 | 1·08E-05 | 3·88E-03 | 1·46E-05 | 6·74E-06 | 4·16E-05 | 3·88E-03 |
| 520 | $2\times10^{7}$ | 4·73E-05 | 1·64E-05 | 1·08E-05 | 1·54E-05 | 1·23E-05 | 6·77E-05 | 6·77E-05 |

| Primary TcpA IgG (RAU) | | | | | | | |
|---|---|---|---|---|---|---|---|
| **Dose interval (days)** | **Dose (CFU)** | **D1** | **D7** | **D15** | **D29** | **D180** | **max** |
| 389 | $10^{8}$ | | 4·66E-06 | | | 1·73E-05 | 1·73E-05 |
| 405 | $10^{8}$ | | 7·38E-06 | 4·66E-06 | 2·92E-06 | 1·65E-05 | 1·65E-05 |
| 322 | $10^{7}$ | 3·44E-05 | 2·98E-06 | | 9·42E-06 | 2·71E-05 | 3·44E-05 |
| 418 | $10^{7}$ | 1·21E-05 | 1·29E-05 | | | 1·49E-05 | 1·49E-05 |
| 378 | $10^{8}$ | 2·55E-05 | 1·10E-05 | 1·33E-05 | 1·98E-05 | 1·17E-05 | 2·55E-05 |
| 399 | $10^{8}$ | | 1·93E-06 | | 8·11E-06 | 1·73E-05 | 1·73E-05 |
| 568 | $10^{7}$ | | 3·76E-03 | | | 1·54E-03 | 3·76E-03 |
| 367 | $10^{8}$ | 2·47E-05 | 1·65E-05 | 2·63E-05 | 1·17E-05 | 2·14E-05 | 2·63E-05 |
| 503 | $10^{7}$ | 1·12E-06 | 2·37E-05 | 2·85E-07 | | 8·64E-06 | 2·37E-05 |
| 501 | $10^{8}$ | | 4·05E-06 | 5·05E-07 | 1·12E-06 | 1·82E-05 | 1·82E-05 |
| 751 | $10^{10}$ | | 3·38E-06 | 1·76E-06 | 1·81E-05 | 1·06E-06 | 1·81E-05 |
| 546 | $10^{8}$ | | 4·66E-06 | | | 3·92E-05 | 3·92E-05 |
| 945 | $10^{8}$ | 2·22E-06 | 1·06E-06 | 2·22E-06 | | 9·41E-08 | 2·22E-06 |
| 802 | $10^{10}$ | 4·84E-07 | | 1·06E-06 | | 4·28E-06 | 4·28E-06 |
| 960 | $10^{8}$ | | 1·21E-05 | | 1·06E-06 | 4·84E-07 | 1·21E-05 |
| 375 | $10^{5}$ | 2·95E-05 | | 3·20E-05 | 4·40E-05 | 4·21E-06 | 4·40E-05 |
| 350 | $10^{5}$ | 5·68E-06 | 8·64E-06 | 1·49E-05 | 1·02E-05 | 3·59E-05 | 3·59E-05 |
| 1111 | $10^{6}$ | 3·61E-06 | 5·29E-05 | | | | 5·29E-05 |
| 576 | $10^{7}$ | | 1·01E-06 | 5·60E-07 | | 2·79E-05 | 2·79E-05 |
| 520 | $10^{7}$ | 3·36E-05 | 2·38E-05 | 1·65E-05 | 1·17E-05 | | 3·36E-05 |

| Boost TcpA IgA (RAU) | | | | | | | | |
|---|---|---|---|---|---|---|---|---|
| Dose interval (days) | Dose (CFU) | D1 | D4 | D7 | D15 | D29 | D180 | max |
| 389 | $2\times10^7$ | 6·14E-05 | 5·81E-05 | 3·98E-02 | 1·03E-04 | 2·37E-05 | 1·76E-05 | 3·98E-02 |
| 405 | $2\times10^7$ | 5·07E-05 | 2·48E-05 | 1·15E-01 | 7·34E-05 | 7·65E-05 | 6·40E-05 | 1·15E-01 |
| 322 | $2\times10^7$ | 4·86E-05 | 3·24E-05 | 6·35E-04 | 2·70E-05 | 5·22E-05 | 3·95E-05 | 6·35E-04 |
| 418 | $2\times10^7$ | 2·59E-05 | 3·80E-05 | 1·86E-03 | 3·99E-05 | 4·76E-05 | 4·96E-05 | 1·86E-03 |
| 378 | $2\times10^7$ | 3·39E-05 | 6·39E-05 | 1·94E-01 | 1·84E-04 | 5·32E-05 | 8·54E-05 | 1·94E-01 |
| 399 | $2\times10^7$ | 3·54E-05 | 6·33E-05 | 3·59E-05 | 1·91E-05 | 3·41E-05 | 4·72E-05 | 6·33E-05 |
| 568 | $2\times10^7$ | 2·81E-05 | 1·08E-05 | 3·74E-02 | 5·83E-05 | 4·12E-05 | 7·85E-05 | 3·74E-02 |
| 367 | $2\times10^7$ | 4·57E-05 | 4·76E-05 | 1·39E-04 | 3·82E-05 | 4·38E-05 | 9·81E-05 | 1·39E-04 |
| 503 | $2\times10^7$ | 2·57E-05 | 5·03E-05 | 4·67E-04 | 3·71E-05 | 5·74E-05 | 8·53E-05 | 4·67E-04 |
| 501 | $2\times10^7$ | 5·45E-05 | 3·02E-05 | 1·71E-02 | 2·59E-05 | 3·09E-05 | 9·37E-05 | 1·71E-02 |
| 751 | $2\times10^7$ | 4·88E-05 | 4·79E-05 | 5·19E-02 | 1·20E-04 | 3·11E-05 | 6·46E-05 | 5·19E-02 |
| 546 | $2\times10^7$ | 9·89E-04 | 3·09E-04 | 5·18E-03 | 9·75E-05 | 8·82E-05 | | 5·18E-03 |
| 945 | $2\times10^7$ | 7·94E-05 | 9·27E-05 | 1·94E-02 | 8·34E-05 | 8·14E-05 | | 1·94E-02 |
| 802 | $2\times10^7$ | 2·32E-05 | 4·46E-05 | 2·73E-04 | 2·58E-05 | 2·69E-05 | 3·02E-05 | 2·73E-04 |
| 960 | $2\times10^7$ | 7·56E-05 | 1·47E-04 | 1·14E-02 | 1·51E-04 | 4·74E-05 | 6·30E-04 | 1·14E-02 |
| 375 | $2\times10^7$ | 2·82E-05 | 9·90E-05 | 8·40E-02 | 3·16E-04 | 3·04E-04 | 5·18E-04 | 8·40E-02 |
| 350 | $2\times10^7$ | 8·75E-05 | 1·15E-04 | 6·00E-03 | 8·92E-05 | 4·04E-05 | 4·22E-04 | 6·00E-03 |
| 1111 | $2\times10^7$ | 2·49E-05 | 2·71E-05 | 9·53E-04 | 4·45E-05 | 3·57E-05 | 2·14E-04 | 9·53E-04 |
| 576 | $2\times10^7$ | 1·90E-05 | 4·44E-05 | 1·94E-02 | 6·07E-05 | 1·19E-04 | | 1·94E-02 |
| 520 | $2\times10^7$ | 9·93E-05 | 5·66E-05 | 5·51E-05 | 3·91E-05 | 3·77E-05 | 3·44E-04 | 3·44E-04 |

| Primary TcpA IgA (RAU) | | | | | | | |
|---|---|---|---|---|---|---|---|
| Dose interval (days) | Dose (CFU) | D1 | D7 | D15 | D29 | D180 | max |
| 389 | $10^8$ | 5·71E-05 | 2·08E-04 | 1·02E-04 | 6·38E-05 | 8·10E-08 | 2·08E-04 |
| 405 | $10^8$ | 1·53E-05 | 5·14E-05 | 2·37E-05 | 4·60E-05 | 8·37E-05 | 8·37E-05 |
| 322 | $10^7$ | 5·81E-06 | 7·03E-05 | 5·62E-05 | 3·05E-05 | 2·47E-05 | 7·03E-05 |
| 418 | $10^7$ | 6·73E-05 | 3·12E-05 | 5·31E-05 | 2·09E-05 | 1·70E-05 | 6·73E-05 |
| 378 | $10^8$ | 7·03E-06 | 1·35E-04 | 1·54E-05 | 1·40E-04 | 2·25E-05 | 1·40E-04 |
| 399 | $10^8$ | 2·18E-05 | 9·49E-05 | 3·78E-05 | 3·57E-05 | 2·18E-05 | 9·49E-05 |
| 568 | $10^7$ | 2·78E-05 | 2·02E-05 | 1·05E-05 | 3·22E-05 | 2·62E-05 | 3·22E-05 |
| 367 | $10^8$ | 3·12E-05 | 3·69E-05 | 5·34E-05 | 4·04E-05 | 1·06E-05 | 5·34E-05 |
| 503 | $10^7$ | 2·80E-05 | 4·86E-04 | 8·07E-05 | 1·85E-04 | 6·37E-06 | 4·86E-04 |
| 501 | $10^8$ | 1·20E-05 | 3·12E-05 | | 2·48E-05 | 3·96E-05 | 3·96E-05 |
| 751 | $10^{10}$ | 4·01E-07 | 1·11E-04 | 2·99E-06 | 2·12E-05 | 8·31E-07 | 1·11E-04 |
| 546 | $10^8$ | 2·96E-05 | 4·60E-05 | 3·43E-05 | 5·98E-05 | 2·46E-05 | 5·98E-05 |
| 945 | $10^8$ | 5·65E-06 | 1·45E-05 | 5·95E-05 | 1·19E-05 | 9·66E-06 | 5·95E-05 |
| 802 | $10^{10}$ | 2·03E-07 | 2·37E-05 | 1·52E-06 | | 1·60E-06 | 2·37E-05 |
| 960 | $10^8$ | 6·52E-05 | 1·73E-04 | 3·65E-05 | 3·77E-05 | 7·93E-06 | 1·73E-04 |
| 375 | $10^5$ | 2·03E-06 | 7·72E-06 | 7·03E-06 | 6·37E-06 | 7·48E-05 | 7·48E-05 |
| 350 | $10^5$ | 1·76E-05 | 1·99E-05 | 4·74E-05 | 5·62E-05 | 6·34E-04 | 6·34E-04 |
| 1111 | $10^6$ | 8·15E-06 | 2·86E-04 | 6·55E-06 | 1·21E-05 | 7·00E-06 | 2·86E-04 |
| 576 | $10^7$ | 2·31E-05 | 3·05E-05 | 2·31E-05 | 2·87E-05 | 2·25E-05 | 3·05E-05 |
| 520 | $10^7$ | 3·42E-05 | 4·83E-05 | 3·48E-05 | 6·54E-05 | 1·33E-05 | 6·54E-05 |

**Table S7: Quantification of PanChol bacteria in fecal samples after primary and booster vaccination.** Blue: last observation carried forward; orange: negative rectal swab.

| Boost: Abundance of PanChol (CFU/g) | | | | | | |
|---|---|---|---|---|---|---|
| **Dose interval (days)** | **Dose (CFU)** | **D2** | **D3** | **D4** | **D5** | **max** |
| 322 | $2{\times}10^7$ | 9·00E+01 | 3·20E+03 | 9·00E+01 | 9·00E+01 | 3·20E+03 |
| 350 | $2{\times}10^7$ | 9·00E+01 | 9·00E+01 | 9·00E+01 | 9·00E+01 | 9·00E+01 |
| 367 | $2{\times}10^7$ | 9·00E+01 | 9·00E+01 | 9·00E+01 | 9·00E+01 | 9·00E+01 |
| 375 | $2{\times}10^7$ | 2·00E+02 | 1·18E+04 | 9·00E+01 | 9·00E+01 | 1·18E+04 |
| 378 | $2{\times}10^7$ | 9·00E+01 | 9·00E+01 | 9·00E+01 | 9·00E+01 | 9·00E+01 |
| 389 | $2{\times}10^7$ | 7·10E+03 | 5·85E+04 | 6·20E+05 | 3·20E+04 | 6·20E+05 |
| 399 | $2{\times}10^7$ | 9·00E+01 | | 9·00E+01 | 9·00E+01 | 9·00E+01 |
| 405 | $2{\times}10^7$ | 3·00E+02 | 1·83E+07 | 4·55E+04 | 6·60E+03 | 1·83E+07 |
| 418 | $2{\times}10^7$ | 2·40E+03 | 2·95E+03 | 9·00E+01 | 9·00E+01 | 2·95E+03 |
| 501 | $2{\times}10^7$ | 9·00E+01 | 9·00E+01 | 9·00E+01 | 9·00E+01 | 9·00E+01 |
| 503 | $2{\times}10^7$ | 9·00E+01 | 9·00E+01 | 9·00E+01 | 9·00E+01 | 9·00E+01 |
| 520 | $2{\times}10^7$ | | 9·00E+01 | 9·00E+01 | 9·00E+01 | 9·00E+01 |
| 546 | $2{\times}10^7$ | 9·00E+01 | 9·00E+01 | 9·00E+01 | 9·00E+01 | 9·00E+01 |
| 568 | $2{\times}10^7$ | 2·00E+02 | 1·60E+05 | 2·70E+05 | 3·35E+03 | 2·70E+05 |
| 576 | $2{\times}10^7$ | 9·00E+01 | 9·00E+01 | 9·00E+01 | 9·00E+01 | 9·00E+01 |
| 751 | $2{\times}10^7$ | 9·00E+01 | 9·00E+01 | 9·00E+01 | 9·00E+01 | 9·00E+01 |
| 802 | $2{\times}10^7$ | 9·00E+01 | 9·00E+01 | 9·00E+01 | 9·00E+01 | 9·00E+01 |
| 945 | $2{\times}10^7$ | 9·00E+01 | 9·00E+01 | 9·00E+01 | 9·00E+01 | 9·00E+01 |
| 960 | $2{\times}10^7$ | 9·00E+01 | 9·00E+01 | 9·00E+01 | 9·00E+01 | 9·00E+01 |
| 1111 | $2{\times}10^7$ | 9·00E+01 | 9·00E+01 | 9·00E+01 | 9·00E+01 | 9·00E+01 |

| Primary: Abundance of PanChol (CFU/g) | | | | | | |
|---|---|---|---|---|---|---|
| **Dose interval (days)** | **Dose (CFU)** | **D2** | **D3** | **D4** | **D5** | **max** |
| 322 | $10^7$ | 9·00E+01 | 1·40E+06 | 5·10E+03 | 2·00E+02 | 1·40E+06 |
| 350 | $10^5$ | 1·90E+04 | 2·24E+07 | 2·90E+06 | 3·99E+06 | 2·24E+07 |
| 367 | $10^8$ | 9·00E+01 | 1·30E+03 | 2·30E+05 | 1·30E+03 | 2·30E+05 |
| 375 | $10^5$ | 9·00E+01 | 9·64E+06 | 1·90E+05 | 7·00E+02 | 9·64E+06 |
| 378 | $10^8$ | 2·10E+03 | 5·80E+04 | 2·00E+02 | 9·00E+01 | 5·80E+04 |
| 389 | $10^8$ | 2·01E+06 | 7·40E+06 | 4·20E+06 | 5·80E+06 | 7·40E+06 |
| 399 | $10^8$ | 9·00E+01 | 9·00E+01 | 9·00E+01 | 9·00E+01 | 9·00E+01 |
| 405 | $10^8$ | 1·00E+02 | 9·50E+05 | 3·80E+06 | 2·00E+05 | 3·80E+06 |
| 418 | $10^7$ | 1·20E+03 | 3·76E+07 | 3·50E+06 | 8·50E+03 | 3·76E+07 |
| 501 | $10^8$ | | 9·00E+01 | | 1·50E+05 | 1·50E+05 |
| 503 | $10^7$ | 4·00E+02 | 1·80E+05 | 9·30E+03 | 4·10E+03 | 1·80E+05 |
| 520 | $10^7$ | 9·00E+01 | | | 1·00E+02 | 1·00E+02 |
| 546 | $10^8$ | 8·00E+03 | 9·00E+01 | 1·00E+02 | 8·00E+02 | 8·00E+03 |
| 568 | $10^7$ | 9·00E+01 | 5·00E+04 | 2·42E+05 | 4·30E+05 | 4·30E+05 |
| 576 | $10^7$ | 1·00E+02 | 6·60E+03 | 9·00E+01 | 2·08E+06 | 2·08E+06 |
| 751 | $10^{10}$ | 1·10E+06 | 1·10E+04 | 9·70E+05 | 2·10E+04 | 1·10E+06 |
| 802 | $10^{10}$ | 3·30E+04 | 3·67E+07 | 1·93E+04 | 1·29E+05 | 3·67E+07 |
| 945 | $10^8$ | 1·00E+02 | 3·00E+02 | 4·00E+02 | 3·30E+04 | 3·30E+04 |
| 960 | $10^8$ | 3·50E+04 | 4·90E+04 | 1·80E+03 | 2·41E+04 | 4·90E+04 |
| 1111 | $10^6$ | 9·00E+01 | 1·00E+02 | 5·80E+03 | 4·00E+02 | 5·80E+03 |

| | |
|---|---|
| (blue) | last observation carried forward |
| (orange) | negative rectal swab |
